# Intensity-specific leisure time-physical activity and all-cause, cardiovascular, and cancer mortality in 3.36 million adults from 17 countries: a systematic review, meta-analysis, and individual participant data pooled analysis

**DOI:** 10.1101/2024.10.23.24315963

**Authors:** Matthew N. Ahmadi, Gerson Ferrari, Leandro F.M. Rezende, Gregore Mielke, Ruth Brady, Susan Paudel, Tze Rui Ma, Po-Wen Ku, Raaj Biswas, Nicholas A. Koemel, Emmanuel Stamatakis

## Abstract

**Background:** Studies examining the associations of intensity-specific leisure time physical activity duration with all-cause, cardiovascular disease (CVD), and cancer mortality are scarce and no quantitative or dose-response meta-analysis has been published.

**Objective:** We examined the associations of moderate, vigorous, and moderate to vigorous leisure time physical activity duration with all-cause, CVD, and cancer mortality, using aggregate and individual participant data.

**Methods:** We performed a systematic review and meta-analysis of both published and unpublished cohort studies that included data on intensity-specific leisure time physical activity. Hazard ratios (HR) were calculated by comparing high versus low levels of physical activity. We also harmonized and pooled individual participant data from unpublished large cohorts to assess dose-response associations with the same three mortality outcomes, as retrieved from National Death Registries.

**Results:** A total of 3.36 million participants across 25 cohorts and 17 countries, corresponding to 247,463 all-cause, 70,204 CVD, and 76,294 cancer deaths were included in our aggregate meta-analysis. Compared to low physical activity, the association of high moderate intensity leisure time physical activity with mortality ranged from an HR of 0.84 (95% CI= 0.79, 0.89) for all-cause mortality to 0.90 (0.86, 0.95) for cancer mortality; and vigorous intensity from 0.86 (0.79, 0.93) for all-cause mortality to 0.88 (0.83, 0.91) for cancer mortality.

Our pooled individual participant data analysis included 967,184 participants with an average follow-up time of 12.2 (SD= 4.7) years and 60,206 all-cause, 11,525 CVD, and 23,740 cancer deaths. The dose-response analysis showed a general L-shaped association across each outcome. For all-cause mortality, compared to the reference group with no leisure time activity, the minimal and optimal doses of vigorous intensity were 60 mins/week (0.86 [0.84, 0.89]) and 200 mins/week (0.69 [0.67, 0.71]), respectively. For moderate intensity, the corresponding doses were 100 mins/week (0.88 [0.86, 0.90]) and 340 mins/week (0.77 [0.75, 0.79]).

**Conclusions:** Our meta-analysis shows distinct differential associations of moderate and vigorous physical activity with all-cause, cardiovascular, and cancer mortality risk. Improvements in leisure time physical activity approximately equivalent to 60 mins/week of vigorous or 100 mins/week of moderate activity, may be linked with measurable health benefits. Our findings, synthesized uniquely through aggregated and pooled individual participant meta-analyses offer novel evidence to guide decisions on contents of leisure time physical activity focused interventions and preventive guidelines.

## Introduction

Cardiovascular disease (CVD) and cancer are the two leading causes of morbidity and mortality globally^1^. Physical inactivity is a primary risk factor for both diseases^2^. Despite significant public health and clinical efforts, global physical activity levels have shown little improvement^3^. The 2020 World Health Organization (WHO) Guidelines Development Group highlighted the lack of evidence on the differential effects of physical activity intensity—moderate versus vigorous—on mortality and disease risk^4^. Current WHO guidelines recommend 150-300 minutes of moderate-intensity or 75-150 minutes of vigorous-intensity physical activity per week, or a combination of both. This 2:1 ratio of vigorous to moderate activity is rooted in the assumption vigorous activity (>6 MET) requires twice the energy expenditure of moderate activity (3-6 MET)^5^. Although this assumption is widely used, it has not been empirically derived and has yet to be evaluated at the population level, despite the availability of international population cohorts. Addressing these gaps is vital for refining public health policies and improving patient care in the prevention of CVD and cancer, beyond the crude “more is better” approach.

Meta-analyses address concerns and challenges of generalisability and robustness by pooling studies from multiple cohorts. Pooled analyses have typically been heterogeneous due to differences in study design and contrasts examined (e.g.: tertiles vs quintiles). Such heterogeneity can attenuate risk estimates and mask true underlying associations. Individual participant data meta-analyses, that harmonises data at the individual level across cohorts, mitigates issues of heterogeneity and offers flexibility to understand non-linear dose-response associations. Individual participant data meta-analyses can therefore provide complementary information to pooled aggregate meta-analyses.

Previous aggregate meta-analyses have shown an inverse association between the combined duration of leisure time moderate to vigorous physical activity (MVPA) with mortality and disease risk^6^ ^7^. Extending on this, individual cohort studies have assessed the risk differences between separate contributions of moderate activity and vigorous activity, reporting an approximate 25% to 50% equivalent of vigorous activity duration to observe similar risk reductions as combined MVPA with all-cause and cause-specific mortality^8^ ^9^. Notably, recent narrative reviews^10^ and population cohort studies^11^ ^12^ have indicated the risk differences associated with vigorous and moderate intensity physical activity may be larger than previous guidelines have suggested. To address these gaps, dose-response meta-analyses of individual participant intensity data synthesised with traditional meta-analyses of published data may optimally inform public health and clinical prevention strategies.

We carried out a systematic review and meta-analysis of published and unpublished cohort studies to assess the association of leisure time MVPA duration, and its constituent components (moderate activity and vigorous activity), with all-cause, CVD, and cancer mortality. We further provide a complementary pooled individual participant data analysis of large unpublished cohorts to examine the dose-response associations between the duration of intensity-specific leisure time physical activity with the same three mortality outcomes.

## Methods

### Meta-analysis of published studies

#### Literature search and study selection

This systematic review was registered with the PROSPERO database (CRD42022323901) and the reporting followed the Meta-analysis Of Observational Studies in Epidemiology (MOOSE) checklist recommendations ^13^.

We conducted a broad search, without restriction of date of publication and language in Medline (PubMed), Scopus, Web of Science and Embase in August 2024. The search was restricted to adult (≥18 years at baseline) human studies. In addition, reference lists of studies included in systematic review and reviews about physical activity and mortality were screened for additional studies using search terms related to exposure (“leisure-time physical activity”, “recreational physical activity”, “physical activity”, “exercise”, “physical exercise”, “physical fitness”, “walk”, “jog”, “run”) and outcome (“mortality”, “all-cause mortality”, “cardiovascular mortality”, “cancer mortality”). **Supplemental Text 1** provides the full list of search terms used in each database Prospective or longitudinal studies (cohort) studies that evaluated the association between lower intensity or moderate intensity or lower weekly duration of leisure-time physical activity with all-cause, cardiovascular and cancer mortality, were eligible for inclusion. To be included studies were required to have information on leisure-time physical activity questionnaire in adults (≥18 years at baseline) and report hazard ratio, relative risk or odds ratio for intensity of leisure time physical activity with mortality adjusted for total physical activity. Theoretically, randomized clinical trials were eligible for our analysis, but no such studies were identified, probably due to the absence of such evidence, that is likely due to the prohibitively large resources required for setting up, running, and completing such studies^14^. Congress abstracts and narrative reviews were ineligible. Studies of non-domain specific device measured physical activity, clinical/diseased cohorts of either impatient or free-living (i.e. cohorts that exclusively comprise people with a specific disease) were also excluded. Selection of articles first involved reading and evaluating titles and abstracts considering the scope of the systematic review using Covidence. Then, the full texts were read for the final selection. In both stages, the articles were selected by two researchers (RB and SP, NK, RM, RKB) and compared; in cases of disagreement, a third researcher (GF) was asked to arbitrate.

#### Data extraction

Data were extracted on health status (e.g., prevalent cardiovascular disease or cancer), general study characteristics (publication date, sample size, country), general participant characteristics, sex, length of follow-up of the study, assessment of exposure, solely leisure-time physical activity vs mostly leisure-time physical activity, age group (18-40 vs 40+) scores obtained in ROBINS-E). When the information was unavailable in the study, the authors were contacted for further clarification.

#### Risk of bias assessment

The ROBINS-E tool (“Risk Of Bias In Non-randomised Studies -of Exposures”) includes seven domains to assess confounding factors that have the potential to introduce material bias into an estimated effect^15^. Domain level judgements about risk of bias are conceived hierarchically: 1) Low risk of bias (where the study is comparable to a well-performed randomized trial with regard to that domain); 2) Moderate risk of bias (where the study is sound for a non-randomized study but cannot be considered comparable to a well-performed randomized trial with regard to that domain); 3) Serious risk of bias (the study has some important problems in that domain); 4) Critical risk of bias (the study is too problematic in that domain to provide any useful evidence on the effects of the intervention); 5) No information on which to base a judgement about risk of bias for that domain. Disagreements were arbitrated by a third researcher (GM).

### Aggregate meta-analysis

For our aggregate meta-analysis of previously published cohort studies, we considered high leisure time physical activity as the group with the highest duration of leisure-time physical activity performed in the study, and “Low” as the group that reported none or the lowest duration of leisure-time physical activity. Heterogeneity between studies was quantified using *I*^S^, Cochrańs Q statistics ^16^, and *τ*^S^ estimate. The systematic review protocol included assessing and sources of heterogeneity between studies. We also performed small study effects and funnel plots to assess publication bias (**Supplemental Figure 1)**

### Pooled individual participant data meta-analysis

We pooled individual participant data from 11 annual baseline data collections of the Health Survey for England (1994-2008)^17^, 3 baseline data collections for the Scottish Health Survey (1995-2003)^18^, 18 baseline data collections of the MJ Health Database in Taiwan (1998-2016)^19^, the UK Biobank (2006-2010)^20^, and the Taiwan Biobank (2008-2018)^21^. All cohorts used recall questionnaires to assess moderate and vigorous leisure-time physical activity duration. The methods used in the harmonisation of leisure-time physical activity are described fully in **Supplemental Text 2**. For each cohort, covariates were measured during clinic visits or home visits, and chosen a priori based on previous literature were harmonised for: age, sex, smoking status, alcohol consumption, body mass index, education, and history of illnesses (cardiovascular disease and cancer obtained through self-report or linkage with hospitalisation records, and cancer registries). Covariate harmonization procedures are described in **Supplemental Text 2**. Mortality data was linked with the NHS Digital of England and Wales, the NHS Central Register and National Records of Scotland, and the Taiwan National Death File registry. Primary cause of death was recorded using the International Classification of Diseases 10^th^ revision (ICD-10). CVD deaths included ICD-10 codes: I01 – I199. Cancer deaths included ICD-10 codes: C00 – C97.

### Statistical analysis

For our aggregate meta-analysis, we used random effect model to estimate summary measures (hazard ratios [HR] and 95%CI [confidence interval]) for the association of high vs low levels leisure-time physical activity with all-cause, CVD and cancer mortality.

For our pooled individual participant meta-analysis, we used Cox proportional hazards regression models to estimate HR with 95% CIs for all-cause mortality. For CVD and cancer analyses, we applied the Fine-Gray subdistribution method, treating non-cardiovascular/cancer deaths as a competing risk when appropriate. We calculated the dose-response association of MVPA duration (mins/week) as well as the duration of moderate intensity and vigorous intensity using restricted cubic splines with knots placed at the 10th, 50th, and 90th percentiles^22^. The reference value was set to no leisure time physical activity. To estimate the plausibility of bias from unmeasured confounding, we calculated E-values for all-cause mortality. To provide conservative point estimates for E-values, we assessed the minimum dose^11^, defined as the duration of physical activity associated with 50% of the lowest HR (‘optimal dose’; i.e., the nadir of the dose-response curve).

#### Sensitivity analyses

For sensitivity analyses of the aggregate meta-analysis, we performed leave-one-study-out analyses. We also conducted an analyses after excluding the unpublished individual participant data (IPD) estimates. For sensitivity analyses of the pooled individual participant meta-analysis, we included clinical factors available in each cohort that maybe mediators, including high density lipoprotein, low density lipoprotein, triglycerides, and diastolic and systolic blood pressure. We excluded participants with missing covariate data, and an event within the first 2 years of follow-up. Cause-specific analyses for CVD and cancer excluded participants with history of CVD or cancer, respectively.

We also performed sensitivity analyses to minimize the risk of reverse causation by excluding participants with self-rated fair or poor health. We included another analysis excluding participants with an event within the first 5 years of follow-up. We also included a sensitivity analysis excluding participants less than 40 years old are at a lower risk of CVD and cancer mortality during the follow-up time and may affect dose-response associations. We performed additional analysis using a leave-one-cohort-out approach, excluding one cohort at a time to ensure results were not due to one particular cohort with extreme results.

All statistical analyses were performed using R version 4.3.1. We reported this study as per the Preferred Reporting Items for Systematic Reviews and Meta-Analyses (PRISMA).

## Results

### Systematic Review

Searches retrieved a total of 14719 articles. After removing duplicates, the titles and abstracts of 10195 manuscripts were screened concerning the eligibility criteria, and 9918 were excluded. The full texts of the remaining 277 articles were evaluated and 252 were excluded for the following reasons: No adjustment for total physical activity, not primarily leisure-time physical activity, device-based physical activity only, no full text, wrong outcomes or patient population, no comparison of physical activity levels, physical activity not assessed using questionnaire, and wrong study design. The included 25 studies were carried out in different countries: Finland ^23^, United States ^24–32^, Taiwan ^33^, China ^34^ ^35^, England/Scotland/Wales ^36–39^, Denmark ^40^ ^41^, France/Germany/Italy/Netherlands/Greece ^42^, Sweden ^43^ ^44^, Australian ^45^, Spain ^46^, and Japan ^47^. A detailed definition of physical activity duration by type of activity and analytical categories across the 25 studies is available in **Supplemental Table 1**. Regarding the outcomes, 24 articles assessed all-cause mortality ^23–43^ ^45–47^, 18 cardiovascular mortality ^24–32^ ^34–37^ ^39^ ^41^ ^42^ ^44^ ^46^ and 11 cancer mortality ^26^ ^29–32^ ^34^ ^35^ ^37^ ^41^ ^42^ ^46^.

### Aggregate meta-analysis

Of these, 25 met the eligibility criteria for the systematic review and meta-analysis (**Figure 1**) totalling 3,355,076 participants and 247,463 all-cause, 70,204 CVD, and 76,294 cancer ^23–47^ deaths. Participant characteristics from 25 studies meeting the eligibility criteria of the systematic review are summarized in **Supplemental Table 1**.

**Figure 1.**
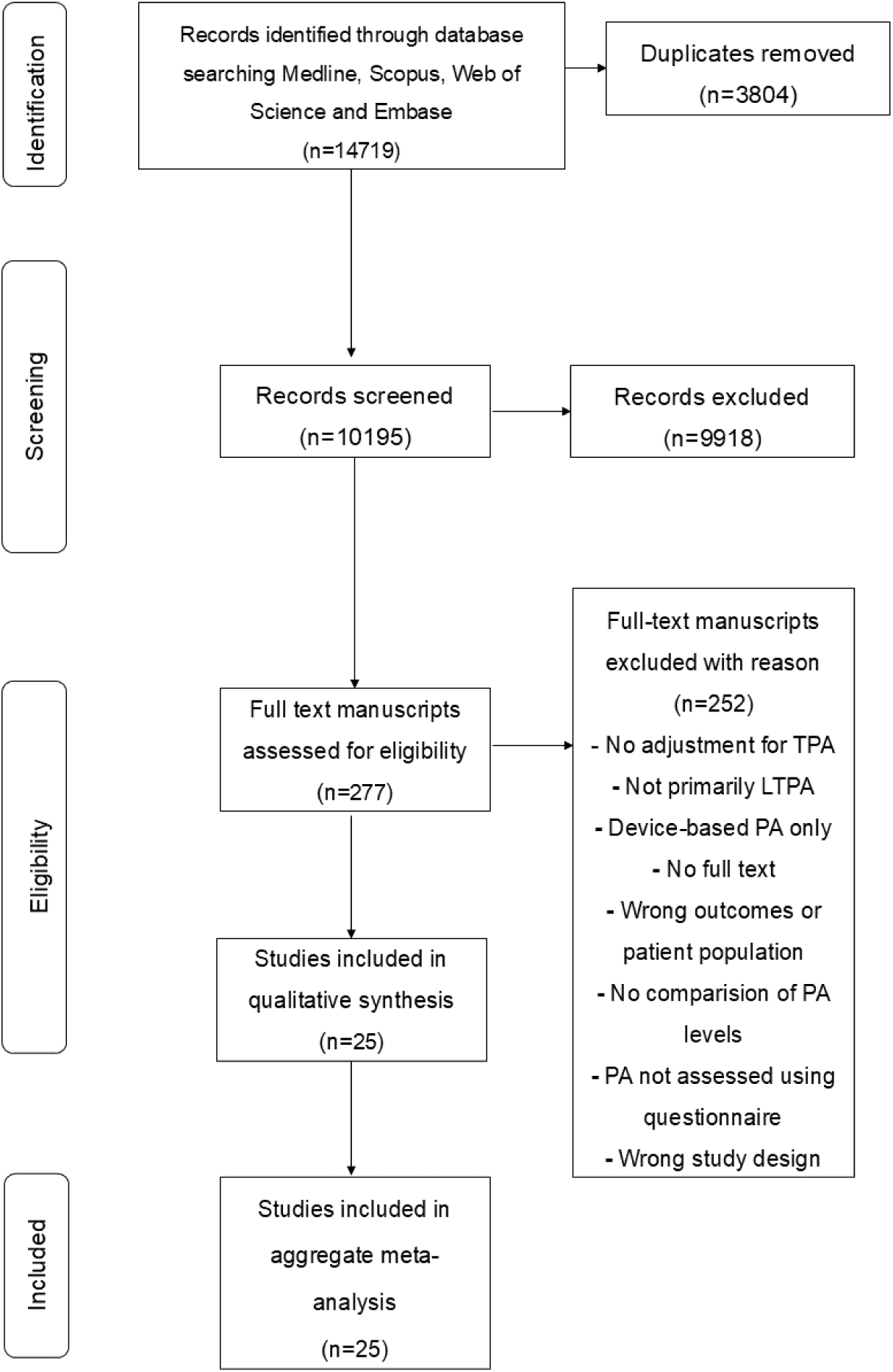
Preferred Reporting Items for Systematic Reviews and Meta-analysis (PRISMA) flow diagram for search strategy.

#### Moderate intensity leisure time-physical activity and mortality

Twenty studies ^23^ ^25–35^ ^37–40^ ^42^ ^45–47^ plus the IPD analysis were included in the meta-analysis for the association between high vs low moderate intensity leisure time-physical activity and all-cause mortality adjusting for total duration of physical activity. The summary HR for high vs low moderate intensity leisure time-physical and all-cause mortality was 0.84 (0.79-0.89) (**Figure 2A**).

**Figure 2.**
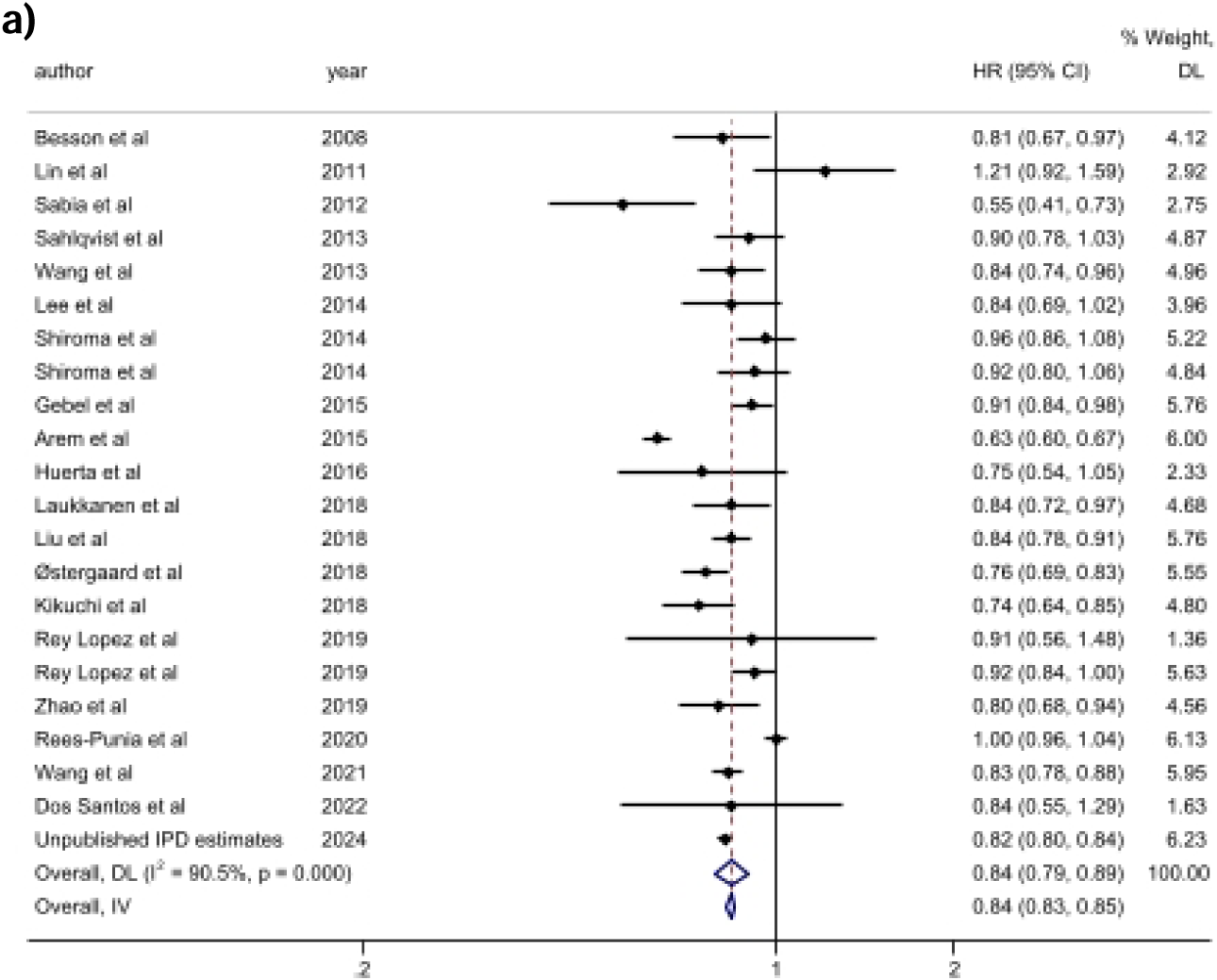
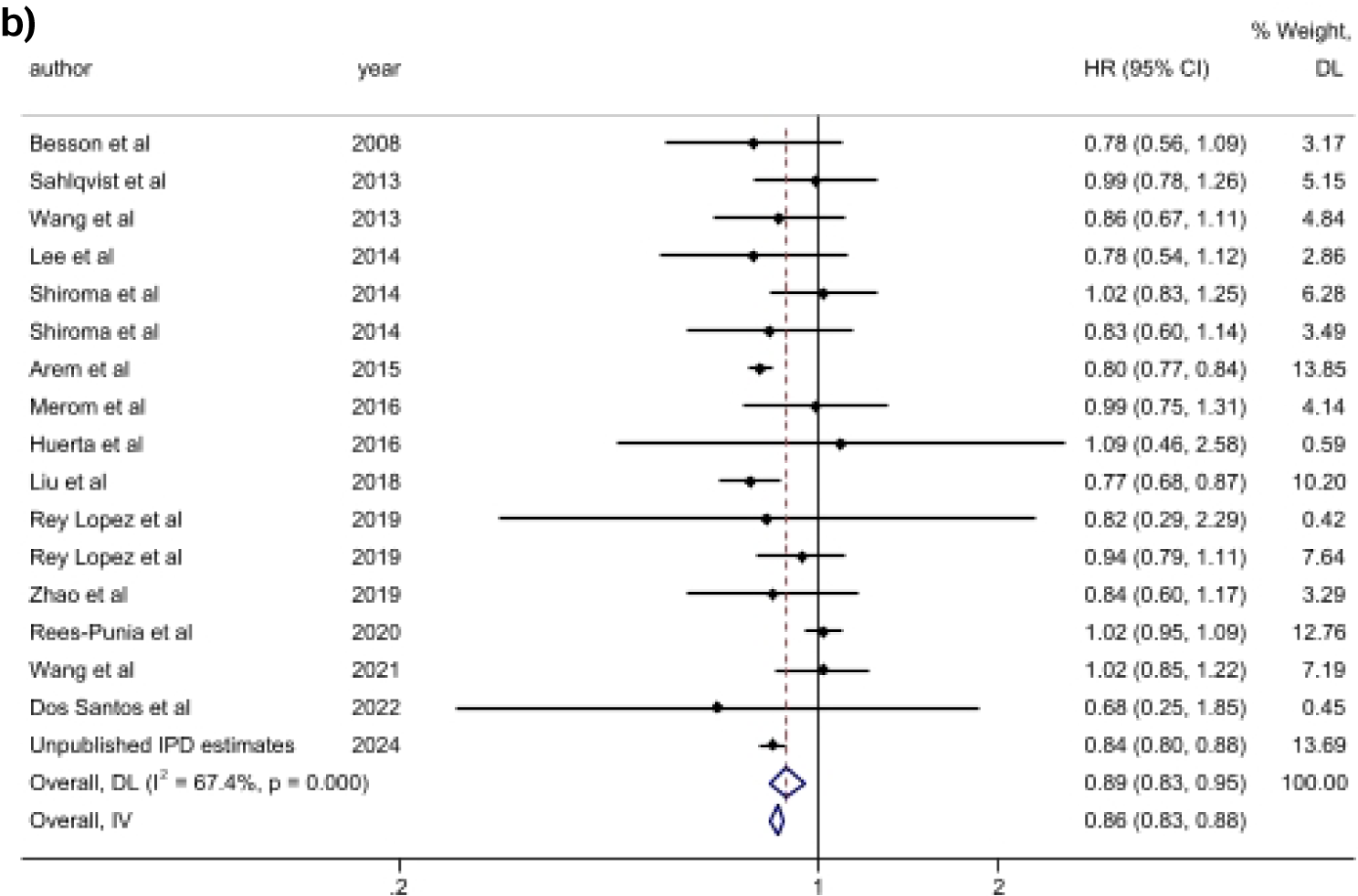
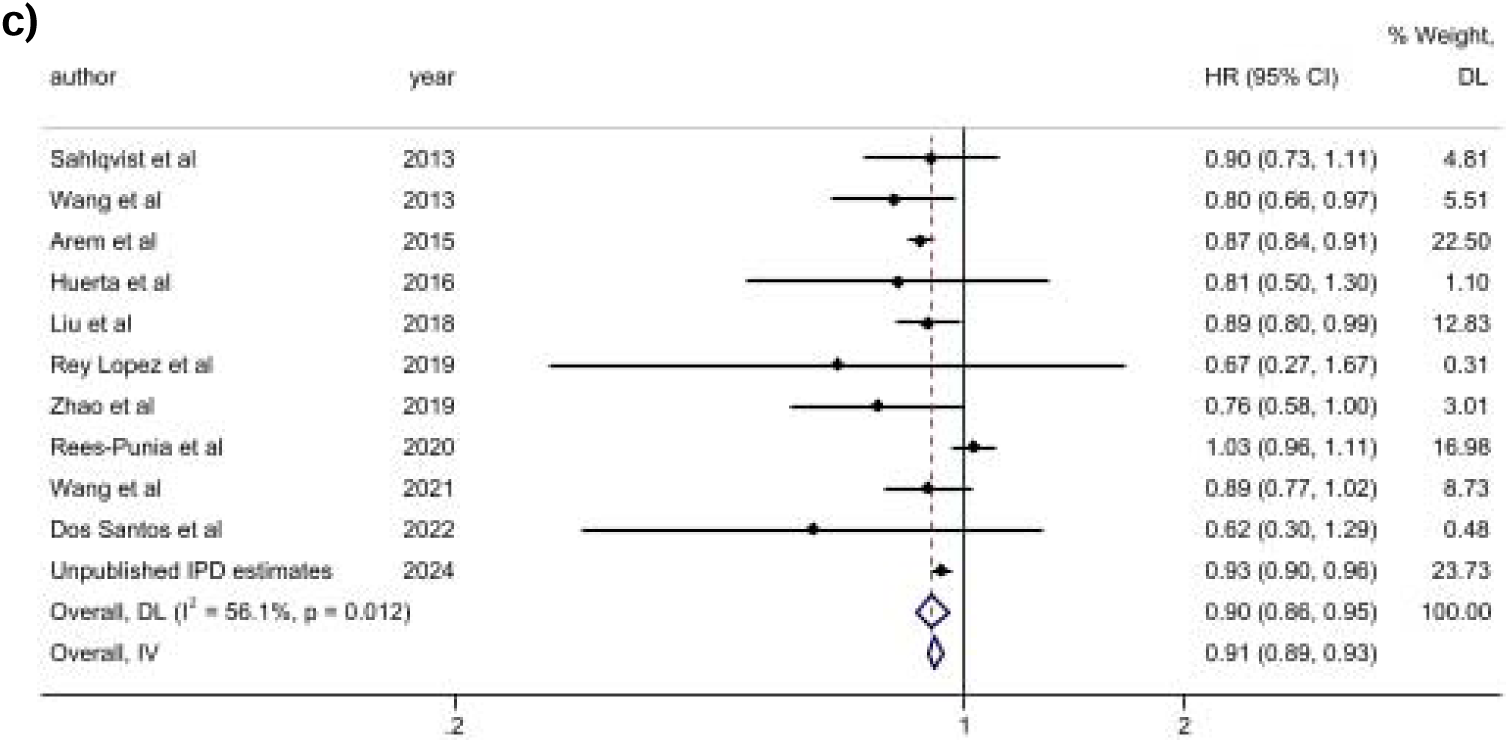
Meta-analysis of high vs low moderate intensity leisure time-physical activity and mortality: a) all-cause mortality; b) cardiovascular mortality; c) cancer mortality.

Fifteen ^25–32^ ^34–37^ ^39^ ^42^ ^46^ and ten ^26^ ^29–32^ ^34^ ^35^ ^37^ ^42^ ^46^ studies examined the association between high vs low moderate intensity leisure time-physical activity and cardiovascular and cancer mortality, respectively. Compared with participants who performed high vs low moderate intensity leisure time-physical activity, the association was similar between cardiovascular disease and cancer mortality at 0.89 (0.83-0.95) and 0.90 (0.86-0.93), respectively (**Figure 2B-C**).

#### Vigorous intensity leisure time-physical activity and mortality

Twenty studies ^23^ ^25–32^ ^34^ ^35^ ^37–42^ ^45–47^ plus the IPD analysis assessed the association of high vs low vigorous intensity leisure time-physical activity and all-cause mortality adjusting for total duration of physical activity. Compared with participants who performed low vigorous intensity leisure time-physical activity, the summary HR for all-cause mortality was 0.86 (0.79-0.93) (**Figure 3A**).

**Figure 3.**
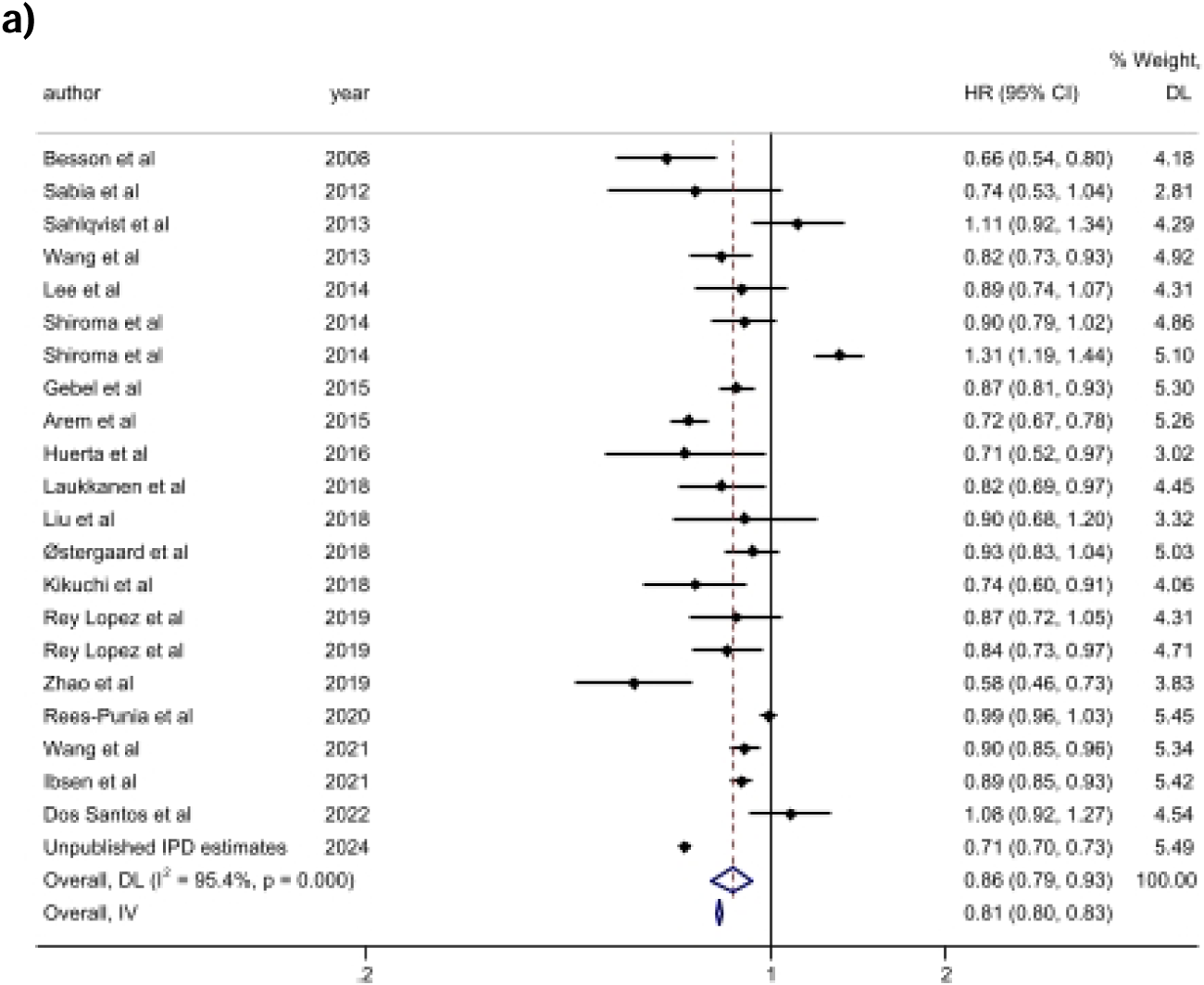
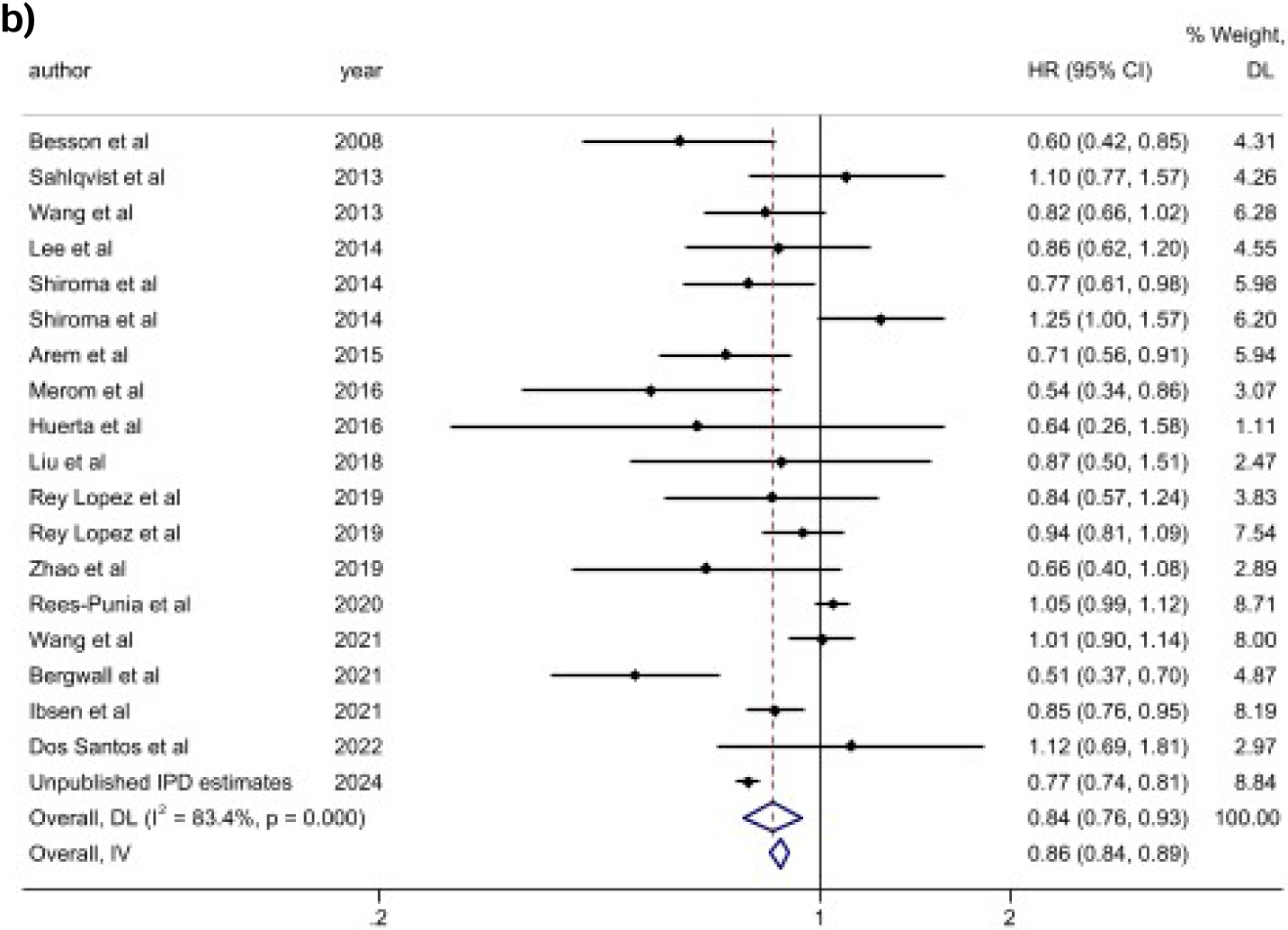
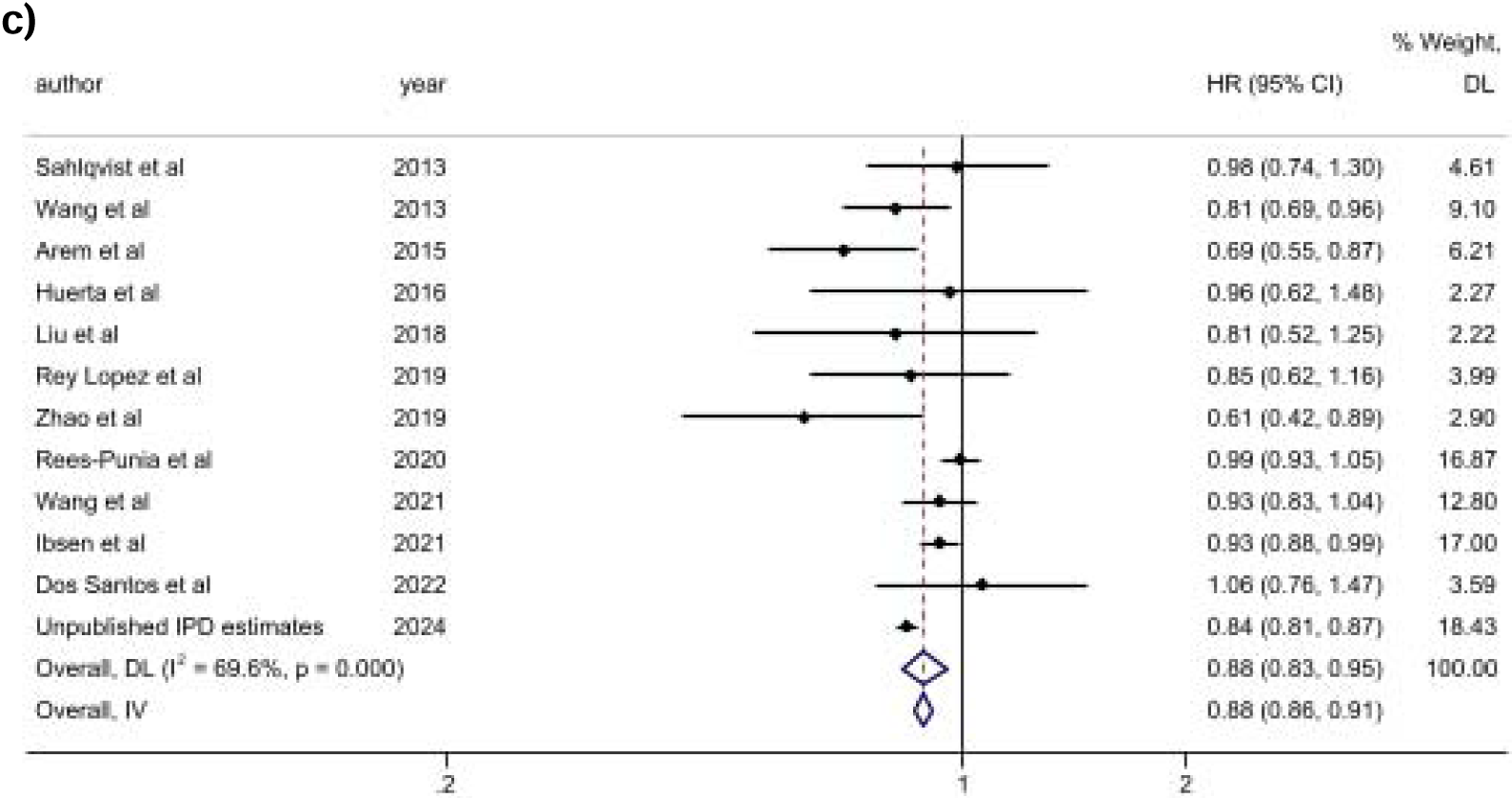
Meta-analysis of high vs low vigorous intensity of leisure time-physical activity and mortality: a) all-cause mortality; b) cardiovascular mortality; c) cancer mortality.

Seventeen ^25–32^ ^34–37^ ^39^ ^41^ ^42^ ^44^ ^46^ for cardiovascular disease and eleven ^26^ ^29–32^ ^34^ ^35^ ^37^ ^41^ ^42^ ^46^ studies for cancer, plus the IPD analysis, were included in the meta-analysis for the association between high vs low vigorous intensity leisure time-physical activity. The summary HR for the association between high vs low levels of vigorous intensity of leisure time-physical activity and cardiovascular disease and cancer mortality were 0.84 (0.76-0.93) and 0.88 (0.86-0.91) (**Figure 3B-C**).

#### Sensitivity analyses

Leave-one-out sensitivity analysis and exclusion of the unpublished IPD estimates showed consistent results with the main high vs low meta-analysis for vigorous and MVPA (**Supplemental Figures 2 and 3**).

### Pooled individual participant data meta-analysis

Our individual participant data meta-analysis included 967,184 participants with an average follow-up time of 12.2 (SD= 4.7) years, and 60,206 all-cause, 11,525 CVD, and 23,740 cancer deaths.

We observed an inverse association between leisure time physical activity duration with all-cause and cause-specific mortality (**Figure 4**). For each outcome, the dose-response curves were L-shaped, with the strength of the association plateauing or mildly inverting (MPA) at different weekly doses. For all-cause mortality (**Figure 4A**), the optimal dose (nadir of the curve) of MVPA was observed at 450 mins/week (HR [95%CI] = 0.67 [0.65, 0.69]). For moderate intensity and vigorous intensity, we observed the optimal dose at 340 mins/week (HR= 0.77 [0.75, 0.79]) and 200 mins/week (HR= 0.69 [0.67, 0.71]), respectively. We observed the minimal dose for MVPA at 115 mins/week (HR= 0.82 [0.80, 0.84]; E-value = 1.74 [1.67]). For moderate intensity and vigorous intensity, the minimal dose was observed at 100 mins/week (HR= 0.88 [0.86, 0.90]; E-value = 1.53 [1.46]) and 60 mins/week (HR=0.82 [0.80, 0.84]; E-value = 1.73 [1.67]).

**Figure 4:**
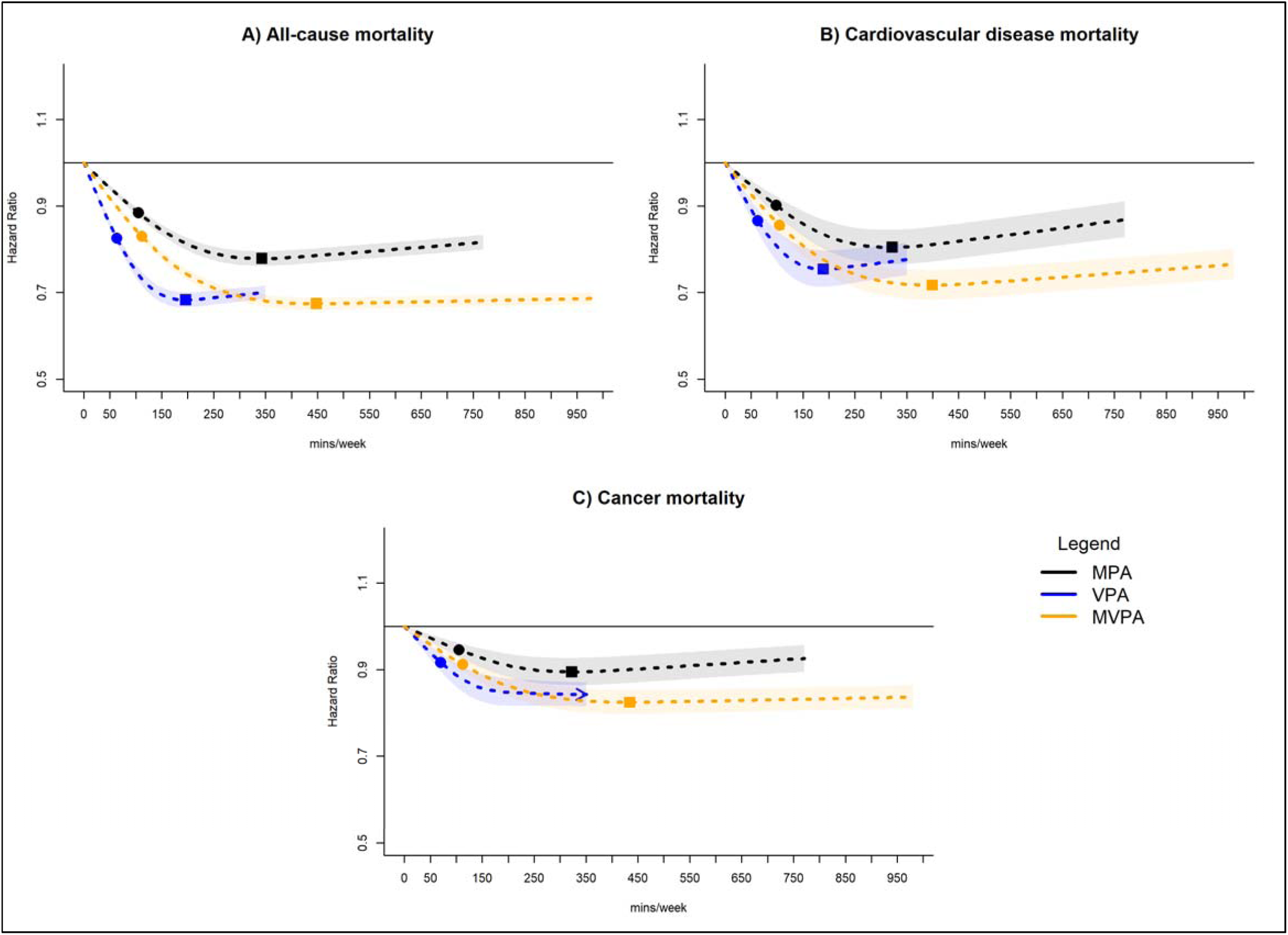
Dose-response association of leisure time physical activity intensity volume with all-cause, cardiovascular disease, and cancer mortality. Adjusted for age, sex, smoking status, alcohol consumption, body mass index, education, prevalent cvd (for all-cause and cancer mortality), prevalent cancer (for all-cause and cardiovascular disease mortality). Square = nadir of curve (optimal dose); circle = ED50 (minimal dose; 50% of optimal dose); arrow = no nadir of curve, magnitude of association increases with higher volume.

We observed a similar dose-response association for CVD mortality (**Figure 4B**), with optimal doses of 400 mins/week for MVPA (HR = 0.71 [0.68, 0.75]), 320 mins/week for moderate intensity (HR = 0.80 [0.76, 0.84]), and 190 mins/week for vigorous intensity (HR = 0.75 [0.71, 0.80]). The corresponding minimal doses were 100 mins/week for MVPA (HR = 0.86 [0.84, 0.88]; E-value = 1.60 [1.53]), 90 mins/week for moderate intensity (HR = 0.90 [0.88, 0.92]; E-value = 1.46 [1.39]), and 60 mins/week for vigorous intensity (HR = 0.86 [0.84, 0.89]; E-value = 1.60 [1.50]). For cancer mortality (**Figure 4C**) we observed an attenuated dose-response with the nadir of the curve at higher durations for each exposure. For example, the nadir for MVPA was at 430 mins/week, with a corresponding HR of 0.82 [0.80, 0.85], and for moderate intensity the nadir was at 322 mins/week (HR= 0.89 [0.86, 0.93]). For vigorous intensity, the dose-response began to plateau at about 175 mins/week (HR= 0.85 [0.82, 0.88]). The corresponding minimal doses were 110 mins/week for MVPA (HR= 0.91 [0.89, 0.93]; E-value= 1.42 [1.36]), 105 mins/week for moderate intensity (HR= 0.94 [0.92, 0.96; E-value= 1.32 [1.25]), and 70 mins/week for vigorous intensity (HR= 0.92 [0.89, 0.94]; E-value= 1.39 [1.32]).

#### Sensitivity analyses

Sensitivity analyses excluding the first 5 years of follow-up, participants with poor or fair self-rated health, participants under 40 years of age, and adjusted for biomarkers (high-and low-density lipoprotein, triglycerides, and blood pressure) showed consistent results with the main dose-response analyses, for vigorous and MVPA. For moderate intensity activity duration, associations became null for cancer mortality after exclusion of fair and poor self-rated health; however, dose-response results remained significant after excluding the first five years of follow up and adjustment for biomarkers. (**Supplemental Figures 4-7**). We found no appreciable differences in the dose-response association for moderate, vigorous, and moderate to vigorous intensity with each outcome (all-cause, cardiovascular, and cancer mortality) in the leave-one-cohort-out analyses (**Supplemental Figures 8-10**)

## Discussion

In our study, we systematically reviewed and meta-analysed the associations of leisure-time MVPA and its constituent components for weekly duration with all-cause, CVD, and cancer mortality. Pooling data from 3,355,076 participants across 25 cohorts, our findings provide additional insights into the differing contributory health-enhancing benefits of moderate and vigorous physical activity beyond traditionally assessed total duration of activity. Our pooled individual participant data meta-analysis from 964,923 participants extends on our aggregate meta-analysis to assess the intensity-specific dose-response relationship with all three outcomes. Our novel findings indicate a minimal leisure time physical activity duration of 115 mins/week for MVPA, 100 mins/week for moderate intensity, and 60 mins/week for vigorous intensity, with optimal durations of 450 mins/week, 340 mins/week, and 200 mins/week, respectively. For CVD and cancer mortality the minimal dose ranged from 100-175 mins/week of MVPA, 90-105 mins/week of moderate intensity, and 60-70 mins/week of vigorous intensity.

Our pooled meta-analysis of 25 cohorts showed higher durations of either moderate or vigorous intensity leisure-time physical activity were associated with lower all-cause, CVD, and cancer mortality risk. While previous meta-analyses have primarily focused on the combined effects of MVPA^6^ ^7^ ^48–50^, our analysis is among the first to separately examine the associations of moderate and vigorous intensities, encompassing 25 cohorts from 17 countries. We gave attention to the relative contributions of each intensity after adjustment for other activity levels that had not previously considered in other meta-analyses that reported contradictory results, suggesting that moderate and vigorous intensity reduced mortality risk to the same extent^51^. Extending on the aggregate meta-analysis, our individual participant data meta-analysis suggests the combined effects of leisure time MVPA may be as beneficial as vigorous activity alone, in lowering all-cause, CVD, and cancer mortality, although requiring a greater overall duration of activity – e.g. minimal dose for CVD mortality of 100 mins/week of MVPA had a similar magnitude of association as the minimal dose of 60 mins/week of vigorous activity. The similar mortality risks we observed could be attributable to the balance of physical stress induced by a mixture of moderate and vigorous activity, and the complementary physiological adaptations induced by moderate intensity and vigorous intensity together. In regards to cancer mortality risk, moderate intensity activity can lead to reduced inflammation and improved hormonal regulation, whereas vigorous intensity activity contributes to inhibiting carcinogen activation and enhanced detoxification^51^. Specifically for CVD mortality, moderate intensity activity may lead to reduced blood pressure, improved cholesterol levels, and enhanced autonomic tone. Conversely, vigorous intensity activity contributes to lower CVD risk partly through cardiac hypertrophy, increased blood volume, and overall cardiorespiratory fitness^52–54^. Our findings may inform future clinical trials and targeted patient treatment to mitigate disease risk through leveraging the unique physiological adaptations induced by different physical activity intensities.

In our pooled individual participant data meta-analysis dose-response, across each intensity exposure there was a consistent attenuated association for cancer mortality compared to CVD mortality. The attenuated dose-response association we observed is likely reflective of the diverse physiological effects of physical activity and differences in disease pathogenesis. Previous research has indicated the mechanistic link between physical activity and cancer risk is attributable to alterations in metabolic hormones and heightened anti-inflammatory responses^55^ ^56^. However, prior research has suggested this may be an indirect relationship and mediated by other factors such as body fat and dysregulation of cell apoptosis^56^ ^57^. Collectively, this suggests duration of leisure-time moderate and vigorous intensity activity might be more causally linked to CVD than to cancer. Notably, our dose-response results remained consistent after stringent sensitivity analyses that excluded the first five years of follow up and adjustment for biomarkers. Due to the diverse etiology of different cancer sites, it has been indicated that physical activity has a more direct effect on certain cancer sites than others^58–60^. The diversity in cancer etiology may have contributed to the attenuated associations we observed. Future research should target specific cancer sites that are directly linked to physical activity levels to optimise prevention strategies.

### Strengths and limitations

Our study has several strengths. This is, to our knowledge, the largest study conducted on intensity-specific leisure-time physical activity with all-cause, CVD, and cancer mortality. This afforded us the statistical precision to examine the minimum and optimum dose-response relationship across different intensities and mortalities. Our consistent methodological approach, including restriction to prospective cohort studies, and leisure-time physical activity, as well as analyzing the same physical activity level contrasts across studies. This approach minimizes heterogeneity, improves consistency of results, and maximizes power. We completed a systematic review of the literature for studies that assessed leisure time physical activity and intensity specific activities to pool 25 cohort studies that included more than 2.3 million participants. In addition, we provide a pooled individual participant data meta-analysis of new data from more than 967,000 participants. Our pooled individual participant data meta-analysis allowed us to examine the dose-response relationship between the duration of intensity-specific leisure-time physical activity and mortality risk. This provided more nuanced and generalizable information on the optimal duration of physical activity at different intensities with all-cause, CVD, and cancer mortality. To minimize bias from reverse causation, the first 5 years of follow-up and participants with poor or fair self-rated health were excluded in the sensitivity analyses. Despite these extensive precautionary measures, the potential for reverse causation from prodromal disease remains. Due to the observational design, the possibility of unmeasured confounding cannot be excluded. However, our E-values indicate an unmeasured confounder would need to have an association between 1.53 to 2.26 with the exposure and outcome for the association to be null. We used self-reported physical activity, which includes some recall error. Mitigating this is the structured nature of leisure-time physical activity makes it comparatively easy to recall^61^.

### Conclusions

Relatively modest amounts of leisure time moderate intensity or vigorous intensity, between 60 to 100 mins/week, may significantly lower the risk of all-cause, CVD, and cancer mortality. Engaging in a combination of moderate to vigorous leisure time physical activity may offer additional protective benefits against mortality compared to each intensity level individually. This could be due to the complementary health adaptations induced by each intensity level; however, vigorous activity may provide the strongest protective effect at a comparatively lower duration. Our findings suggest that promoting even small improvements in leisure-time physical activity could lead to substantial health benefits for adults, making it an effective strategy to mitigate mortality risk.

## Data Availability

All data generated as part of this systematic review are included as additional files in this published article.

