## Supplemental Materials for "Intensity-specific leisure time-physical activity and all-cause, cardiovascular, and cancer mortality in 3.36 million adults from 17 countries: a systematic review, meta-analysis, and individual participant data pooled analysis"

**SUPPLEMENTARY MATERIALS**

| **Page** | **Item** |
| --- | --- |
| **1** | **Supplemental Text 1: SEARCH STRATEGY** |
| **2** | **Supplemental Table 1: Main characteristics of the 25 studies included in the systematic review** |
| **16** | **Supplemental Text 2: Individual participant data meta-analysis harmonization procedures** |
| **23** | **Supplemental Table 2: Individual participant data meta-analysis baseline characteristics by cohort** |
| **26** | **Supplemental Figure 1: Funnel Plot** |
| **32** | **Supplemental Figure 2: Aggregated meta-analysis results after exclusion of unpublished IPD estimates** |
| **38** | **Supplemental Figure 3: Leave-one-out sensitivity analysis for the aggregated meta-analysis of high vs low leisure time physical activity by intensity and mortality outcomes** |
| **44** | **Supplemental Figure 4: Dose-response association of leisure time physical activity intensity volume with all-cause, cardiovascular disease, and cancer mortality; exclusion of participants with <5 years of follow-up** |
| **45** | **Supplemental Figure 5: Dose-response association of leisure time physical activity intensity volume with all-cause, cardiovascular disease, and cancer mortality; exclusion of participants with poor or fair self-rated health** |
| **46** | **Supplemental Figure 6: Dose-response association of leisure time physical activity intensity volume with all-cause, cardiovascular disease, and cancer mortality; exclusion of participants under 40 years of age** |
| **47** | **Supplemental Figure 7: Dose-response association of leisure time physical activity intensity volume with all-cause, cardiovascular disease, and cancer mortality; adjustment for biomarkers** |
| **48** | **Supplemental Figure 8: Leave-one-cohort-out dose response association with all-cause mortality** |
| **49** | **Supplemental Figure 9: Leave-one-cohort-out dose response association with cardiovascular disease mortality** |
| **50** | **Supplemental Figure 10: Leave-one-cohort-out dose response association with cancer mortality** |

**Supplemental Text 1: SEARCH STRATEGY**

**Pubmed/Medline – 05/15/2023**

(((((((((((((“leisure-time physical activity*”) OR (“recreational physical activity*”)) OR (“physical activity*”)) OR (“exercise*”)) OR (“physical exercise”)) OR (“physical fitness”)) OR (“walk*”)) OR (“jog*”)) OR ("run")) AND (((((((((((mortality) OR (all-cause mortality)) OR (cardiovascular mortality*)) OR (cancer mortality*)) Filters: Humans

**Web of Science - 05/15/2023**

(((((((((((((“leisure-time physical activity*”) OR (“recreational physical activity*”)) OR (“physical activity*”)) OR (“exercise*”)) OR (“physical exercise”)) OR (“physical fitness”)) OR (“walk*”)) OR (“jog*”)) OR ("run")) AND (((((((((((mortality) OR (all-cause mortality)) OR (cardiovascular mortality*)) OR (cancer mortality*))

**Scopus - 05/15/2023**

( TITLE-ABS-KEY ( leisure-time physical activity ) ) OR ( TITLE-ABS-KEY ( recreational physical activity ) ) OR ( TITLE-ABS-KEY ( physical activity* ) ) OR ( TITLE-ABS-KEY ( physical fitness* ) ) OR ( TITLE-ABS-KEY ( walk* ) ) OR ( TITLE-ABS-KEY ( jog* ) ) OR ( TITLE-ABS-KEY ( run* ) ) AND ( ALL ( " mortality*" ) ) OR ( TITLE-ABS-KEY ( "all-cause mortality*" ) ) OR ( TITLE-ABS-KEY ( "cardiovascular mortality*" ) ) OR ( TITLE-ABS-KEY ( "cancer mortality*" ) ) AND ( LIMIT-TO ( SRCTYPE , "j" ) ) AND ( LIMIT-TO ( DOCTYPE , "ar" ) ) AND ( LIMIT-TO ( EXACTKEYWORD , "Human" ) OR LIMIT-TO ( EXACTKEYWORD , "Humans" ) )

**Embase – 05/15/2023**

('leisure-time physical activity'/exp OR 'leisure-time physical activity' OR 'recreational physical activity'/exp OR ' recreational physical activity' OR 'physical activity'/exp OR 'physical activity' OR 'exercise'/exp OR 'exercise' OR 'physical exercise'/exp OR 'physical exercise' OR 'physical fitness'/exp OR 'physical fitness' OR 'walk'/exp OR 'walk' OR 'jog'/exp OR 'jog' OR 'run'/exp OR 'run') AND ('mortality '/exp OR 'mortality ' OR ' all-cause mortality'/exp OR 'all-cause mortality' OR 'cardiovascular mortality'/exp OR 'cardiovascular mortality' OR 'cancer mortality'/exp OR 'cancer mortality') AND [embase]/lim AND 'human'/de

**Supplemental Table 1**: **Main characteristics of the 25 published studies included in the systematic review**.

| **Reference** | **Country/ Region** | **Cohort sampling and representativeness** | **Sample size/deaths** | **Participants characteristics** | **Measurement of leisure-time physical activity** | **Summary statistics of LTPA** | **Adjustment for PA** | **Exclusion criteria and multivariable models** | **Outcomes** | **Main multivariable adjusted results** |
| --- | --- | --- | --- | --- | --- | --- | --- | --- | --- | --- |
| Laukkanen et al., [14] | Finland/ The city of Kuopio and surrounding rural communities in eastern Finland | This is a follow-up study of participants in the Kuopio Ischemic Heart Disease Study. | 2087 men/ 1028 (all-cause mortality was assessed during 26.1(18.7-28.0) years of follow-up [median(IQR)]. | Age at study entry was 53(5) years [mean(sd)]. | Type of measure: Cross-country skiing and total leisure-time physical activity in the past year were self-reported using a modified version of the Minnesota Leisure-Time Physical Activity Questionnaire. Cross-country skiing was assigned an intensity of 9.6 METS.  Type of LTPA: Total physical activity was classified as conditioning (e.g. walking, skiing and other aerobic activities) and non-conditioning (e.g. crafts, gardening). Categories of LTPA by volume: 0, 1-200, and >200 MET-hours per year.  Categories of LTPA by duration: 0, 1-60, or >60 minutes per week. | Total volume of cross-country skiing was the exposure in one analysis, defined as: 0, 1-200, and >200 MET-hours per year. Average duration of cross-country skiiing was the outcome in the other analysis, defined as: 0, 1-60, or >60 minutes per week. At baseline, total volume of skiing was 42.5 (0-195.5) MET-h/y,[mean(IQR)] and total duration of skiing was 60 (60-90) min/wk. | Associations between skiing and mortality were adjusted for total physical activity, which was expressed as kcal/d (i.e., kilocalories per day). | Participants were "apparently healthy middle-aged men" with "no missing data on… cross-country skiing".   Model 1: adjusted for age.  Model 2: adjusted for age, body mass index, systolic blood pressure, HDL-cholesterol, smoking status, alcohol consumption, prevalent coronary heart disease, history of diabetes mellitus, resting heart rate, and total physical activity. | All deaths. | All-cause mortality 1028 deaths HR (95% CI)  For total volume of cross-country skiing: 1.00 for 0 MET-h/y (reference); 0.84 (0.73, 0.97) for 1-200 MET-h/y; and 0.80 (0.67, 0.96) for >200 MET-h/y.  HR (95% CI)  For duration of cross-country skiing: 1.00 for 0 min/wk (reference); 0.84 (0.72, 0.97) for 1-60 min/wk; and 0.82 (0.69, 0.97) for >60 min/wk. |
| Lee et al., [15] | USA /  Texas | Participants self-referred to the clinic or were referred by their empolers or physicians. This is not a representative sample. | All cause mortality: 55137 adults / 3413 all-cause deaths during 14.7 years of follow-up (6.5-21.7) [mean (IQR)] CVD mortality: 52941 / 1217 CVD deaths during 14.6 (6.3-21.8) years of follow-up. | Participants were men and women aged 18 to 100 years. Mean age was 44 years. Ages were not provided for men and women separately. | Type of measure: At baseline, running or jogging in the last three months was assessed using a questionnaire that had four questions about duration, distance, frequency, and speed. The assessment of total physical activity is described in another paper, to which the reader is directed (Lee et al., Br J Sports Med, 2011, 45, 6, 504-510). In that paper, it says that total physical activity was assessed as self-reported leisure-time physical activity in the last three months. Participants were asked about ten activities and the total volume of physical activity was expressed as MET-minutes per week. Type of LTPA: running or jogging Categories of LTPA: non-runners and 5 quintiles of weekly running time (minutes), distance (miles), frequency (times), amount (MET-minutes), and speed (mph) in runners. | In the main analysis, the reference group was non-runners and the comparison groups were quintiles of running time per week. | In the main analysis, associations between running time and mortality were adjusted for "other physical activities except running", categorised as 0, 1-499, or >/= 500 MET-minutes/week (see Table 3 in manuscript). | Participants reporting myocardial infarction, stroke, or cancer at baseline were excluded. Participants with less than one year of follow-up were also excluded.   Main model: was adjusted for age, sex, examination year, smoking, alcohol consumption, other physical activities except running, BMI, presence or absence of abnormal ECG, hypertension, diabetes, and hypercholesterolaemia. | All-cause mortality, CVD Mortality. | All-cause mortality 3413 deaths  HR (95% CI) for all-cause mortality: 1.00 for non runners (reference); 0.80 (0.66-0.97) for quintile 1 of running time; 0.76 (0.63-0.91) for quintile 2; 0.78 (0.64-0.95) for quintile 3; 0.84 (0.69-1.02) for quintile 4; and 0.89 (0.74-1.07) for quintile 5.  CVD mortality  1217 deaths HR (95% CI) for CVD mortality: 1.00 for non runners (reference); 0.59 (0.40-0.86) for quintile 1 of running time; 0.67 (0.47-0.95) for quintile 2; 0.82 (0.58-1.16) for quintile 3; 0.78 (0.54-1.11) for quintile 4; and 0.86 (0.62-1.21) for quintile 5. |
| Lin et al., [16] | Taiwan / Tainan | The sampling scheme was a three stage process that generated a stratified cluster sample of eligible subjects throughout the city. | 876 participants / 312 deaths during during eight years of follow-up (1996-2004) Person-years of follow-up not reported. | Participants were community-dwelling men and women aged 65 years or older. Age was 72(6) years [mean(SD)]. | Type of measure: In the main analysis, participants were asked about the frequency and duration of 12 "common" leisure time physical activities in the previous two weeks.  Type of LTPA: 12 "common" leisure time physical activities in the previous two weeks: calisthenics or tai chi, gardening, walking for pleasure, bicycling, jogging, hiking, aerobic dancing, folk dancing, tennis, swimming, golf, and miscellaneous exercise. Non-LTPA was defined as housework and transportation and was assessed in a separate analysis. I have not included the results for non-LTPA because the figures don't tally.  Categories of LTPA: LTPA non-participants (MET-hour/ week = 0) and LTPA participants (MET-hour/week > 0). | Participants were divided in two groups: LTPA non-participants (MET-hour/week = 0) and LTPA participants (MET-hour/week > 0). | Leisure-time physical activity | People with dificulties in carrying out activities of daily living were excluded from the multivariate analyses.   Main multivariate model included: LTPA, non-LTPA, age, gender, level of education, habitual smoking and drinking, living status, BMI, cancer, stroke, heart disease, diabetes, liver disease, renal disease, pulmonary disease, hypertension, and osteoarthropathy. Height and weight were measured. All other variables were self-reported during an interview. | All-cause mortality. | All-cause mortality: 312 deaths HR (95% CI) for all-cause mortality: 1.00 for participants in LTPA (reference) and 1.21 (0.92, 1.59) for non-participants. |
| Liu et al., [18] | USA /  Texas | Participants self-referred to the clinic or were referred by their empolers or physicians. This is not a representative sample. | 12591 adults / 276 all-cause mortality deaths during an average of 10.5 years of follow-up. | The final sample included 12,591 participants (21% women) aged 18 to 89 years (mean age 47) at baseline. Participants were exclulded if they reported myocardial infarction, stroke, or cancer at baseline. | Type of measure: Resistance exercise using either free weights or weight training machines was evaluated by weekly frequency (times/week) and average exercise time (minutes) for each session. The total weekly amount of resistance exercise was calculated by multiplying weekly frequency with the average minutes per session. Type of LTPA: Resistance exercise Categories of LTPA: Resistance Exercise frequency of zero, one, two, three, and four or greater≥ times/week and four categories by total RE amount of 0, 1–59, 60–119, and ≥120 minutes/week | Participants were classified into five categories by Resistance Exercise G7E frequency of zero, one, two, three, and four or greater≥ times/week and four categories by total Resistance Exercise amount of 0, 1–59, 60–119, and ≥120 minutes/week for the main analyses. | Associations between resistance exercise and mortality were adjusted for meeting aerobic exercise guidelines, defined as average physical activity >/=500 MET-min/week or not. | Participants were exclulded if they reported myocardial infarction, stroke, or cancer at baseline.   The fully-adjusted model included: baseline examination year, age, sex, | All-cause mortality. | All-cause mortality: 276 deaths HR (95% CI) for weekly frequency of resistance exercise: 1.00 for 0 d/wk (reference); 0.65 (0.44–0.97) for 1 d/wk; 0.68 (0.46–1.01) for 2 d/wk; 0.67 for 3 d/wk (0.40–1.11); and 1.29 (0.75–2.20) for >/= 4 d/wk.   HR (95% CI) for weekly minutes of resistance exercise: 1.00 for 0 min/wk (reference); 0.64 (0.47–0.88) for 1-59 min/wk; 0.84 (0.53–1.34) for 60-119 min/wk; 1.03 (0.59–1.80) for >/=120 min/wk. |
| Merom et al., [19] | United Kingdom | Samples were drawn using multistage stratified probability design to give a representative sample of the two United Kingdom countries. | 48390 participants / 1714 CVD deaths. During 444,045 person-years of follow-up (mean: 9.7 years, SD: 4.39 years), | Participants were 48,390 adults aged at least 40 years, without doctor diagnosed CVD at baseline. | Type of measure: Physical activity in the last four weeks was assessed using and interviewer-administered questionnaire. Walking intensity was determined based on respondents’ single choice of the usual walking pace: slow, average, brisk, or fast. Moderate-intensity walking included “brisk” or “fast” pace and light-intensity included “slow" or “average” pace. The intensity of other PA, including dancing, was determined by a positive response to the question: Was the effort of [activity] usually enough to make you out of breath or sweaty? Type of LTPA: walking and other prompted recreational PA Categories of LTPA: None, Moderate-intensity and light-intensity | The proportion who reported meeting physical activity guidelines was 57.6% in dancers and 39.9 in non-dancers. | Associations between dancing and CVD mortality were adjusted for average weekly MET-hours of PA including MET-hours of walking and dancing. | Fully-adjusted model included: sex, age, BMI, longstanding illness, psychological distress/depression, alcohol drinking frequency, cigarette smoking, occupational social class, and total MET-hours of physical activity. | CVD mortality | CVD mortality: 1714 deaths HR (95% CI) for dancing: 1.00 for none (reference); 0.99 (0.75, 1.32) for light intensity dancing; 0.54 (0.34, 0.87) for moderate intensity dancing.  HR (95% CI) for walking: 1.00 for none (reference); 0.91 (0.77, 1.07) for light intensity walking; 0.67 (0.52, 0.87) for moderate intensity walking. |
| Østergaard et al., [20] | Denmark / Aarhus and Copenhagen | Participants were identified through the Civil Registration System and 27190 men and 2863 women, 35% of all invited, agreed to participate in the baseline examination. | 28204 adults / 2942 deaths during the five years between baseline and follow-up examinations. | Participants were men and women aged 50 to 64 years at baseline. | Type of measure: Self-reported weekly level of cycling in leisure-time was assessed during baseline and follow-up examinations. Leisure time physical activity was handled as a composite measure of MET hours per week. Type of LTPA: Physical activity at baseline was reported in relation to work, walking, cycling, housework, do-it-yourself work, gardening, sports participation, and stair climbing for both summer and winter. Categories of LTPA: No cycling; Stop cycling; Initiate cycling; Consistent cycling; No cycling or stop cycling; Initiate or consistent cycling | Total cycling was 40 (0, 150) minutes/week [median (25th, 75th percentile)]. LTPA other than cycling was 65.5 (42.0, 98.9) Met hours/week. | Multivariable analyses of recreational cycling were adjusted for leisure time physical activity other than recreational cycling. | The multivariate model included: age, sex, years of basic school, higher education, physical activity at work, leisure time physical activity (MET hours per week excluding cycling), smoking, alcohol, monounsaturated fat, polyunsaturated fat, saturated fat, and coffee consumption. | All-cause mortality. | All-cause Mortality:  2942 deaths HR (95% CI) for cycling: 1.00 for no cycling at baseline or follow-up examinations (reference); 0.98 (0.87, 1.11) for those who stopped cycling; 0.78 (0.67, 0.90) for those who began cycling; and 0.77 (0.71, 0.84) for those who consistently cycled. |
| Rees-Punia et al., [22] | US / All 50 states, District of Columbia, and Puerto Rico | This subset of the Cancer Prevention Study included participants ranging in age from 50 to 74 years, who resided in 21 states with population-based state tumor registries, and were randomly invited to join the subset. | 123232 men and women / 46829 deaths during follow-up (1993-2014) | Participants were 50 to 74 years and resided in states with population-based state tumor registries. | Type of measure and type of leisure time: Light-intensity lesiure time physical activity was assessed with the question: "During the past year, what was the average time per week you spent on the following kinds of activities: ‘gardening, mowing, planting, etc.,’ ‘heavy housework, vacuuming, etc.,’ ‘heavy home repair, painting, etc.,’ and ‘shopping." Corresponding MET values (2.5 METs for heavy housework and shopping, 3.0 for gardening and heavy home repair) were multiplied by the lower limit of each category for hours per week (1, 4, or 7 h/week) and summed for an estimate of LPA MET-h per week. Survey MVPA items included “walking, jogging/running, lap swimming, tennis or racquetball, bicycling or stationary biking, aerobics/calisthenics, dancing,” with responses of: “none,” “1–3 h/week,” “4–6 h/week,” or “7+ h/week.” Categories of LTPA: LPA MET-h per week were categorized into approximate quintiles (0 to < 3, 3 to < 9, 9 to < 15, 15 to < 21, 21+) MVPA was categorized as “none,” < 8.75, 8.75 to < 17.5, or 17.5+ MET-h/week. | In general, women reported more leisure time LPA than men (median LPA: 10 MET-h/week for women vs. 8.5 MET-h/week men) and older adults reported more LPA than adults under the age of 70 years (median LPA: 11 MET-h/week for ≥70 years vs. 8.5 METh/ week for < 70 years). | Associations between light-intensity LTPA and mortality were adjusted for MVPA. | The authors excluded participants with a history of cancer, heart attack, stroke, or lung disease. Participants with missing data were also excluded. Participants who died during the first year of follow-up and those who reported no physical activity were excluded.   Model adjusted for age, sex, race/ethnicity, smoking, alcohol use, aspirin use, education, employment status, marital status, ACS diet score, comorbidity score, BMI,and MVPA. | All-cause mortality. Cancer mortality. CVD mortality. Respiratory mortality. | HR for light-intensity physical activity were reported in Supplemental Table 1.  All-cause mortality: HR (95% CI) for all-cause mortality: 1.00 for 3 to <9 MET-hours/week (reference) and 1.16 (1.12-1.20) for 0 to <3 MET-hours/week.  Cancer mortality: HR (95% CI): 1.00 for 3 to <9 MET-hours/week (reference) and 1.04 (0.98-1.11) for 0 to <3 MET-hours/week.  CVD mortality: HR (95% CI): 1.00 for 3 to <9 MET-hours/week (reference) and 1.24 (1.17-1.31) for 0 to <3 MET-hours/week.  Respiratory mortality: HR (95% CI): 1.00 for 3 to <9 MET-hours/week (reference) and 1.17 (1.04-1.31). |
| Rey Lopez et al., [23] | England and Scottish | The Health Survey for England (HSE) and the Scottish Health Survey (SHeS) are household-based prospective studies in which households were selected using a multistage, stratified probability design to achieve representative samples of the population of England and Scotland. The present study included 11 population cohorts of individuals aged 30 years or older: HSE 1994, 1997, 1998, 1999, 2003, 2004, 2006 and 2008; SHeS 1995, 1998 and 2003. | 64913 / 5064 deaths from all-causes, 1393 from CVD and 1602 from cancer during 435 743 person-years of follow-up | Adults aged ≥30 years. In overall, 56% were women, 59.5% were overweight or obese, 75.8% were non-current smokers, 80% did not drink alcohol frequently (less than five times per week) and 84.7% did not report psychological distress | Type of measure: Self-reported: Type of LTPA: Participants were asked about the frequency, duration and pace of walking (slow, average or brisk) and domestic physical activity. data on physical activity in sports and exercise were collected (frequency, duration and perceived intensity over the last 4 weeks) Categories of PA by intensity:  Proportion of each intensity (moderate and vigorous) to vigorous activity classified as vigorous (o%, 0.1-30%, and >30.1%) Categories of LTPA by frequency:  (1) 0%; (2)>0% to <30%; and (3) ≥30% of MVPA by vigorous activity. | More than half (58.3%) of the participants met the WHO’s PA recommendations 7 of 150 min of weighted MVPA per week. Of those who engaged in any MVPA, 68.6% did not report any vigorous activity, 7.7% reported less than 30% and 23.7% reported 30% or more of their total MVPA as vigorous activity. The highest total volume of physical activity was in the 0%–30% vigorous group, who reported a median of 461 min per week of MVPA | For all sample values were adjusted for alcohol consumption, total weighted volume of moderate-to-vigorous physical activity (MVPA), longstanding illness and CVD diagnosis at baseline | All individuals that reported no MVPA and/or did not provide information for any of the covariates included in the statistical models. Cox proportional hazards regression models were used. For the outcome CVD mortality, models were additionally adjusted for CVD diagnosis at baseline. For cancer mortality, models were additionally adjusted for cancer diagnosis at baseline. For all sample values were adjusted for alcohol consumption, total weighted volume of moderate-to-vigorous physical activity (MVPA), longstanding illness and CVD diagnosis at baseline. | All-cause mortality. CVD mortality. cancer mortality. | All-cause mortality:  Proportion of moderate to vigorous activity classified as vigorous  0% of recreational PA: 44521/4477 N/events >0%-<30% of recreational PA: 5000/151 N/events ≥30% of recreational PA: 15392/436 N/events  Cardiovascular mortality All-cause mortality:  Proportion of moderate to vigorous activity classified as vigorous  0% of recreational PA: 39913/1252 N/events >0%-<30% of recreational PA: 4316/35 N/events ≥30% of recreational PA: 13473/106 N/events Cancer mortality All-cause mortality:  Proportion of moderate to vigorous activity classified as vigorous  0% of recreational PA: 43576/1384 N/events >0%-<30% of recreational PA: 4936/53 N/events ≥30% of recreational PA: 15165/165 N/events All-cause mortality:  0% of recreational PA: reference >0%-<30% of recreational PA: HR and 95% CI: 0.84 (0.71, 0.99) ≥30% of recreational PA: HR and 95% CI: 0.84 (0.70, 1.01)  Cardiovascular mortality HR and 95% CI 0% of recreational PA (reference) >0%-<30% of recreational PA: 0.83 (0.57, 1.18) ≥30% of recreational PA: 0.84 (0.68, 1.04)  Cancer mortality HR and 95% CI: 0% of recreational PA (reference) >0%-<30% of recreational PA: 0.79 (0.60, 1.06) ≥30% of recreational PA: 0.89 (0.75 1.06) |
| Rey-Lopez et al., [24] | USA / Cambridge, Massachusetts | The Harvard Alumni Health Study is a prospective cohort study of men who matriculated as undergraduates at Harvard University between 1916 and 1950.  Men who returned questionnaires in 1988. Baseline characteristics of the men (median age [interquartile range]=65 years [60–71 years]). | 8874 / 4063 men died (1195 from CVD) during a median follow-up of 12.4 years | Mean age 65 (60-71) years. 8.2% current smokers, 27.$% with hipertension, 4.6% with diabetes, and 18.7% with high cholesterol. | Type of measure: Self-reported: Type of LTPA: walking, stair climbing, and sports or recreational activities during the past year LTPA exposure: frequency and duration of each of these activities Categories of LTPA by intensity:  <10 floors/wk, 10-19 floors/wk, 20-34 floors/wk, ≥35 floors/wk | Cox proportional hazards regression models Categories of number of floors climbed (0–9 floors/ wk; 10–19 floors/wk; 20–34 floors/wk; ≥35 floors/wk) | Model 1 adjusted for age, number of blocks walked, and energy expended on sports/recreation. Model 2 included all Model 1 covariates plus smoking, and alcohol intake.  Model 3 included all Model 2 covariates plus BMI, and diagnoses of hypertension, diabetes, or high cholesterol. | Excluded 3487 men with a history of CVD or cancer and 444 men with missing physical activity data, leaving 8874 participants in the present study. Model 1 adjusted for age, number of blocks walked, and energy expended on sports/recreation. Model 2 included all Model 1 covariates plus smoking, and alcohol intake.  Model 3 included all Model 2 covariates plus BMI, and diagnoses of hypertension, diabetes, or high cholesterol. | All-cause mortality. CVD mortality. | Compared with men who reported climbing fewer than 10 floors/ wk, a larger number of stairs climbed was not significantly associated with a lower risk for CVD mortality in any of the models.  For example, (Model 1): [HR (95% CI)=0.94 (0.79–1.11)], [HR (95% CI)=0.90 (0.76–1.08)], and [HR (95% CI)=0.94 (0.81–1.09)] for 10–19 floors/wk, 20–34 floors/wk, and ≥35 floors/wk. |
| Sabia et al., [25] | England / London | Whitehall II Study, which was established in 1985 as a longitudinal study on 10308 civil servants. All civil servants aged 35 to 55 years in 20 London based departments were invited to participate by letter, and 73% agreed. The baseline examination (phase 1) took place from 1985 to 1988 and involved a clinical examination and a self-administered questionnaire that included sections on lifestyle factors. Subsequent phases of data collection alternated between postal questionnaire alone (phases 2 [1988-1990], 4 [1995-1996], 6 [2001], and 8 [2006]) and postal questionnaire accompanied by a clinical examination (phases 3 [1991-1993], 5 [1997- 1999], and 7 [2002-2004]). | 7456 participants / 317 died during the mean follow-up of 9.6 years (SD = 2.7) | A total of 317 participants died during the mean follow-up of 9.6 years (SD=2.7). Mean age 55.2 [SD: 5.9], 55.4 [SD: 5.9], and 57.0 [SD: 6.1] years for ≤8 h/wk, 8.1-12 h/wk, and >12 h/wk of total physical activity | Type of measure: Self-reported: Type of PA: Leisure-time and job-related activities. questionnaire included 20 items on the amount of time spent in the following activities: walking, sports (cycling, soccer, golf, swimming, and 2 open-ended questions on other sports), gardening (weeding, mowing, and 1 open-ended question on other gardening activities), housework carrying heavy shopping, cooking, hanging out washing, and 2 open-ended questions on other housework), do-it-yourself activity (a term used to  describe building, modifying, or repairing something without the aid of experts or professionals, such as manual car washing, painting, or decorating  LTPA exposure: frequency sports (LTPA) Categories of sports (LTPA) by intensity:  None, 0,1-1,9 h/wk, ≥2 h/wk | N participants: 2415 (15%), 2361 (31.7%) and 2680 (35.9%) for ≤8 h/wk, 8.1-12 h/wk, and >12 h/wk of total physical activity | Model 1 was the unadjusted model using age as the timescale. Model 2 was the mutually adjusted model, in which the other measures of intensity (or type) of physical activity were simultaneously entered in the regression to evaluate their independent effects on mortality. Model 3 additionally adjusted for sociodemographic variables (gender, marital status, and socioeconomic status).  Model 4 included other health behaviors (smoking, alcohol consumption, and fruit and vegetable consumption) and health measures (coronary heart disease, stroke, diabetes, and self-rated health). | The proportional hazards assumption for the Cox model was confirmed formally by the Shoenfeld’s test.Tests for linear trend across the categories of duration of physical activity were obtained by entering the categorical variables as a continuous parameter in the Cox model. | Mortality (no additional information). | Mortality: 215 none sports; 42 for 0.1-1.9 h/wk, and 60 for ≥2 h/wk HR and 95% CI (Model 4) for 0.1-1.9 h/wk of sports : HR: 0.67 (95%CI: 0.48, 0.94), and for ≥2 h/wk of sports: HR: 0.73 (95%CI: 0.55, 0.99) |
| Sahlqvist et al., [26] | Europe | EPIC-Norfolk cohort, part of the 10-country collaborative EPIC. Between 1993 and 1997, 25633 adults aged 40–79 years were recruited from general practices in the county of Norfolk in the east of England and attended a health examination. | 22450 participants / 4398 (20%) participants died during 3 425 498 person-years of follow-up. There were 1379 (6.1%) cardiovascular deaths and 1639 (7.3%) cancer deaths | At the first health assessment, participants had a mean age of 58 years (SD=19) and just over half were women (55%). Twenty four per cent of the participants reported cycling for a mean of 165 min/week (SD=246). By the second health assessment, participants had a mean age of 62 (SD 9) years; just over half were women (57%). Thirty per cent (n=4030) reported any cycling. Of those who cycled, 62% (n=2808) reported cycling for recreation and 72% (n=3269) reported cycling for utility purposes with 26% (n=862) of these reporting commuting cycling. The average cyclist spent 83 min/week cycling. | Type of measure: Self-reported: Type of PA: Physical activity, separately for winter and summer, the weekly time (in hours) spent walking and cycling (separately) to work and during leisure, and in other exercise. Total cycling was calculated as the average weekly time spent in winter and summer (min/week) LTPA exposure: frequency sports (LTPA) Categories of sports (LTPA) by intensity:  None, 0,1-1,9 h/wk, ≥2 h/wk | Recreational PA  0 min/week 1-59 min/week | Model 1 was adjusted for sex, age, education level and social class Model 2 was adjusted for smoking status, family history of cardiovascular disease or cancer, as well as time spent walking and in other exercise ( all other physical activity energy expenditure) | Excluded those with cardiovascular disease (n=1102) or cancer (n=1327) and those with missing data (n=784) leaving 22 450 for analysis. Similarly, of those who returned for the second health assessment, we excluded those with self-reported cardiovascular disease (n=772) or cancer (n=1115) and those with missing data (n=286), leaving 13 346 for analysis. All participants were followed up for mortality to 31 March 2011 (mean 15.3 years (SD=3.3) from first health assessment, mean 11.5 years (SD=2.0) from second health assessment).  Model 1 was adjusted for sex, age, education level and social class Model 2 was adjusted for smoking status, family history of cardiovascular disease or cancer, as well as time spent walking and in other exercise. | All-cause. CVD mortality. Cancer mortality. | All-cause mortality:  0 min/week of recreational PA: 1483 death  1-59 min/week of recreational PA: 104 deaths Cardiovascular mortality 0 min/week of recreational PA: 438 death  1-59 min/week of recreational PA: 25 deaths  Cancer mortality 0 min/week of recreational PA: 608 death 1-59 min/week of recreational PA: 56 deaths All-cause mortality:  HR and 95% CI (Model 2):  0 min/week (reference)  1–59 min/week: 0.87 (0.69, 1.04)  Cardiovascular mortality HR and 95% CI (Model 2):  0 min/week (reference)  1–59 min/week: 0.75 (0.50, 1.13)  Cancer mortality HR and 95% CI (Model 2):  0 min/week (reference)  1–59 min/week: 0.95 (0.71, 1.25) |
| Shiroma et al., [27] | USA / Cambridge, Massachusetts | Harvard Alumni Health Study (HAHS) and women in the Women’s Health Study (WHS). The HAHS is a prospective cohort study of men matriculating at Harvard University between 1916 and 1950. | 7979 men and 38 671 women / There were 3551 deaths (1077 from CV disease) among men and 3170 deaths (620 from CV disease) among women Average follow-up time varied from 6.52 years to 17.8 years. | This resulted in a final baseline population of 7979 men. Baseline characteristics of the men (mean age=66.1 years [SD=7.7 years]) and women (mean age=54.6 years [SD=7.0 years]) by total physical activity volume.Of the 7979 men at baseline (year 1988), 3551 died (1077 of CVD) during 17.3 mean years of follow-up. Of the 38 761 women at baseline (year 1992), 3170 died (620 of CVD) during 16.4 mean years of follow-up. | Type of measure: Self-reported: Type of PA: number of flights of stairs climbed daily and the number of blocks walked daily and listed the frequency and duration spent doing sports or recreational activities PA exposure: MET-h/week Categories of PA by intensity:  (0 to <3.75 MET-h/week); (2) “some activity” (3.75 to <7.5 MET-h/week); (3) satisfying the guideline (7.5 to <15 MET-h/week); (4) 2 to <4 times the guideline (15 to <30 MET-h/week); and (5) at least 4 times the guideline (≥30 MET-h/week). | Men expended a median of 18.6 MET-h/week (25th percentile:6.3; 75th percentile: 37.4) in physical activity; the median for women was 8.4 MET-h/week (25th percentile: 2.7; 75th percentile: 20.5). | Model 1 adjusted for age and total MVPA per week;   Model 2 for age plus smoking status; alcohol consumption; vegetable and fruit intake; saturated fat intake; total caloric intake; and, in women only, fiber intake, randomization arm of the clinical trial period, parental history of myocardial infarction, postmenopausal status, and hormone therapy;   Model 3 adjusted for body mass index (kg/m2), high cholesterol, and hypertension | Men were excluded if they had missing information on physical activity at baseline (n=591) or a history of self-reported physician-diagnosed cardiovascular disease (CVD), cancer, or diabetes before 1988 (n=4235). | All-cause mortality. CVD mortality. | All-cause mortality:  Men 0 to <3.75 MET-h/week: cases, no (person-years): 757 (20625) 3.75 to <7.5 MET-h/week: cases, no (person-years): 392 (14244) 7.5 to <15 MET-h/week: cases, no (person-years): 542(21833)  15 to <30 MET-h/week: cases, no (person-years): 1170 (34632) ≥30 MET-h/week: cases, no (person-years): 1114 (46888)  Women 0 to <3.75 MET-h/week: cases, no (person-years): 1141 (194422) 3.75 to <7.5 MET-h/week: cases, no (person-years): 456 (94749) 7.5 to <15 MET-h/week: cases, no (person-years): 533 (125337)  15 to <30 MET-h/week: cases, no (person-years): 653 (136513) ≥30 MET-h/week: cases, no (person-years): 387 (84055)  Cardiovascular mortality:  Men  0 to <3.75 MET-h/week: cases, no (person-years): 233 (20625) 3.75 to <7.5 MET-h/week: cases, no (person-years): 116 (14244) 7.5 to <15 MET-h/week: cases, no (person-years): 150 (21833)  15 to <30 MET-h/week: cases, no (person-years): 223 (34632) Women 0 to <3.75 MET-h/week: cases, no (person-years): 233 (194749) 3.75 to <7.5 MET-h/week: cases, no (person-years): 77 (94749) 7.5 to <15 MET-h/week: cases, no (person-years): 90 (125337)  15 to <30 MET-h/week: cases, no (person-years): 147 (136513) ≥30 MET-h/week: cases, no (person-years): 73 (84055)  All-cause mortality:  Men HR and 95% CI: 0 to <3.75 MET-h/week (Reference) 3.75 to <7.5 MET-h/week: 0.79 (0.68, 0.92) 7.5 to <15 MET-h/week: 0.71 (0.62, 0.82) 15 to <30 MET-h/week: 0.68 (0.59, 0.77) ≥30 MET-h/week: HR and 95% CI: 0.63 (0.55, 0.71) Women HR and 95% CI: 0 to <3.75 MET-h/week (Reference) 3.75 to <7.5 MET-h/week: 0.72 (0.64, 0.81) 7.5 to <15 MET-h/week: 0.59 (0.53, 0.66) 15 to <30 MET-h/week: 0.49 (0.44, 0.54) ≥30 MET-h/week: 0.41 (0.36, 0.47) Cardiovascular mortality:  Men HR and 95% CI: 0 to <3.75 MET-h/week: (Reference) 3.75 to <7.5 MET-h/week: 0.69 (0.51, 0.93) 7.5 to <15 MET-h/week: 0.67 (0.52, 0.88) 15 to <30 MET-h/week: 0.69 (0.55, 0.87) ≥30 MET-h/week: 0.66 (0.53, 0.83) Women HR and 95% CI: 0 to <3.75 MET-h/week: (Reference) 3.75 to <7.5 MET-h/week: 0.74 (0.57, 0.97) 7.5 to <15 MET-h/week: 0.78 (0.62, 0.99) 15 to <30 MET-h/week: 0.58 (0.45, 0.74) ≥30 MET-h/week: 0.64 (0.49, 0.85) |
| Sundquist et al., [28] | Sweden | Simple, random sample from the Swedish Annual Level-of-Living Survey (SALLS), a national survey conducted by Statistics Sweden of the non-institutionalized population was used in this study. The sample consisted of 1792 women and 1414 men, aged 65, surveyed in 1988 and 1989. The sample represented 792000 women and 616000 men of the Swedish population aged 65. | 3206 / 925 deaths among women and 881 deaths among men (mean follow-up time was 11.7 years) | For both men and women, the highest mortality rates were observed among those who reported low educational status, physical inactivity, current smoking, underweight, obesity, diabetes, hypertension, or poor selfrated health. | Type of measure: Self-reported: Leisure time of PA: was based on the individual’s response to the following item. (1) I get practically no exercise at all; (2) I exercise occasionally (e.g., 1-hour walks, skiing a couple of times every year, swimming, picking mushrooms); (3) I exercise about once a week (e.g., fast walks, skiing, swimming, jogging, cycling); (4) I exercise about twice a week (e.g., fast walks, skiing, swimming, jogging, cycling); (5) I exercise vigorously at least twice a week (e.g., skiing, swimming, running, cycling for quite a while, ball games).  LTPA exposure: 1) none, 2) occasionally, 3) once a week, 4) twice a week, 5) vigorously at least twice a week | N = 3206 (Men 794, 516 women). Physical activity: 1) none (1100 men and 776 women), 2) occasionally (744 men and 480 women), 3) once a week (555 men and 393 women), 4) twice a wee (495 men and 331 women)k, 5) vigorously at least twice a week 573 men and 400 women) | Model 1 is adjusted for gender, age, and education. Model 2 is also adjusted for physical activity, smoking habits, and body mass index. Model 3 is adjusted for all explanatory variables | Only those who were interviewed directly by the interviewer were included in the sample. Participants who were interviewed through others, such as relatives, were excluded.  Cox regression model was used to estimate the hazard ratio (HR) of all-cause mortality for the variables. Model 1 is adjusted for gender, age, and education. Model 2 is also adjusted for physical activity, smoking habits, and body mass index. Model 3 is adjusted for all explanatory variables | All-cause mortality | All-cause mortality  1806 deaths HR and 95%CI: 1 (none) (Reference): 2 (occasionally): 0.72 (0.64, 0.81) 3 (once a week): 0.60 (0.50, 0.71) 4 (twice a week): 0.50 (0.42, 0.59) 5 (vigorously at least twice a week): 0.60 (0.46, 0.79) |
| Wang et al., [29] | China / Shanghai | Shanghai Men’s Health Study (SMHS) is a population-based, prospective cohort study of 61491 men who were 40–74 years of age at baseline | 61477 participants / During a mean follow-up of 5.48 years, 2421 deaths were documented, including 1053 from cancer, 800 from CVD, and 568 from other causes | Compared with men who did not exercise, men who participated in regular exercise were older (60.3 vs 52.7 years), had a higher socioeconomic status, and were less likely to have ever drunk alcohol (34.4 vs 31.5) or smoked cigarettes (73.3 vs 62.6). Regular exercisers tended to have a higher prevalence of CVD (4.9% vs 4.5%), diabetes (7.1% vs 5.9%), and hypertension (31.7% vs 28.5%) but a lower prevalence of pulmonary disease (6.8% vs 7.2%). | Type of measure: Self-reported: Leisure time of PA: Exercise on a weekly basis. Regular exercise was defined as engaging in leisure-time exercise of any type and intensity at least once per week. LTPA exposure: Any type of exercise: <13.9 MET-hours/week and ≥ 13.9 MET-hours/week | Of the 61477 men in the SMHS, 35.5% (21847) regularly engaged in 1 or more exercises, and 57.3% (12520) of those men spent an average of more than 30 minutes per day on exercise. The most common types of exercise included walking, Tai Chi, and jogging | Model 1 Adjusted for age; educational level; income; occupation; alcohol consumption; pack-years of smoking; daily intake of energy, red meat, fruits, and vegetables; daily physical activity other Model 2 Excluding participants who reported a history of CVD at baseline or died within the first year of follow-up. Model 3 Additionally adjusted for exercise other than the specific type presented. | Participants with previously diagnosed cancer were excluded. Of the eligible men approached for the study, 61,582 completed an in-person interview, with a response rate of 74.1%. Reasons for nonparticipation included refusal (21%), serious health problems (2%), and absence during the study period (3%). | All-cause mortality. Cancer mortality. CVD mortality. | Any type of exercise <13.9 MET-hours/week - Deaths: All-cause 424, cancer 180, CVD mortality 147 ≥ 13.9 MET-hours/week- Deaths: All-cause 691, cancer 301, CVD mortality 234 All-cause mortality  HR and 95%CI: No regular exercise (reference) <13.9 MET-hours/week: 0.81 (0.73, 0.91) ≥ 13.9 MET-hours/week: 0.79 (0.72, 0.88) Cancer mortality HR and 95%CI: No regular exercise (reference) <13.9 MET-hours/week:0.81 (0.68, 0.96) ≥ 13.9 MET-hours/week: 0.81 (0.69, 0.94) CVD mortality HR and 95%CI: No regular exercise (reference) <13.9 MET-hours/week: 0.82 (0.68, 1.00) ≥ 13.9 MET-hours/week: 0.76 (0.64, 0.90) |
| Wang et al., [30] | USA | The National Health Interview Survey (NHIS), conducted by the Centers for Disease Control and Prevention from the National Center for Health Statistics, is an annual national cross-sectional survey of civilian, noninstitutionalized participants from the US. The survey uses a stratified, multistage sample design select approximately 35000 households from randomly selected clusters. | 403 681 individuals / 36 861 deaths occurred, including 7634 from CVD and 8902 from cancer. | Among the 403 681 individuals (225 569 women [51.7%];mean [SD] age, 42.8 [16.3] years) in the study, during a median 10.1 years (interquartile range, 5.4-14.6 years) of follow-up (407.3 million person-years), 36 861 deaths occurred, including 7634 from CVD and 8902 from cancer. | Type of measure: Self-reported: Leisure time of PA: Frequency of light, moderate and vigorous intensity physical activity. LTPA exposure: Proportion of VPA to total MVPA: >0% to ≤25%, >25% to ≤50%, >50% to ≤75%, >75% to <100% and 100%. Also, MPA (0, 1-149, 150-299, and ≥300 min/wk) and VPA (0, 1-74, 75-149, and ≥150 min/wk) | Less than half (45.0%) of the participants met the physical activity guidelines and 34.3%had no MVPA.Among those who reported any MVPA (65.7%), the proportions of VPA to total physical activity were distributed as follows: 32.5%had 0% of VPA (no vigorous activity), 5.1%had 25%or more of VPA, 10.0%had more than 25% to 50% of VPA, 21.3% had more than 50% to 75% of VPA, 15.8%had more than 75%to less than 100% of VPA, and 15.2% had 100% of VPA (all vigorous activity). Participantswho were younger, men, non-Hispanic White, with a higher educational level, with a normal BMI (<25), with a high income level, andwith no smoking historywere more likely to report 25% or more of VPA to total physical activity | Models were adjusted for the major potential confounders, including age at baseline (as a continuous variable), sex, race/ethnicity , educational level, income, marital status, BMI, smoking status, alcohol consumption), and total amount of physical activity (0, 1-149, 150-299, and ≥300 min/wk). | Excluded thosewith missing data on physical activity (n = 14 994), those with disabilities (needing help for daily life activities) (n = 10388) or unable to perform moderate or vigorous physical activity (n = 10725), and those with a diagnosis of heart disease, stroke, or cancer at baseline (n = 53 577). | All-cause mortality.  CVD mortality. Cancer mortality. | Proportion of VPA to MVPA All-cause mortality No of deaths 0 - 9241 ≥0.25 - 632 >0.25 to 0.5 - 1071 >0.5 to 0.75 - 2100 >0.75 to <1 - 1413 1 – 2148 CVD mortality No of deaths 0 - 1919 ≥0.25 - 116 >0.25 to 0.5 - 190 >0.5 to 0.75 - 346 >0.75 to <1 - 220 1 – 414 Cancer mortality No of deaths 0 - 2338 ≥0.25 - 183 >0.25 to 0.5 - 329  >0.5 to 0.75 - 577 >0.75 to <1 - 428 1 – 591  Compared with participants with 0% of VPA, participants with more than 50% to75% of VPA to total physical activity had a 17% lower all-cause mortality (HR, 0.83; 95% CI, 0.78-0.88), independent of total MVPA. The HR for participants with more than 75% to 99% of VPA was 0.85 (95%CI,0.79-0.91) compared with 0% VPA. Mutually adjusted models considering the recommendations of MPA (150-299 vs 0 min/wk) and VPA (≥75-149 vs 0 min/wk) showed similar associations for all-cause mortality (MPA: HR,0.83; 95%CI,0.80-0.87; and VPA: HR,0.80; 95%CI,0.76-0.84) and CVD mortality (MPA: HR, 0.75; 95% CI, 0.68-0.83; and VPA: HR, 0.79; 95% CI, 0.70-0.91). For the same contrasts, VPA (HR, 0.89; 95% CI, 0.80-0.99) showed a stronger inverse association with cancer mortality compared with MPA (HR, 0.94; 95% CI, 0.86-1.02). Exploratory joint associations of MPA and VPA (reference: no MVPA) suggested the lowest all-cause mortality risk among participants performing 150 to 299 min/wk of MPA and 150 min/wk or more of VPA (HR, 0.64; 95%CI, 0.58-0.71). For CVD mortality, the optimum MPA and VPA combination was 1 to 149 min/wk of MPA and 150 min/wk or more of VPA (HR,0.56; 95% CI, 0.45-0.69). For cancer mortality, 300 min/wk or more of MPA and 1 to 74 min/wk of VPA showed the strongest inverse association (HR, 0.67; CI, 0.52-0.86). |
| Zhao et al., [31] | USA | Data from 366376 study participants aged 18–85 years who participated in 13 cross-sectional waves of the National Health Interview Surveys (NHIS) during 1997–2009. The NHIS is an ongoing national cross-sectional survey administered by the National Center for Health Statistics of the Centers for Disease Control and Prevention (CDC) to monitor the health of the civilian noninstitutionalized US population. | 88 140 / There were 7855 all-cause deaths, 1 695 CVD-specific deaths and 2269 cancer-specific deaths, during a median follow-up of 9.0 years, | Mean age 65 (60-71) years. 8.2% current smokers, 27.$% with hipertension, 4.6% with diabetes, and 18.7% with high cholesterol. | Type of measure: Self-reported: Leisure time of PA: Frequency and duration of PA (eg, running, faster cycling and competitive sports, etc.) and light or moderate PA (eg, brisk walking, dancing and gardening, etc.). LTPA categories: 0, 10.59, 60-149, 150-299, 300-449, 450-799, 1800-1499, ≥1500 min/wk. Also 1-2 times, 2-3 times, 3-5 times, and 5-10 times or 10 or more times. | Participants (n) by leisure time PA level (min/wk): 0 (n= 36701), 10–59 (n= 3695), 60–149 (n = 11739), 150–299 (n= 11580), 300–499 (n= 8280), 450-799 (n=8446), 800-1499 (n= 5198) and ≥1500 (n= 2500) | Model 1 included sex, age and race/ethnicity;  Model 2 included variables in Model 1 plus education and marital status; and  Model 3 included variables in Model 2 plus lifestyle variables | A multivariable Cox proportional hazards regression model was used to estimate the risk of all-cause and cause-specific mortality with level of total leisure time PA, adjusting for potential confounding factors including demographic factors and lifestyle behaviours.  Model 1 included sex, age and race/ethnicity;  Model 2 included variables in Model 1 plus education and marital status; and Model 3 included variables in Model 2 plus lifestyle variables | All-cause mortality. CVD mortality. Cancer mortality. | All cause  5074 deaths 10–59 min/week of leisure time PA had 18% lower risk of all-cause mortality (HR=0.82,95% CI=0.72–0.95) than physically inactive ones. Those who reported 60–149 min/wk of leisure time PA (HR=0.78, 95% CI=0.72–0.86). Those who reported 1–2 times (150–299 min/week) or 2–3 times (300–449 min/week) the recommended level of leisure time PA had 31% (HR=0.69, 95% CI=0.63–0.75) and 33% (HR=0.67,95% CI=0.61–0.74) lower risk of all-cause mortality. Similar benefits were found for PA levels at 3–5 times (450–799 min/week: HR=0.58, 95% CI=0.52–0.66), at 5–10 times (800–1499 min/week: HR=0.63, 95% CI=0.56–0.72), and at 10 or more times (≥1500 min/week) the recommended minimum by the PA guidelines (HR=0.54, 95% CI=0.45–0.64. CVD mortality 1132 deaths The HRs and 95% CIs for CVD-specific mortality for 10–59, 60–149, 150–299, 300–449, 450–799, 800–1499 and ≥1500 min/wk leisure time PA were 0.88 (0.67–1.17), 0.76 (0.63–0.92), 0.63 (0.52–0.78), 0.64 (0.49–0.82), 0.64 (0.49–0.83), 0.74 (0.57–0.97) and 0.67 (0.45–0.99), respectively.  Cancer mortality:  1341 deaths The corresponding HRs and 95% Ci for cancer-specific mortality were 0.86 (0.66–1.11), 0.84 (0.72– 0.98), 0.76 (0.64–0.89), 0.85 (0.71–0.99), 0.71 (0.59–0.86), 0.65 (0.51–0.83) and 0.53 (0.39–0.73), respectively |
| Arem et al., [32] | USA | Pooled data from six studies in the NCI Cohort Consortium (baseline 1992–2003) | 661137 participants/116686 deaths during a mean of 14.2 years of follow-up | Median Age: 62 (SD) y % women: 55,91% | Type of measure: Self-reported: Type of LTPA: Walking, jogging/running, swimming, tennis/racquetball, bicycling, aerobics and dance, horseback riding, or in strenuous activities LTPA exposure: overall leisure-time physical activity energy expenditure in MET h/wk, Leisure-time moderate- to vigorous-intensity physical activity.  Categories of LTPA by intensity:  moderate (3-<6 METs) and vigorous (6+ <METs); MET h/wk. Categories were 0, 0.1-<7.5, 7.5-<15, 15-<30, and 30+ due to a lower range of MET h/wk Categories of LTPA by total:  1–2 times the recommended minimum (7.5-<15 MET h/wk) to up to 10+ times the recommended levels (75+ MET h/wk) (Exercise levels compared to the federally recommended minimum of 7.5 MET h/wk). Categories were MET h/wk: 0, 0,1-<7,5, 7,5-<15, 15-<22,5, 22,5-<40, 40-<75 y 75+ | Median physical activity level: 8 MET h/wk (interquartile range 4–22)  Categories of LTPA : Level of LTPA n (%) 0 MET: 52 848 (8.0) 0.1-<7.5 MET h/w: 170563 (25.8); 7.5-<15 MET h/wk: 100687 (28.9) 15-<22.5 MET h/wk 118169 (17.9); 22.5-<40 MET h/wk: 124 446 (18.8) 40-<75 MET h/wk: 18 831 (2.9); 75+ MET h/wk: 4077 (0.6)  Categories of LTPA by intensity: n (%) 0 MET h/wk: 53376 (15.3) 0.1-<7.5 MET h/wk: 122522 (35.1) 7.5-<15 MET h/wk: 100 687 (28.9) 15-<30 MET h/wk: 59 304 (17.0) 30+ MET h/wk: 12 836 (3.7) | Analysis 1 included LTPA level into the multivariable model Analysis 2: included leisure time physical activity and mortality, by activity intensity into de multivariable model Analysis 3: risk of cardiovascular mortality in those with changes in LTPA after five years for high-intensity activity and the most common activities into de multivariable model | Individuals with missing BMI data or reporting a BMI of less than 15 or more than 60.   Model 1: stratified by cohort, adjusted for age, and mutually adjusted for both moderate- and vigorous-intensity activities.  Model 2: aditionally adjusted for age, gender, race, education, smoking status, cancer history, heart disease, alcohol consumption, marital status, and BMI | All-cause mortality. CVD mortality. Cancer mortality. | All cause: 2564 deaths HR and 95% CI (Model 2) Level of LTPA 0 MET h/wk (reference) vs  (0.1-<7.5 MET h/wk) - 0.80 (0.78–0.82); 7.5-<15 MET h/wk - 0.69(0.67–0.70); 15-<22.5 MET h/wk - 0.63, (0.62–0.65); 22.5-<40 MET h/wk - 0.61(0.59–0.62); 40-<75 MET h/wk - 0.61(0.58–0.64); 75+ MET h/wk - 0.68(0.59–0.78) Activity intensity of LTPA Moderate-intensity activity 0 MET h/wk (reference) vs  0.1-<7.5 MET h/wk- 0.80(0.78–0.83); 7.5-<15 MET h/wk - 0.73 (0.71–0.75); 15-<30 MET h/wk - 0.71 (0.68–0.73); 30+ MET h/wk - 0.72 (0.68–0.76) Vigorous-intensity activity 0 MET h/wk (reference) vs 0.1 -7.5 MET h/wk - 0.80 (0.78–0.83); 7.5-<15 MET h/wk - 0.77 (0.71–0.84); 15-<30 MET h/wk - 0.78 (0.73–0.83); 30+ - 0.79 (0.73–0.85) Cancer mortality  3143 deaths HR and 95% CI (Model 2) 0 MET h/wk (reference) vs  (0.1-<7.5 MET h/wk) - 0,87 (0,83–0,90); 7.5-<15 MET h/wk - 0,79 (0,75–0,82); 15-<22.5 MET h/wk - 0,75 (0,72–0,79); 22.5-<40 MET h/wk - 0,74 (0,71–0,77); 40-<75 MET h/wk - 0,72 (0,66–0,79); 75+ MET h/wk - 0,69 (0,55–0,87) CVD mortality: 3238 deaths HR and 95% CI (model 2) 0 MET h/wk (reference) vs  (0.1-<7.5 MET h/wk) - 0,80 (0,77–0,84); 7.5-<15 MET h/wk - 0,67 (0,65–0,70); 15-<22.5 MET h/wk - 0,59 (0,57–0,63); 22.5-<40 MET h/wk -0,58 (0,56–0,61); 40-<75 MET h/wk - 0,61 (0,55–0,67); 75+ MET h/wk - 0,71 (0,56–0,91) |
| Bergwall et al., [33] | Sweden / Malmo | Prospective cohort study (baseline 1991-1996 and the five-year follow-up) In total, the cohort consists of 30446 men and women born between 1923 and 1950. Among the participants, 28098 individuals completed an extensive lifestyle questionnaire, underwent anthropometric measurements, and completed dietary assessments. This resulted in a study population of 25876 individuals. | 25876 participants/2564 deaths during a mean of 20 years of follow-up. | Mean Age: Category 1 (≤7.5 MET-h/week): 57.6 (7.4); Category 2 (7.5–15 MET-h/week): 57.4 (7.4); Category 3 (15–25 MET-h/week): 57.6 (7.5); Category 4 (25–50 MET-h/week): 57.7 (7.7); Category 5 (> 50 MET-h/week): 59.0 (7.9) % women: Category 1 (≤7.5 MET-h/week): 60; Category 2 (7.5–15 MET-h/week): 62; Category 3 (15–25 MET-h/week): 64; Category 4 (25–50 MET-h/week): 64; Category 5 (> 50 MET-h/week): 58  Hypertension at baseline was prevalent in 76–78% of the participants across the categories. | Type of measure: Self-reported: extensive lifestyle questionnaire Type of LTPA: 17 common physical activities in their spare time (orienteering, walking up stairs, high intensity, lawn tennis, Running, Soccer, swimming, ball sports, ballroom dancing, grass cutting, digging, Badminton, golk dancing, golf, cycling, gardening, gymnastics, table tennis, walking). LTPA exposure: Total leisure-time physical activity in MET-h/week Categories of LTPA by intensity:  VPA: estimated MET factor > 6, Categories LTPA By Total:  Category 1 (≤7.5 MET-h/week) Category 2 (7.5–15 MET-h/week; Category 3 (15–25 MET-h/week); Category 4 (25–50 MET-h/week); Category 5 (> 50 MET-h/week) | Categories of LTPA: (n) Category 1 (≤7.5 MET-h/week); 2434  Category 2 (7.5–15 MET-h/week): 3869 Category 3 (15–25 MET-h/week): 5953  Category 4 (25–50 MET-h/week): 9490 Category 5 (> 50 MET-h/week): 4130 | Analysis 1 included total leisure-time physical activity into the multivariable model  Analysis 2: included participation in different activities into the multivariable model    Analysis 3: risk of cardiovascular mortality in those with changes in LTPA after five years for high-intensity activity and the most common activities | Individuals with prevalent diabetes or cardiovascular disease at baseline, individuals who died within the first year of follow-up and those with unrealistic leisure-time physical activity levels exceeding > 50 h/week. Model 1: Adjusted for age, sex and screening date Model 2: Adjusted for age, sex, screening date, education, smoking status, alcohol, diet index and total energy intake Model 3: Adjusted for age, sex, screening date, education, smoking status, alcohol, diet index and total energy intake and BMI Model 4: Adjusted for age, sex, screening date, education, smoking status, alcohol, diet index, total energy intake and mutually adjusted for the 17 activit | CVD mortality. | CVD mortality: 2564 deaths HR and 95% CI (Model 3) Total LTPA  ≤7.5 MET-h/week vs:  7,5–15 MET-h/semana - 0,96 (0,82–1,13); 15–25 MET-h/semana - 0,84 (0,72–0,97); 25–50 MET-h/semana - 0,87 (0,76–1,01); > 50 MET-h/week - 0,87 (0,75–1,02).  high-intensity leisure-time physical activity High-intensity exercise No activity vs tertile: 1 (Mean MET-h/week: 2,29; Mean hours/week: 0,33) - 0,82 (0,72–0,93);  2 (Mean MET-h/week: 6,53; Mean hours/week: 0,93) - 0,82 (0,69–0,97); 3 (Mean MET-h/week: 18,25; Mean hours/week: 2,57) - 0.84 (0,70–0,99)  Running No activity vs tertile: 1 (Mean MET-h/week: 2,29; Mean hours/week: 0,33) - 0,63 (0,46–0,88);  2 (Mean MET-h/week: 6,53; Mean hours/week: 0,93) - 0,65 (0,46–0,91); 3 (Mean MET-h/week: 18,25; Mean hours/week: 2,57) - 0,71 (0,53–0,94) Swimming No activity vs tertile: 1 (Mean MET-h/week: 2,29; Mean hours/week: 0,33) - 0,85 (0,71–1,02);  2 (Mean MET-h/week: 6,53; Mean hours/week: 0,93) - 0,89 (0,75–1,04); 3 (Mean MET-h/week: 18,25; Mean hours/week: 2,57) - 1,01 (0,85–1,21) Cycling No activity vs tertile: 1 (Mean MET-h/week: 2,29; Mean hours/week: 0,33) - 0,79 (0,71–0,89);  2 (Mean MET-h/week: 6,53; Mean hours/week: 0,93) - 0,84 (0,76–0,94); 3 (Mean MET-h/week: 18,25; Mean hours/week: 2,57) - 0,93 (0,83–1,03) Gymnastics No activity vs tertile: 1 (Mean MET-h/week: 2,29; Mean hours/week: 0,33) - 093 (0,81–1,07);  2 (Mean MET-h/week: 6,53; Mean hours/week: 0,93) - 0,77 (0,60–1,00) 3 (Mean MET-h/week: 18,25; Mean hours/week: 2,57) - 0,94 (0,79–1,12) |
| Besson et al., [34] | United Kingdom / Norfolk | EPIC study is a is a prospective cohort study (1993 - 1997), EPIC-Norfolk (UK) recruited a population-based cohort of 25639 men and women aged 45-79, identified from participating general practice lists. | 14903 participants/2256 death After a median follow-up of 7 years | Mean age:  Working: 57,1 (7,2)  Nonworking: 68.0 (7,2)   % women:  Working: 53,9  Nonworking: 58.6 | Type of measure: Self-reported: EPIC Physical Activity Questionnaire (EPAQ2) Type of LTPA: Recreational physical activity; LTPA exposure: A score for physical activity by the energy expenditure in MET h/wk  Categories of PA by total:  inactive: <60 MET h/wk; moderately inactive: between 60 and 90 MET h/wk; moderately active: between 90 and 120 MET h/wk; active: >125 MET h/wk   Categories by PA for sport or exercise Inactive (<2 MET h/wk): 386 Moderately inactive (2–9 MET h/wk): 279 Moderately active (9–20 MET h/wk): 233 | Physical activity : MET h/wk  Working:  All domains combined: 122.1 (53.7) For sport or exercise 14.4 (16.7) Nonworking:  All domains combined: 72.3 (37.3)  For sport or exercise 14.7 (18.5)   Categories of LTPA: Total PA inactive (<60 MET h/wk): 526 moderately inactive (60 -90 MET h/wk): 312  moderately active (90 and 120 MET h/wk): 161 Active (>125 MET h/wk) : 129   Total PA (n) inactive (<60 MET h/wk): 526 moderately inactive (60 -90 MET h/wk): 312  moderately active (90 and 120 MET h/wk): 161 Active (>125 MET h/wk) : 129   PA for sport or exercise (n) Inactive (<2 MET h/wk): 386 Moderately inactive (2–9 MET h/wk): 279 Moderately active (9–20 MET h/wk): 233 Active >20 MET h/wk): 230 | Analysis 1 included total physical activity into the multivariable model  Analysis 2: included PA for sport or exercise into the multivariable model | Participants who died within 2 yr of follow-up and participants who had a history of heart disease, stroke, or cancer.  Final model: adjustment for baseline age, sex, social class, alcohol consumption, smoking status, history of diabetes, history of cancer, and history of cardiovascular disease and stroke. When examining the domain-specific association of physical activity with mortality, all analyses were additionally adjusted for the other domains of activity.  PA for sport or exercise: additionally adjusted for activity at home, at work, and for transportation | All-cause mortality. CVD mortality. | All-cause mortality: 1028 deaths HR and 95% CI  Total PA Inactive (<60 MET h/wk) Vs: moderately inactive: between 60 and 90 MET h/wk - 0.87 (0.73–1.03); moderately active (between 90 and 120 MET h/wk) - 0.67 (0.54–0.83) active ( >125 MET h/wk) - 0.77 (0.61–0.98) PA for sport or exercise Inactive (<60 MET h/wk) Vs: moderately inactive: between 60 and 90 MET h/wk - 0.81 (0.67–0.973); moderately active (between 90 and 120 MET h/wk) - 0.73 (0.60–0.88) active ( >125 MET h/wk) - 0.66 (0.54–0.80) CVD mortality: 370 deaths HR and 95% CI  Total PA Inactive (<60 MET h/wk) Vs: moderately inactive: between 60 and 90 MET h/wk - 0.88 (0.65–1.18); moderately active (between 90 and 120 MET h/wk) - 0.61 (0.41–0.91); active ( >125 MET h/wk) - 0.77 (0.61–0.98) PA for sport or exercise Inactive (<60 MET h/wk) Vs: moderately inactive: between 60 and 90 MET h/wk - 0.78 0.56–1.10); moderately active (between 90 and 120 MET h/wk) - 0.71 (0.51–1.00); active ( >125 MET h/wk) - 0.60 (0.42–0.85) |
| Dos Santos et al., [36] | United States | The NHIS is a nationally representative survey of civilian, noninstitutionalized participants in the USA. In brief, the NHIS conducted in 1997 to 2013 used a stratified, complex multistage sampling design to select households from random clusters. From these households, a sample of adults (≥18 years) was randomly selected to respond to a questionnaire. | 350978 participants/32062 deaths were followed during a median of 10.4 years (3.6 million person-years) | Mean [SD] age, 41.4 [15.2] years; 192 432 [50.8%] women; 209 432 [67.8%] [14.5%] Hispanic, 52 620 [12.1%] non-Hispanic Black, and 209 432 [67.8%]non-HispanicWhite individuals). | Type of measure: self-reported physical activity to the US National Health Interview Survey Type of LTPA: Moderate to Vigorous leisure-time physical activities LTPA exposure: Participants were grouped by self-reported activity level; The active group was further classified by frequency, duration/session, and intensity of activity) Categories of LTPA by activity level:  physically inactive (<150 minutes per week [min/wk] of MVPA) or physically active (150 min/wk of moderate or75 min/wk of vigorous activity) Categories of PA by frecuency (active group):  weekend warrior (1-2 sessions/wk) or regularly active (3 session/wk)  Categories of MVPA by session duration (physically active group):  ≤20 min, >20-30 min, >30-60 min, or >60 min/session Categories of MVPA by intensity (physically active):  0% (all min at moderate intensity), 1% to 24%, 25% to 49%, 50% to 74%, 75% to 99%, and 100% (all min at vigorous intensity) | *Categories of LTPA: n* participants  Inactive: 190 080 Weekend warrior: 9992 Regularly active: 150 906 | Analysis 1: included BMI in the primary model  Analysis 2: included the total amount of MVPA into the model Analysis 3: when we investigated whether, for the same amount of total MVPA, weekly frequency, duration of sessions, and intensity of physical activity were associated with mortality. Analysis 4: excluded participants with ≥600 min/wk of physical activity and ≥780 min/wk of physical activity | Participants who had been diagnosed with cancer, chronic bronchitis, emphysema, heart disease,and stroke at baseline; with missing data for physical activity; with limitations on their instrumental and other activities of daily living; or who were unable to perform moderate or vigorous physical activity. Lastly, we excluded the first 2 years of follow-up to further mitigate the influence of reverse causation bias  Model 1: Adjusted for age, sex, race and ethnicity, education, income, marital status, smoking, alcohol intake, self-rated health, psychological distress, number of comorbidities, and mobility.  Model 2: Model 1 + body mass index  Model 3: Model 1 + total volume of physical activity | All-cause mortality. CVD mortality. Cancer mortality. | All-cause mortality: 21898 deaths HR and 95% CI  Physical Activity Pattern (Model 3) Inactive (reference) vs  Weekend warrior (1-2 sessions/wk) - 0.92 (0.83-1.02) Regularly active (3 session/wk) - 0.85 (0.82-0.88) Inactive vs Regularly active (Model 1) 0.85 (0.83-0.88)  Frequency, sessions/wk  Inactive (reference) vs 3-4 - 0.81 (0.76-0.86); ≥5 - 0.87 (0.83-0.90); Duration of session, min  Inactive (reference) vs ≤20 - 1.02 (0.86-1.21); >20-≤30 - 0.92 (0.85-0.99); >30-≤60 - 0.85 (0.80-0.89); >60 - 0.83 (0.79-0.87) Intensity (VPA/MVPA), % Inactive (reference) vs 0 - 0.91 (0.86-0.95); 1-25 - 0.80 (0.71-0.91); 26-50 - 0.94 (0.86-1.03); 51-75 - 0.80 (0.75-0.85); 76-99 - 0.81 (0.75-0.88); 100 - 0.83 (0.76-0.90); Inactive (reference) vs The weekend warriors (Model 1) 0.92 (0.83-1.02)  Frequency, sessions/wk  Inactive (reference) vs 1 - 0.87 (0.71-1.06); 2 - 0.94 (0.83-1.05); Intensity (VPA/MVPA), % Inactive (reference) vs 0 - 0.98 (0.84-1.14); 1-25 - 0.63 (0.33-1.19); 26-50 0.88 (0.55-1.39); 51-75 - 0.71 (0.46-1.10); 76-99 - 0.84 (0.57-1.26); 100 - 0.94 (0.80-1.11) Regular active (reference) vs the weekend warriors (model 3) 1.08 (0.97-1.20) Frequency, sessions/wk  Inactive (reference) vs 1 - 1.01 (0.82-1.24); 2 - 1.10 (0.97-1.24); Intensity (VPA/MVPA), % Inactive (reference) vs 0 - 1.16 (0.99-1.35) 1-25 - 0.74 (0.39-1.41) 26-50 - 1.04 (0.66-1.67) 51-75 0.84 (0.55-1.30) 76-99 - 0.98 (0.65-1.46) 100 - 1.08 (0.92-1.27) CVD mortality: 4130 deaths HR and 95% CI  Physical Activity Pattern (Model 3) Inactive (reference) vs  Weekend warrior (1-2 sessions/wk) - 0.87 (0.66-1.15) Regularly active (3 session/wk) - 0.77 (0.71-0.84) 0.76 ( Inactive vs Regularly active (Model 1) 0.77 (0.71-0.84  Frequency, sessions/wk  Inactive (reference) vs 3-4 - 0.81 (0.68-0.97) ≥5 - 0.76 (0.70-0.83) Duration of session, min  Inactive (reference) vs ) ≤20 - 0.85 (0.56-1.31) >20-≤30 - 0.75 (0.62-0.90) >30-≤60 - 0.77 (0.68-0.87) >60 - 0.77 (0.69-0.87) Intensity (VPA/MVPA), % Inactive (reference) vs 0 - 0.74 (0.65-0.85) 1-25 - 0.88 (0.67-1.16) 26-50 - 0.89 (0.71-1.13); 51-75 - 0.73 (0.62-0.87) 76-99 - 0.64 (0.51-0.80) 100 - 0.92 (0.77-1.09) Inactive (reference) vs The weekend warriors (Model 1) 0.87 (0.66-1.15) Frequency, sessions/wk  Inactive (reference) vs 1 - 1.14 (0.71-1.83) 2 - 0.75 (0.55-1.02) Intensity (VPA/MVPA), % Inactive (reference) vs 0 - 0.97 (0.66-1.42) 1-25 - 0.45 (0.06-3.21) 26-50 NA 51-75 - 0.52 (0.19-1.41) 76-99 - 1.28 (0.63-2.62) 100 - 0.86 (0.53-1.40) Regular active (reference) vs the weekend warriors (model 3) 1.14 (0.85-1.53) Frequency, sessions/wk  Inactive (reference) vs 1 - 1.44 (0.89-2.32) 2 - 0.97 (0.71-1.33) Intensity (VPA/MVPA), % Inactive (reference) vs 0 - 1.22 (0.82-1.82) 1-25 - 0.58 (0.08-4.2) 26-50 - NA 51-75 0.68 (0.25-1.85) 76-99 - 1.62 (0.78-3.35) 100 - 1.12 (0.69-1.81) Cancer mortality: 6034 deaths HR and 95% CI  Physical Activity Pattern (Model 3) Inactive (reference) vs  Weekend warrior (1-2 sessions/wk) - 0.94 (0.77-1.14) Regularly active (3 session/wk) - 0.88 (0.82-0.94) Inactive vs Regularly active (Model 1) 0.88 (0.83-0.94) Frequency, sessions/wk  Inactive (reference) vs 3-4 - 0.83 (0.73-0.93); ≥5 - 0.90 (0.84-0.96) Duration of session, min  Inactive (reference) vs ≤20 - 1.11 (0.84-1.47) >20-≤30 - 0.98 (0.86-1.11) >30-≤60 - 0.87 (0.80-0.96) >60 - 0.85 (0.77-0.93) Intensity (VPA/MVPA), % Inactive (reference) vs 0 - 0.93 (0.84-1.03) 1-25 - 0.75 (0.60-0.94) 26-50 - 1.01 (0.86-1.18) 51-75 - 0.76 (0.67-0.86) 76-99 - 0.90 (0.78-1.05) 100 - 0.94 (0.82-1.08) Inactive (reference) vs The weekend warriors (Model 1) 0.94 (0.77-1.15) Frequency, sessions/wk  Inactive (reference) vs 1 - 0.63 (0.40-1.00) 2 - 1.06 (0.85-1.32) Intensity (VPA/MVPA), % Inactive (reference) vs 0 - 1.12 (0.84-1.49) 1-25 - 0.84 (0.28-2.49); 26-50 0.94 (0.36-2.49); 51-75 - 0.54 (0.26-1.11) 76-99 - 0.64 (0.26-1.56) 100 - 0.90 (0.65-1.26) Regular active (reference) vs the weekend warriors (model 3) 1.07 (0.87-1.31) Frequency, sessions/wk  Inactive (reference) vs 1 - 0.74 (0.47-1.17) 2 - 1.23 (0.98-1.54) Intensity (VPA/MVPA), % Inactive (reference) vs 0 - 1.29 (0.96-1.72) 1-25 - 0.97 (0.33-2.90) 26-50 - 1.11 (0.42-2.94) 51-75 0.62 (0.30-1.29) 76-99 - 0.76 (0.31-1.87) 100 - 1.06 (0.76-1.47) |
| Gebel et al., [37] | Australia/New South Wales | The baseline data were collected in 2006-2009. Individuals 45 years and older were randomly sampled from the Medicare Australia database, through which national health care is administered and includes all citizens and permanent residents of Australia and some temporary residents and refugees. Approximately 10%ofthe whole population of New South Wales 45 years and older were included in the final sample. | 204542 participants/7435 deaths (mean [SD] follow-up, 6.52 [1.23] years). | Mean Age: not reported: Age was divided into 3 intervals  45-54 yr: 76428 55-64 yr: 84174 65-75 yr: 57153  55.2% were women, 63.8% were overweight or obese, 25.5% had completed a university degree | Type of measure: Self-reported: Active Australia Survey. Type of LTPA: walking, moderate (like gentleswimming, social tennis, vigorous gardening or work around the house), and vigorous activity (jogging, cycling, aerobics, competitive tennis, but not household chores or gardening) LTPA exposure: Different proportions of total MVPA as vigorous activity  Categories for the percentage of total MVPA that was vigorous: :  no vigorous activity (0%), some vigorous activity (>0% to <30%), and 30% or more of total MVPA as vigorous activity Categories of Total MVPA  0, 10 to 149, 150 to 299, and 300 min/wk or more | 78.3% met minimal activity recommendations, and 62.1% met the new Australian high active recommendation 40 of 300 min/wk or more of activity.   Categories of LTPA: who reported some MVPA, more than half (55.3%) did not report any vigorous activity, 16.3% reported more than 0% to less than 30%, and 28.4 reported 30% or more of their total MVPA as vigorous activity. Younger, better educated, and male participants were more likely to report 30% or more of their MVPA as vigorous activity, and those with underweight or obese BMI and with poor physical function scores were less likely to report this.  n (%) No MPVA: 13 213 (6.1) Vigorous Activity: 0%: 113 181 (52.0) >0% to <30%: 33 371 (15.3) ≥30%: 57 990 (26.6)  Total MVPA, min (n) 0: 13 213 10-149: 34 053 150-299: 35 302 ≥300: 135 187 | Analysis 1: Stratified by volume of MVPA  Analysis 2: We examined effect modification by amount of MVPA using likelihood ratio tests to compare modelswith and without an interaction term  Anaysis 3: Further stratified analyseswere conducted to compare the models of each subsample. | Participants older than 75 years. Those with missing values for physical activity. those who did not report any activity  Model 1: adjusted for baseline age, sex, educational level, marital status, urban or rural residence, BMI, smoking status, physical function, Aalcohol consumption, fruit and vegetable consumption, and total MVPA. | All-cause mortality. | all-cause mortality: 7435 deaths HR and 95% CI no MPVA (reference) VS 10 -149 min/wk - 0.66 (0.61-0.71),  150 - 299 min/w - 0.53 (0.48-0.57),  ≥300 min/wk - 0.46 (95%CI, 0.43-0.49, 10 -149 min/wk (reference) Vs >0% to <30% - 1.09 (0.84-1.42)  ≥30% - 0.76 (0.63-0.92) 150 - 299 min/w (reference) VS >0% to <30% - 0.83 (0.67-1.03)   ≥30% - 1.03 (0.88-1.20) ≥300 (reference) Vs >0% to <30% - 0.90 (0.82-0.98)   ≥30% - 0.85 (0.78-0.92) |
| Huerta et al., [38] | Spain / Asturias, Gipuzkoa, Navarra, Granada, and Murcia | The EPIC (European Prospective Investigation into Cancer and Nutrition) study is a multi-center prospective investigation on half a million volunteers from ten European countries ). The Spanish branch (EPIC-Spain) ncluded 41,438 participants (62.3% women), 29 to 69 years old at baseline, recruited between 1992 and 1996. | 38379 participants/ 1371 After 13.6 years of mean follow-up | 62.4% women, 30–65 years old, and free of chronic disease  men age  Q1 (<7.1): 49.9 (6.6) Q2 (27.1–45.0): 50.3 (7.0) Q3 (45.1–67.4):50.7 (7.3) Q4 (≥67.5 ):51.3 (7.6)  Women  Q1 (<94.6): 46.7 (8.5) Q2 (94.6–124.5): 48.5 (8.5) Q3 (124.6–150.4): 48.9 (8.2) Q4 (≥150.5): 48.7 (7.8) | Type of measure: Self-reported: The EPIC physical activity questionnaire (EPIC-PAQ); face-to-face interview, and follow-up telephone survey Type of LTPA: recreational activities (walking, cycling, and sports); LTPA exposure: energy expenditure . MET intensity value (MET-h/week)  Categories of LTPA by Recreational physical activity (MET-h/week) <7.5 7.5–14.9 15.0–29.9 ≥30.0 | Men (MET-h/week, mean) Q1 (<7.1): 16.3 Q2 (27.1–45.0): 36.4 Q3 (45.1–67.4): 55.6 Q4 (≥67.5 ): 90.9  Women (MET-h/week, mean) Q1 (<94.6): 68.2 Q2 (94.6–124.5): 111.0 Q3 (124.6–150.4): 137.4 Q4 (≥150.5): 170.7  Categories of LTPA:  Men (person years) <7.5: 18.819 7.5–14.9; 26.500 15.0–29.9: 57.219 ≥30.0: 93.161  Women (person years) <7.5: 28.053 7.5–14.9; 60.131 15.0–29.9: 130.929 ≥30.0: 108.961 | Analysis 1: Recreational PA using multivariate Cox regression models  Analysis 2: Stratified by Interaction terms between PA and stratification variables were included in the models and interaction | participants without complete physical activity data in the two assessments including lost to follow-up, and further those with prevalent ischemic heart disease, stroke, cancer, or asthma at baseline  The final models were mutually adjusted for all PA variables considered, and also center, educational level, body mass index, waist and hip circumferences, baseline hypertension, hyperlipidemia, or diabetes, habit of smoking, alcohol consumption, energy intake, and Mediterranean diet score | Cancer mortality. CVD mortality. Overall mortality. | Cancer mortality: 758 deaths HR and 95% CI  Men:  426 deaths <7.5 MET-h/week (reference) Vs 7.5–14.9 MET-h/week - 0.96 (0.63–1.46)  15.0–29.9 MET-h/week - 0.96 (0.66–1.40) ≥30.0 MET-h/week - 0.86 (0.59–1.24) Women:  332 deaths <7.5 MET-h/week (reference) Vs 7.5–14.9 MET-h/week - 0.81 (0.50–1.29) 15.0–29.9 MET-h/week - 1.09 (0.72–1.65) ≥30.0 MET-h/week - 0.96 (0.62–1.47) Cardiovascular disease mortality : 283 deaths  HR and 95% CI  Men:  208 deaths <7.5 MET-h/week (reference) Vs 7.5–14.9 MET-h/week - 1.15 (0.64–2.06) 15.0–29.9 MET-h/week - 1.11 (0.65–1.91) ≥30.0 MET-h/week - 0.82 (0.47–1.43) Women:  75 deaths <7.5 MET-h/week (reference) Vs 7.5–14.9 MET-h/week - (1.09 0.46–2.58) 15.0–29.9 MET-h/week - 1.18 (0.54–2.60) ≥30.0 MET-h/week - 0.64 (0.26–1.58) Overall mortality: 1371 deaths HR and 95% CI  Men:  826 deaths <7.5 MET-h/week (reference) Vs 7.5–14.9 MET-h/week 1.06 (0.79–1.42) 15.0–29.9 MET-h/week - 0.96 (0.73–1.26) ≥30.0 MET-h/week - 0.96 (0.73–1.25) Women:  545 deaths <7.5 MET-h/week (reference) Vs 7.5–14.9 MET-h/week - 0.75 (0.54–1.05) 15.0–29.9 MET-h/week - 0.87 (0.65–1.16) ≥30.0 MET-h/week - 0.71 (0.52–0.98) |
| Ibsen et al., [39] | Denmark/Copenhagen and Aarhus | The Danish Diet, Cancer and Health cohort was established from December 1993 to May 1997. A total of 160725 men and women aged 50−64 years and resident in the greater municipal areas of Copenhagen and Aarhus. | 54276 participants/8860 deaths during a median follow-up of 17.0 years | Age mean: 56.1 (4.4) y % women: 52.1 | Type of measure: Self-reported: the lifestyle questionnaire Type of LTPA: walking, gardening, housework, home maintenance, biking, and sports LTPA exposure: hours per week Categories of LTPA by number of hours per week <30 minutes/day ≥30 minutes/day | Categories of LTPA: n(%) <30 minutes/day: 32757 (60.4) | Analysis 1: include PA in the multivariable model   First, given the low cut off for PA and the potential for underreporting, the dose−response relationship with all-cause mortality was modeled using restricted cubic spline variables with 4 knots. | Exclusion criteria for this study were cancer diagnoses before enrollment that had not been registered at the invitation to the study owing to delay in processing at the Danish Cancer Registry, diagnosis of stroke or myocardial infarction before baseline, or missing information on included variables.  The multivariable model included adherence to the modifiable lifestyle recommendations on diet, smoking, alcohol consumption, and physical activity. Each recommendation was adjusted for the other recommendations. | All-cause mortality. Cancer mortality.  CVD mortality. | All-cause mortality: 8860 deaths HR and 95% CI <30 minutes/day (reference) vs ≥30 minutes/day - 0.89 (0.85, 0.93) CVD mortality: 1,753 deaths HR and 95% CI (Model 3) <30 minutes/day (reference) vs ≥30 minutes/day - 0.85 (0.76, 0.94) Cancer mortality: 4,307 deaths HR and 95% CI  <30 minutes/day (reference) vs ≥30 minutes/day - 0.93 (0.88, 0.99) |
| Kikuchi et al., [40] | Japan | The Japan Public Health Center–based prospective study was started in 1990–1994, In brief, it targeted all registered Japanese inhabitants in 11 public health center areas who were 40–69 yr of age at the start of the baseline survey. | 83454 participants/8891 deaths during 894,718 person-years of follow-up, | Age mean:  Men: 61.5 (7.4) yr Women: 62.0 (7.6)  % women: 53.7 | Type of measure: Self-reported: survey questionnaire Type of LTPA: slow walking, brisk walking, moderate intensity of activity such as playing golf or gardening, and vigorous intensity of activity such as jogging or playing tennis. LTPA exposure: leisure time MVPA in MET-h/week Categories of LTPA by total:  Physically inactive: < 450 MET-h/week Physically active:≥ 450 MET-h/week  Categories of LTPA by VPA: 0% VPA  > 0% to e30% ≥ 30% | Categories of LTPA: Men (n): Physically inactive (< 450 MET-h/week): 27839 Physically active:≥ 450 MET-h/week : 10759 Women (n): Physically inactive (< 450 MET-h/week): 31351 Physically active:≥ 450 MET-h/week : 13505 Proportion of VPA to Total MVPA Volume: Men (n): 0% VPA : 4782 > 0% to ≤30%: 3937 ≥ 30% : 2040 Women (n): 0% VPA : 6300 > 0% to ≤30%: 4676 ≥ 30% : 2529 | The HR values of all-cause mortality were calculated for each physical activity group. Total MVPA volume included in multivariable models | Persons with non-Japanese nationality, duplicate enrollment, late report of emigration occurring before the start of the baseline study, and ineligibility owing to an incorrect birth date, persons who had died or moved out of Japan, subjects remained before the third follow-up survey, subjects with a history of cancer, or cardiovascular disease, and those with physical limitations, or missing data. Model 1: adjusting for age and residential area . Model 2: We then calculated the HR, adjusted for smoking, drinking, body mass index, diabetes history, and hypertension status. Model 3. adjusted for total MVPA volume (model 3). | All-cause mortality. | All- cause mortality: 8891 deaths HR and 95% CI (Model 3) Men Physically inactive (< 450 MET-h/week) (reference) Vs 0% - 0.75 (0.68–0.83) > 0% to e30% - 0.73 (0.65–0.82) ≥ 30% - 0.74 (0.62–0.89) Women Physically inactive (< 450 MET-h/week) (reference) Vs 0% - 0.71 (0.62–0.81) > 0% to e30% - 0.75 (0.64–0.88) ≥ 30% - 0.74 (0.58–0.94) |

**Supplemental Text 1: Individual participant data meta-analysis harmonization procedures**

**Physical activity questions**

| **UK Biobank** | **Health Survey for England and Scottish Health Survey** | **Taiwan Biobank** | **MJ Health Database** |
| --- | --- | --- | --- |
| In a typical week, on how many days did you do 10 minutes or more of moderate physical activities like carrying light loads, cycling at a normal pace? (open ended response)  How many minutes did you usually spend doing moderate activities on a typical day? (open ended response) | Leisure time moderate intensity activities such as light housework and gardening).  For each activity, participants are then asked to list the frequency over the last 7 days. For each frequency, participants are asked to provide the duration | Participants are asked to list the moderate activities they do such as:  Brisk walking, swimming, gymnastics, badminton, aerobic dance, resistance training (multiple choice)  Participants are then asked how many times/sessions a month they do these activities (open ended response)  Participants are asked how many minutes they spend doing the activities each session (open ended response) | What kind of moderate physical activities do you usually do? Such as daning, brisk walking, aerobics , badminton, aerobics, volleyball, gymnastics (multiple choice)  How much time do you devote to each activity in the last four weeks (multiple choice): <1 hour a week; 1-2 hours a week, 3-4 hours a week, 5-6 hours a week, over 7 hours a week  How often did you do each activity during the last four weeks? (multiple choice) 2-3 times/day, once a day, once every 2-3 days, once a week, none or rarely |
| In a typical week, on how many days did you do 10 minutes or more of walking (open ended response)  How many minutes did you usually spend walking on a typical day? (open ended response) | For each walking activity, participants are then asked to list the frequency. For each frequency, participants are asked to provide the duration, and intensity | Participants are asked to list the vigorous activities they do such as: Jogging, rope jumping, soccer, tennis, basketball, other ball sports, cycling, mountain climbing, stair climbing. (multiple choice)  Participants are then asked how many times/sessions a month they do these activities (open ended response)  Participants are asked how many minutes they spend doing the activities each session (open ended response) | What kind of vigorous physical activities do you usually do? Such as jogging, mountain climbing, climbing stairs, swimming, exercise, jump rope, rowing (multiple choice)  How much time do you devote to each activity in the last four weeks (multiple choice): <1 hour a week; 1-2 hours a week, 3-4 hours a week, 5-6 hours a week, over 7 hours a week  How often did you do each activity during the last four weeks? (multiple choice) 2-3 times/day, once a day, once every 2-3 days, once a week, none or rarely |
| In a typical week, on how many days did you do 10 minutes of vigorous moderate physical activities like that make you sweat or breathe hard  How many minutes did you usually spend doing vigorous activities on a typical day? (open ended response) | Leisure time vigorous activities. For each vigorous activity, participants are then asked to list the frequency over the last 7 days. For each frequency, participants are asked to provide the duration |  |  |
| How many times in the last 4 weeks did you do exercises such as swimming and cycling? (open ended response)  Each time you did exercises, how long did you spend doing it(open ended response) | For each sports and exercise activity, participants are then asked to list the frequency over the last 7 days. For each frequency, participants are asked to provide the duration |  |  |
| How many time in the last 4 weeks did you do strenuous sports? (open ended response)  Each time you did strenuous sports, how long did you spend doing it (open ended response) |  |  |  |

**Physical activity harmonisation**

| **Intensity** | **UK Biobank** | **Health Survey for England and Scottish Health Survey** | **Taiwan Biobank** | **MJ Health Database** |
| --- | --- | --- | --- | --- |
| Moderate | Duration and frequency of walking, DIY, and moderate activities were multiplied together and summed to get total weekly minutes. This was then divided by 7 to get minutes/day | Duration and frequency of each selected moderate intensity activities were multiplied together and summed to get total weekly minutes. This was then divided by 7 to get minutes/day | Duration and frequency of each selected moderate intensity activities were multiplied together and summed to get total monthly minutes. This was then divided by 30 to get minutes/day | Duration and frequency of each selected moderate intensity activities were multiplied together and summed to get total monthly minutes. This was then divided by 30 to get minutes/day |
| Vigorous | Duration and frequency of sports, exercise, and vigorous activities were multiplied together and summed to get total weekly minutes. This was then divided by 7 to get minutes/day | Duration and frequency of each selected vigorous intensity activities were multiplied together and summed to get total weekly minutes. This was then divided by 7 to get minutes/day | Duration and frequency of each selected vigorous intensity activities were multiplied together and summed to get total monthly minutes. This was then divided by 30 to get minutes/day | Duration and frequency of each selected vigorous intensity activities were multiplied together and summed to get total monthly minutes. This was then divided by 30 to get minutes/day |

**Covariate harmonisation**

|  | UK Biobank | Health Survey for England and Scottish Health Survey | Taiwan Biobank | MJ Health Database |
| --- | --- | --- | --- | --- |
| Age | Calculated from date of birth and date of assessment centre visit | | | |
| Sex | Participants in each cohort reported their sex as Male/Female | | | |
| Smoking history | Participants in each cohort reported their smoking status as Current/Former/Never | | | |
| Alcohol consumption | Participants reported the frequency of their alcohol intake over the past month. Those who reported quit were classified as Former, those who reported <1 drink/week were classified as Seldom. All other were classified as Current | Participants reported the units of alcohol in the past 7 days. Those who reported quit were classified as Former, and those who reported ≤8g of ethanol (equivalent to 1 standard drink) were classified as Seldom. All other were classified as Current | Participants reported their alcohol habits as: Quit (classified as Former in the current study), Seldom, and Current | Participants reported how many drinks they consumed in the past week as: Quit (classified as Former in the current study), ≤1 glass (classified as Seldom). All other were classified as Current |
| Body mass index | Measured at assessment centre: Height in cm and weight in kg to calculate kg/m^2^ | | | |
| Education | All participants reported their highest attained education level. This was then recategorized as college/university level and higher or lower (binary 1/0) | | | |
| Prevalent CVD | Self-reported and health linkage with hospitalisation records | | | |
| Prevalent cancer | Self-reported and health linkage with hospitalisation records | | | |

**Supplemental Table 2: Individual participant data meta-analysis baseline characteristics by cohort**

|  | **UK Biobank** | **MJ Cohort** | **HSE/SHS** | **Taiwan Biobank** | **Overall** |
| --- | --- | --- | --- | --- | --- |
| **Sample** | 403,153 | 363,514 | 97,998 | 100,258 | 964,923 |
| **Follow up, yrs** | 12.5 (1.6) | 14.7 (5.1) | 9.5 (4.5) | 4.9 (2.0) | 12.2 (4.7) |
| **Age, yrs** | 56.2 (8.1) | 39.9 (12.7) | 46.9 (16.9) | 49.8 (10.9) | 48.5 (13.5) |
| Range | 38.0 – 73.0 | 18.0 – 98.0 | 18.0 – 97.0 | 30.0 – 70.0 | 18.0 – 98.0 |
| **Male (%)** | 190,491 (47.3) | 177,864 (48.9) | 44,440 (45.3) | 64,225 (64.1) | 477,020 (49.4) |
| **Moderate intensity, mins/day** | 45.9 (36.6) | 13.3 (18.1) | 62.4 (48.5) | 9.4 (18.5) | 42.3 (67.7) |
| **Vigorous intensity, mins/day** | 21.3 (20.6) | 2.3 (8.8) | 12.1 (20.6) | 3.1 (9.8) | 14.9 (28.4) |
| **Moderate-vigorous physical activity, mins/day** | 70.4 (50.7) | 15.8 (21.5) | 84.5 (60.9) | 13.0 (23.5) | 57.2 (82.6) |
| **Smoking history (%)** |  |  |  |  |  |
| Former | 140,922 (35.0) | 21,997 (6.1) | 24,172 (24.7) | 10,013 (10.0) | 197,104 (20.4) |
| Never | 221,588 (55.0) | 261,763 (72.0) | 47,425 (48.4) | 80,821 (80.6) | 611,597 (63.4) |
| Current | 40,643 (10.1) | 79,754 (21.9) | 26,401 (26.9) | 9,424 (9.4) | 156,222 (16.2) |
| **Alcohol consumption history (%)** |  |  |  |  |  |
| Former | 13,849 (3.4) | 9,938 (2.7) | 3,838 (3.9) | 2,499 (2.5) | 30,124 (3.1) |
| Seldom^1^ | 15,813 (3.9) | 22,044 (6.1) | 7,647 (7.8) | 5,920 (5.9) | 51,424 (5.3) |
| Current | 373,491 (92.6) | 331,532 (91.2) | 86,513 (88.3) | 91,839 (91.6) | 883,375 (91.5) |
| **Obese (%)^2^** | 94,123 (23.3) | 48,332 (13.3) | 21,157 (21.6) | 20,457 (20.4) | 184,069 (19.1) |
| **College/University (%)** | 256,336 (63.6) | 219,957 (60.5) | 35,515 (36.2) | 56,859 (56.7) | 568,667 (58.9) |
| **Prevalent CVD (%)** | 25,134 (6.2) | 14,693 (4.0) | 5,256 (5.4) | 1,412 (1.4) | 46,495 (4.8) |
| **Prevalent cancer (%)** | 13,984 (3.5) | 5,536 (1.5) | 1,450 (1.5) | 858 (0.9) | 21,828 (2.3) |
| **Biomarkers** |  |  |  |  |  |
| High density lipoprotein, mmol/L | 1.4 (0.4) | 1.5 (0.4) | 1.5 (0.4) | 1.5 (0.4) | 1.5 (0.4) |
| Low density lipoprotein, mmol/L | 3.6 (0.9) | 3.3 (0.9) | 3.4 (1.0) | 3.4 (0.9) | 3.4 (0.9) |
| Triglycerides, mmol/L | 1.7 (1.0) | 3.2 (2.8) | 1.6 (1.2) | 3.2 (2.6) | 2.5 (2.3) |
| **Blood pressure** |  |  |  |  |  |
| Systolic | 139.3 (19.6) | 123.7 (22.5) | 131.5 (19.2) | 118.6 (17.9) | 134.2 (21.0) |
| Diastolic | 82.1 (10.7) | 74.6 (14.3) | 74.0 (11.8) | 72.9 (11.0) | 79.3 (11.7) |
| **Self-rated health (%)** |  |  |  |  |  |
| Excellent | 71,478 (17.8) | 4,138 (5.8) | 32,344 (33.0) | - | 107,960 (18.9) |
| Good | 234,695 (58.4) | 4,138 (5.8) | 59,722 (61.0) | - | 350,063 (61.3) |
| Fair | 79,384 (19.7) | 4,138 (5.8) | 4,722 (4.8) | - | 94,200 (16.5) |
| Poor | 16,439 (4.1) | 4,138 (5.8) | 1,195 (1.2) | - | 18,891 (3.3) |
| **Event rate (%)** |  |  |  |  |  |
| All-cause mortality | 6.6 | 6.8 | 8.2 | 0.7 | 6.2 |
| CVD | 1.6 | 1.0 | 2.0 | 0.1 | 1.3 |
| Cancer | 3.1 | 2.5 | 2.3 | 0.3 | 2.5 |

**Supplemental Figure 1: Funnel Plots**

1a) Meta-analysis of high vs low moderate intensity leisure time-physical activity and mortality: a) all-cause mortality; b) cardiovascular mortality; c) cancer mortality.

**a)**

| 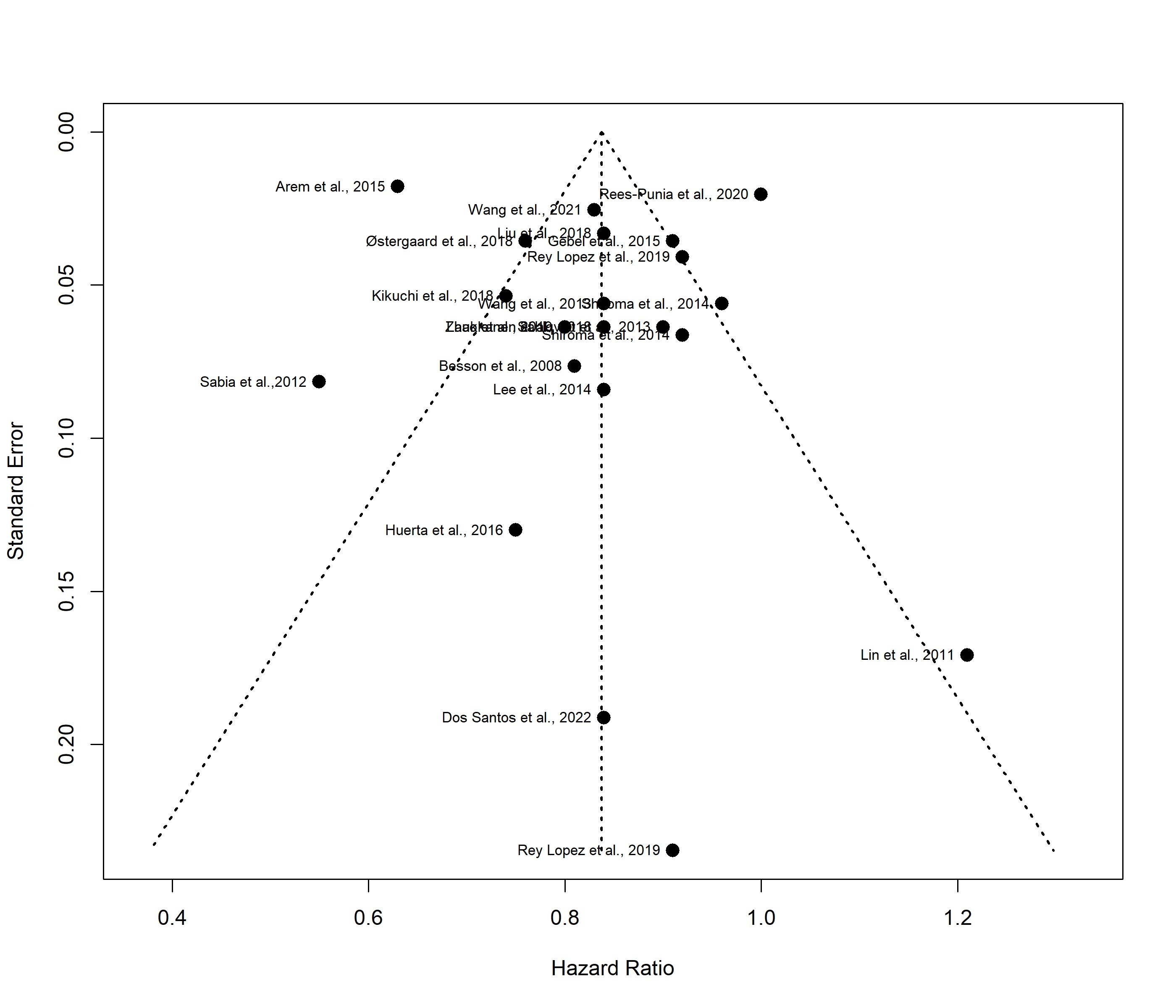 | 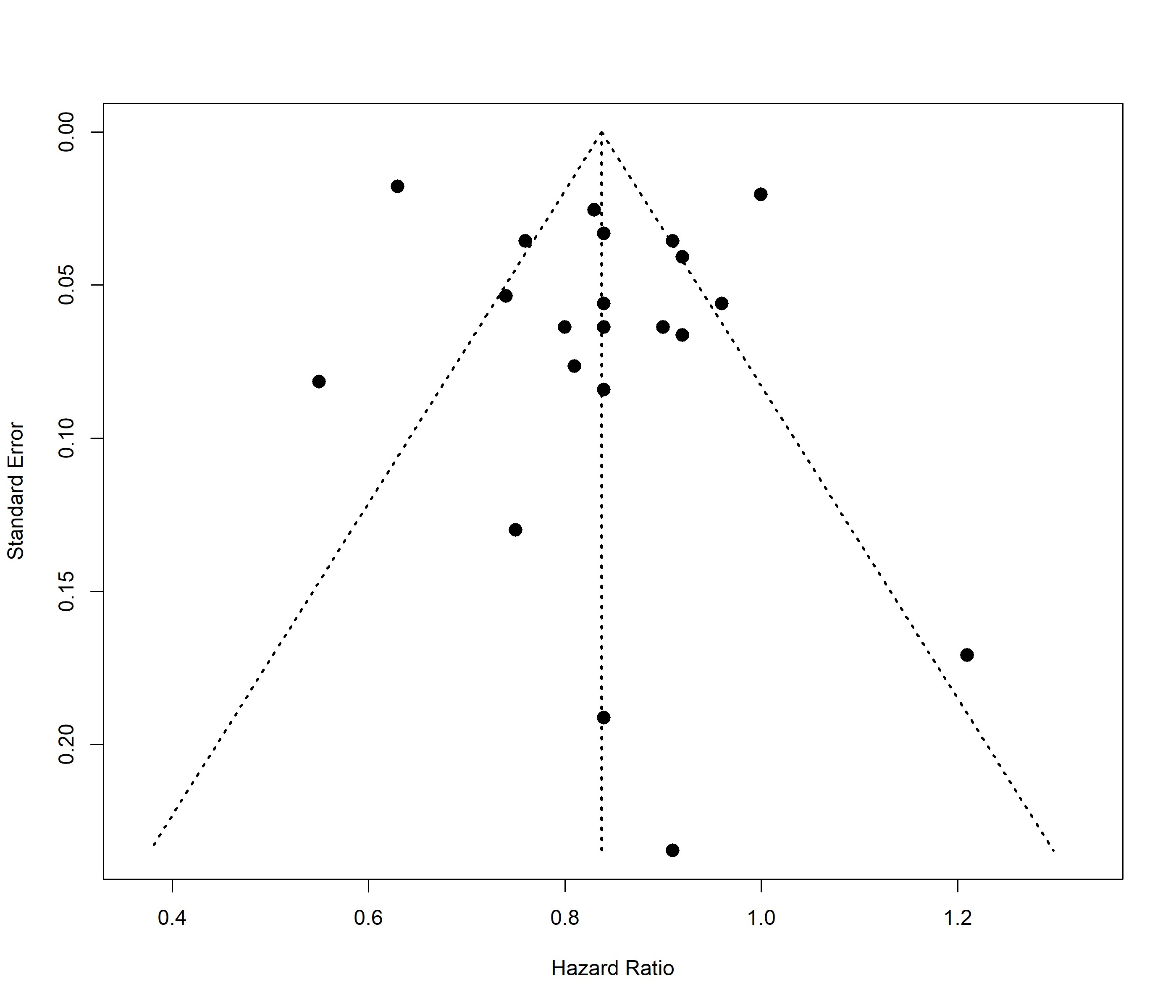 |
| --- | --- |

**b)**

| 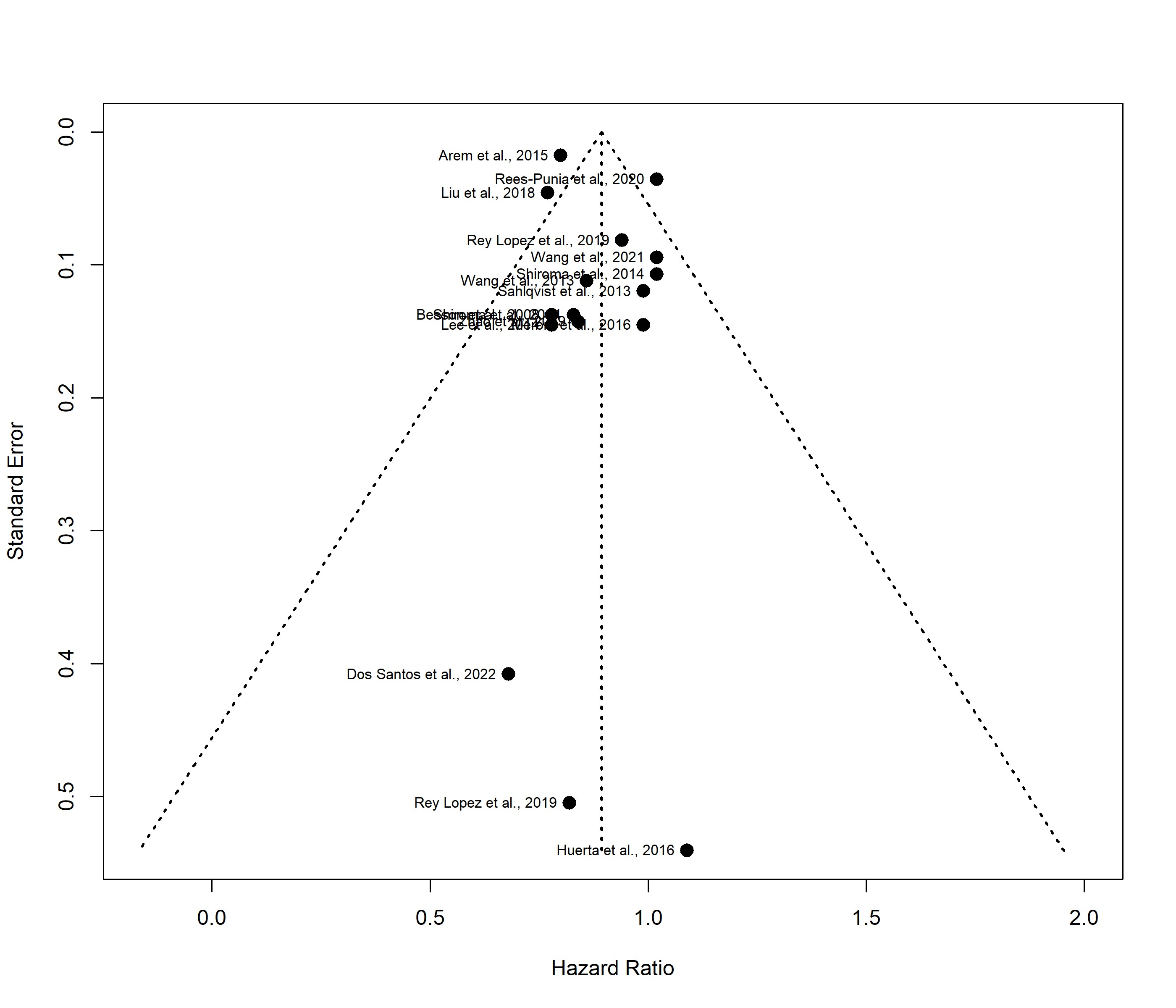 | 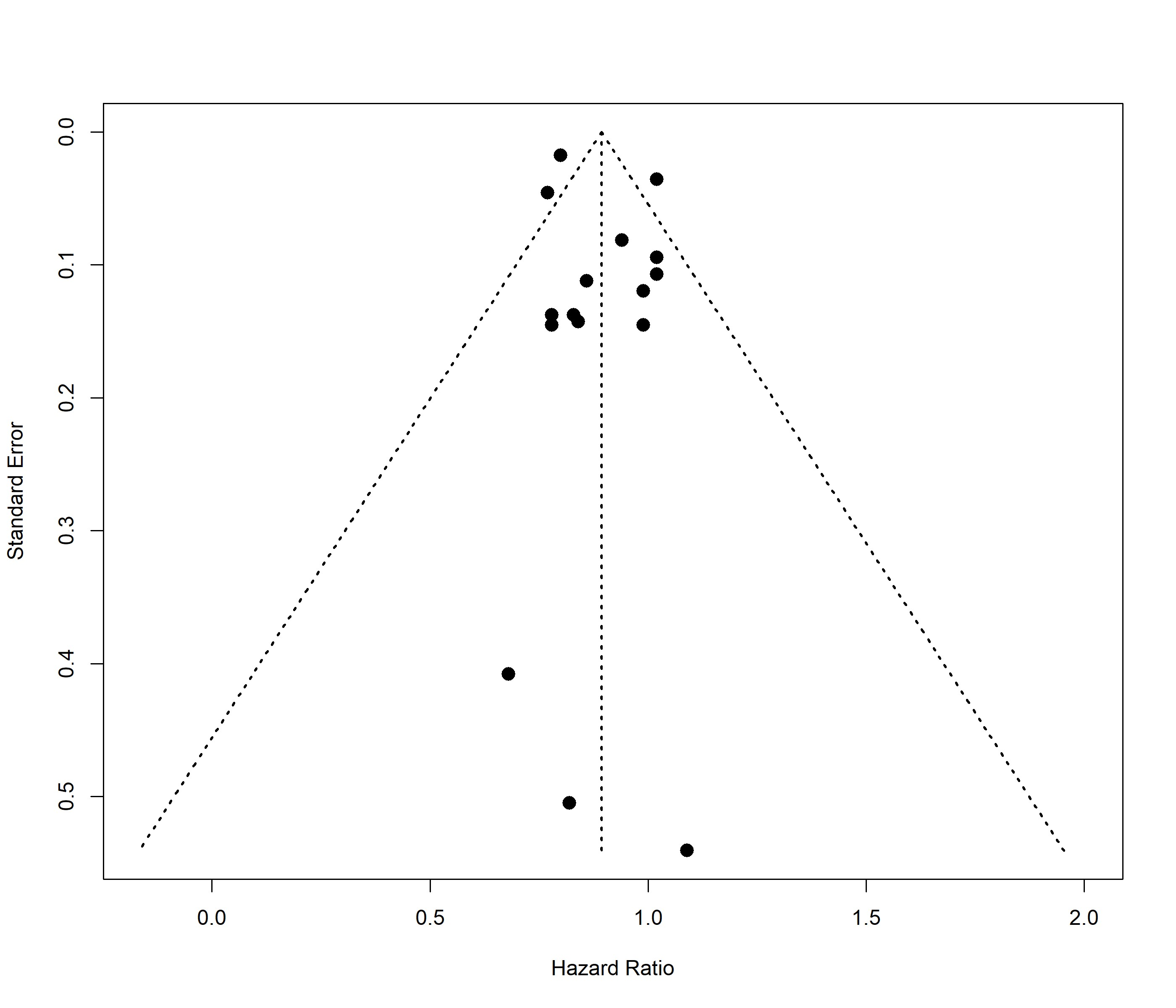 |
| --- | --- |

**c)**

| 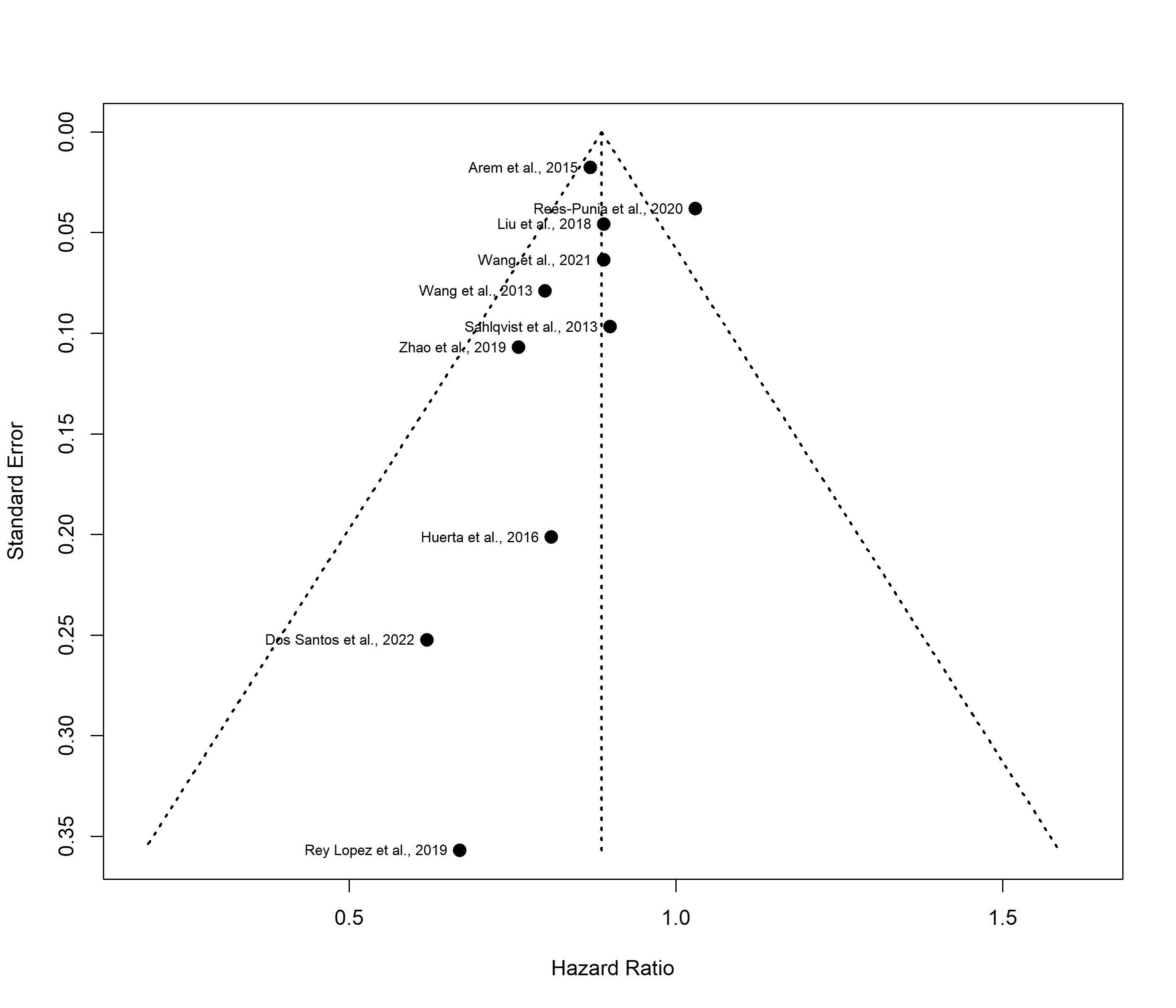 | 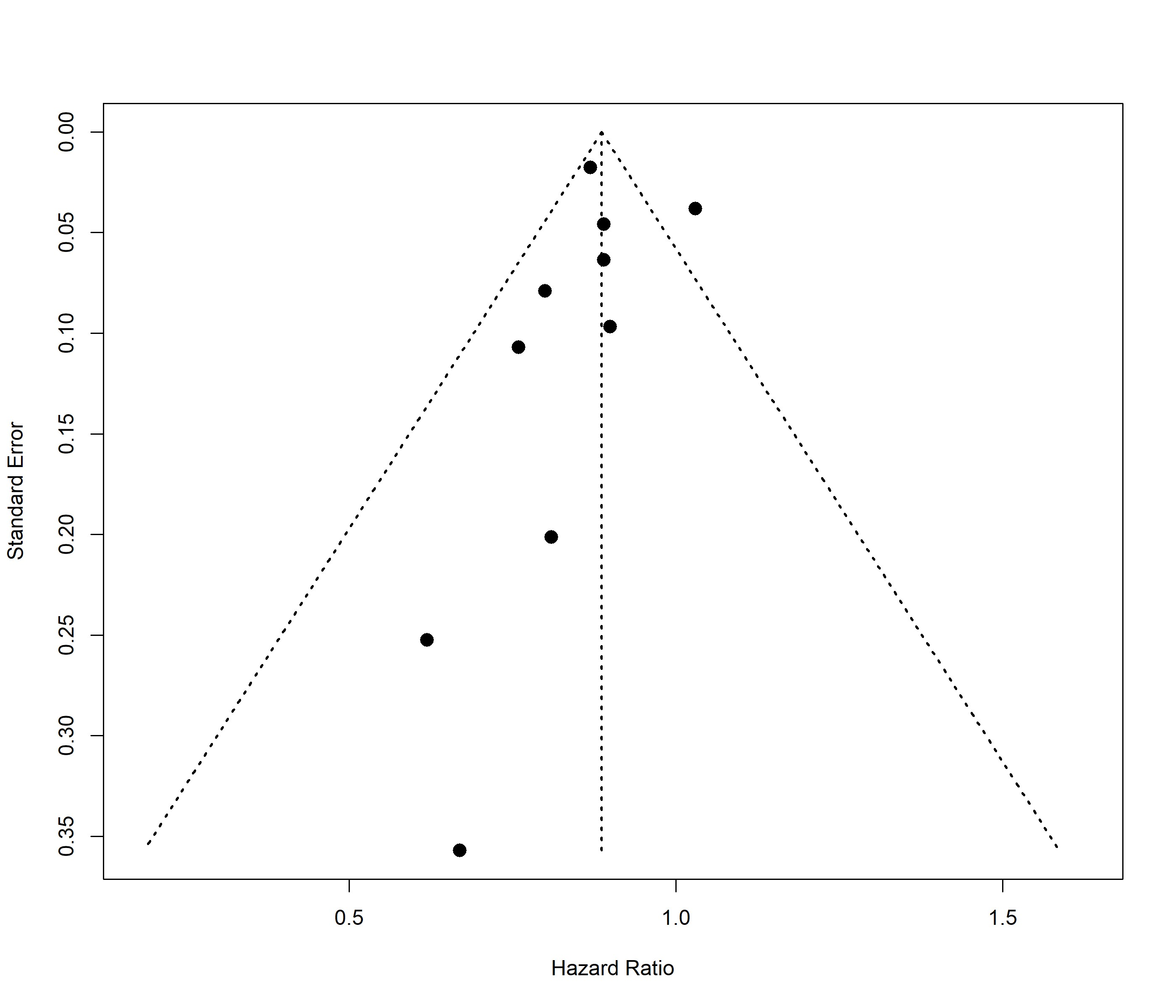 |
| --- | --- |

1b) Meta-analysis of high vs low vigorous intensity of leisure time-physical activity and mortality: a) all-cause mortality; b) cardiovascular mortality; c) cancer mortality.

**a)**

| 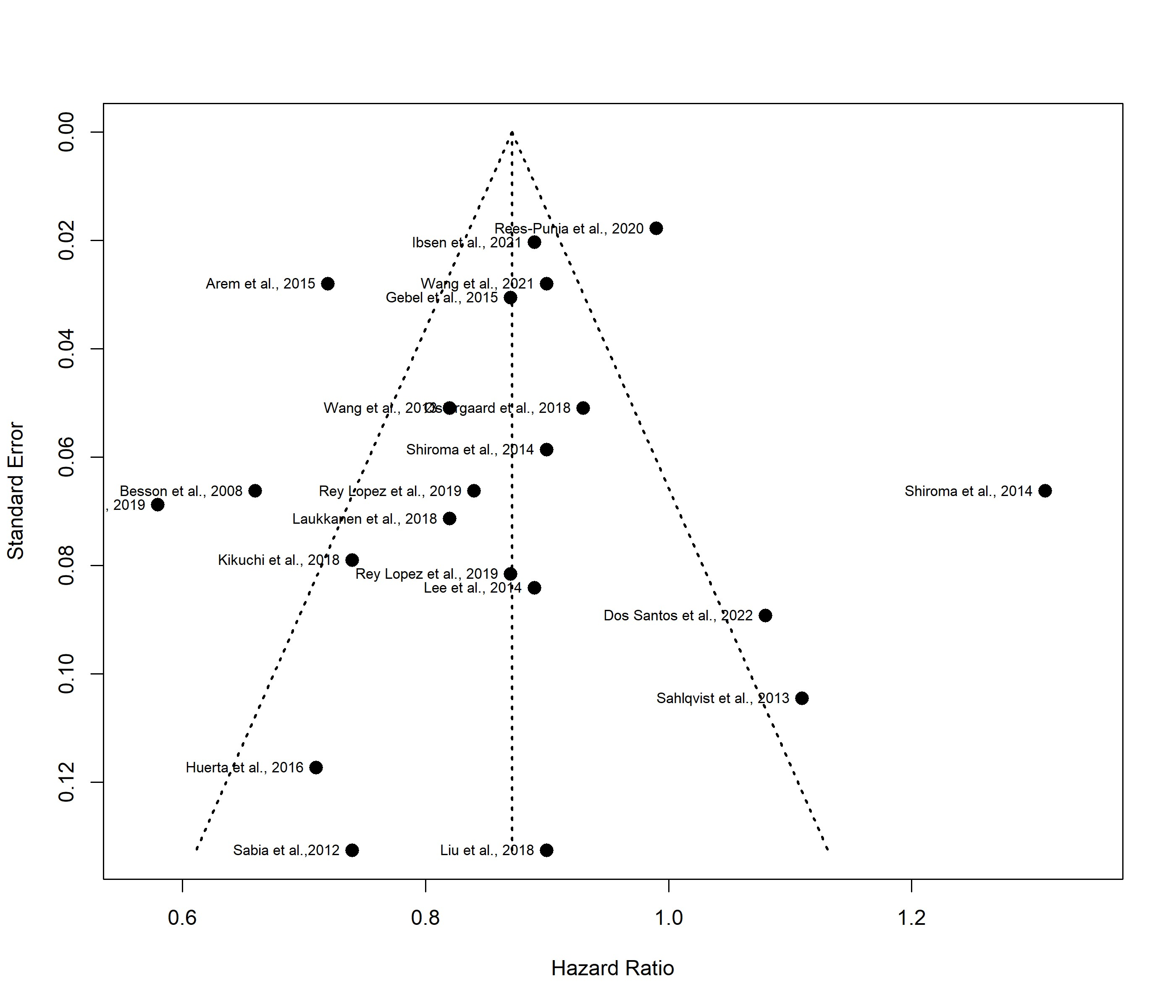 | 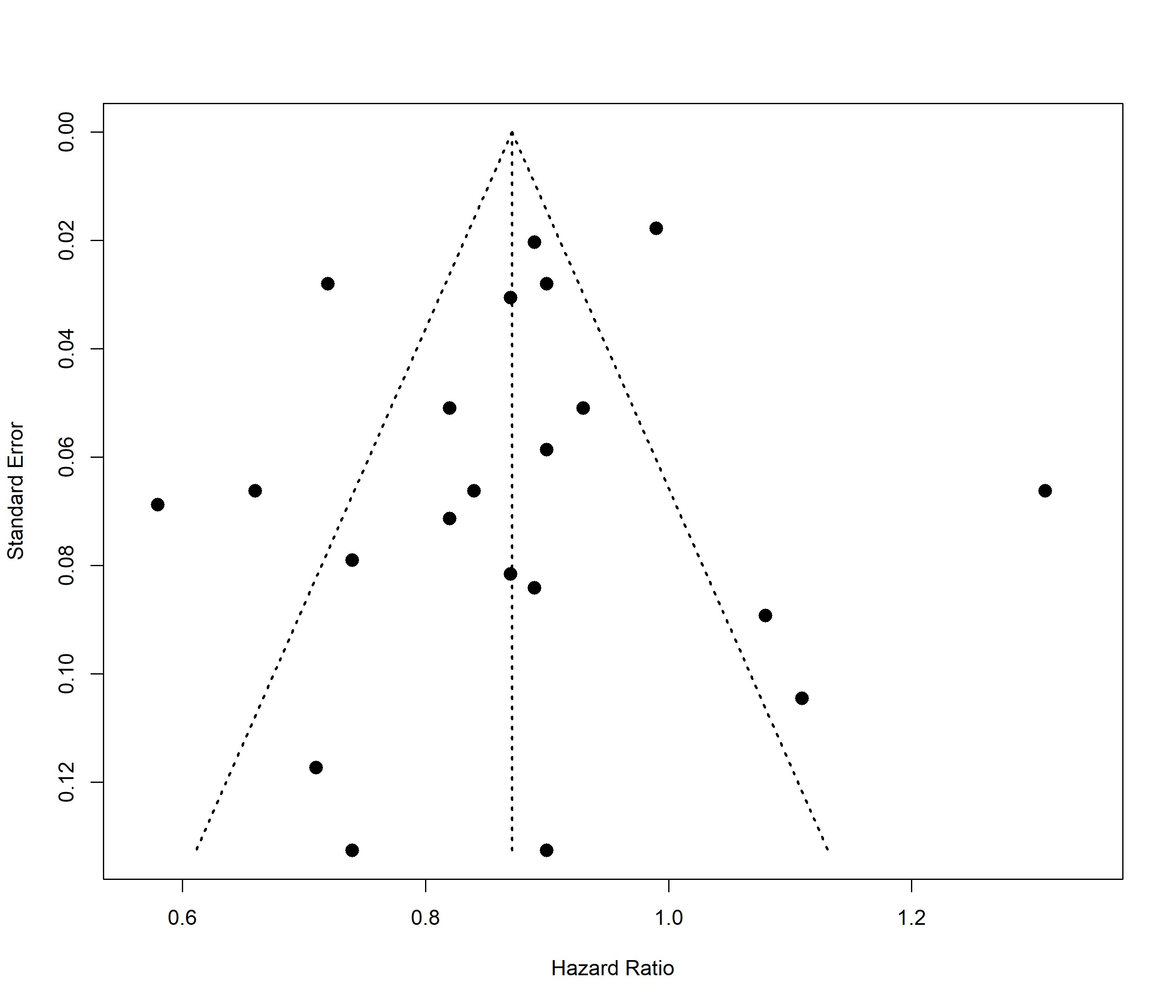 |
| --- | --- |

**b)**

| 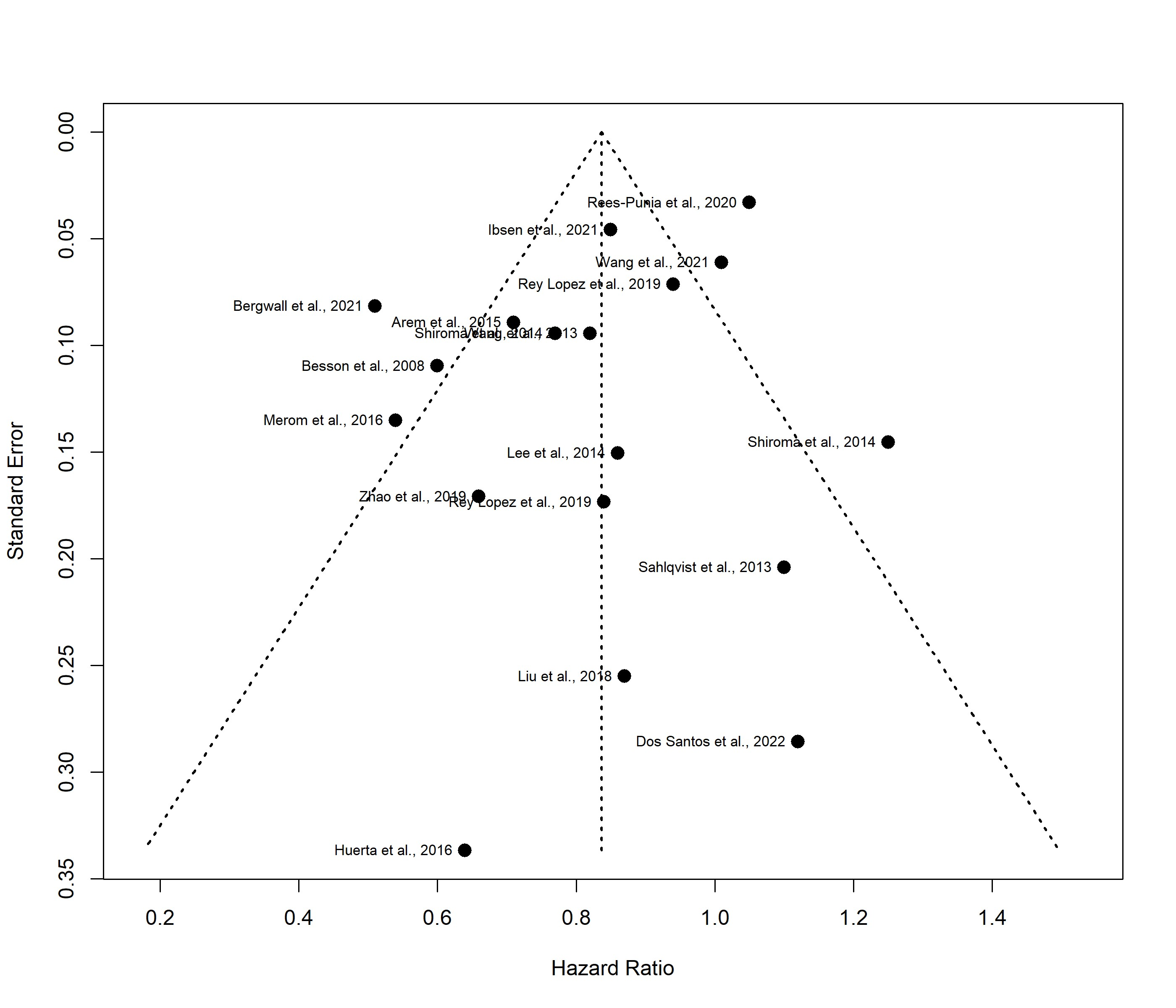 | 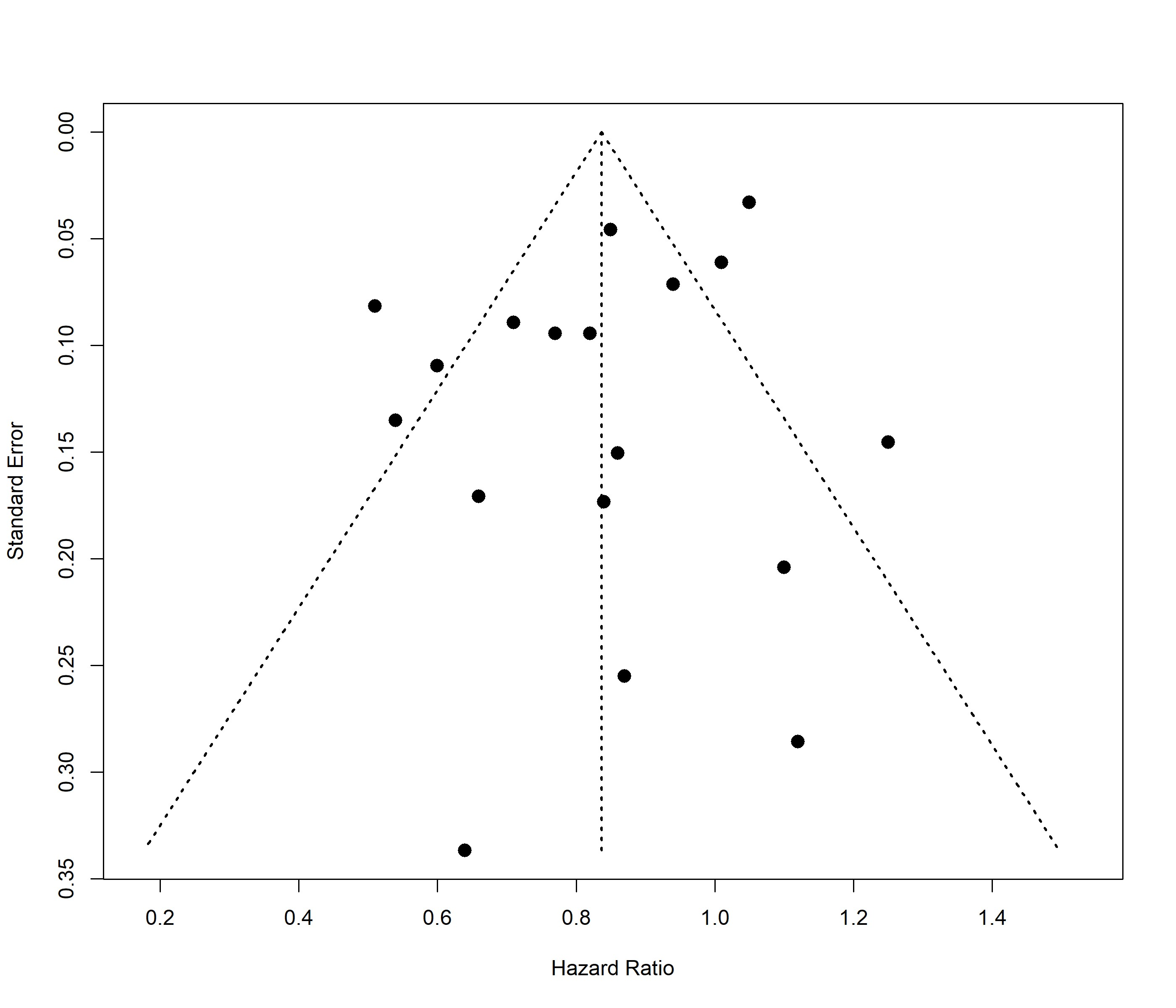 |
| --- | --- |

**c)**

| 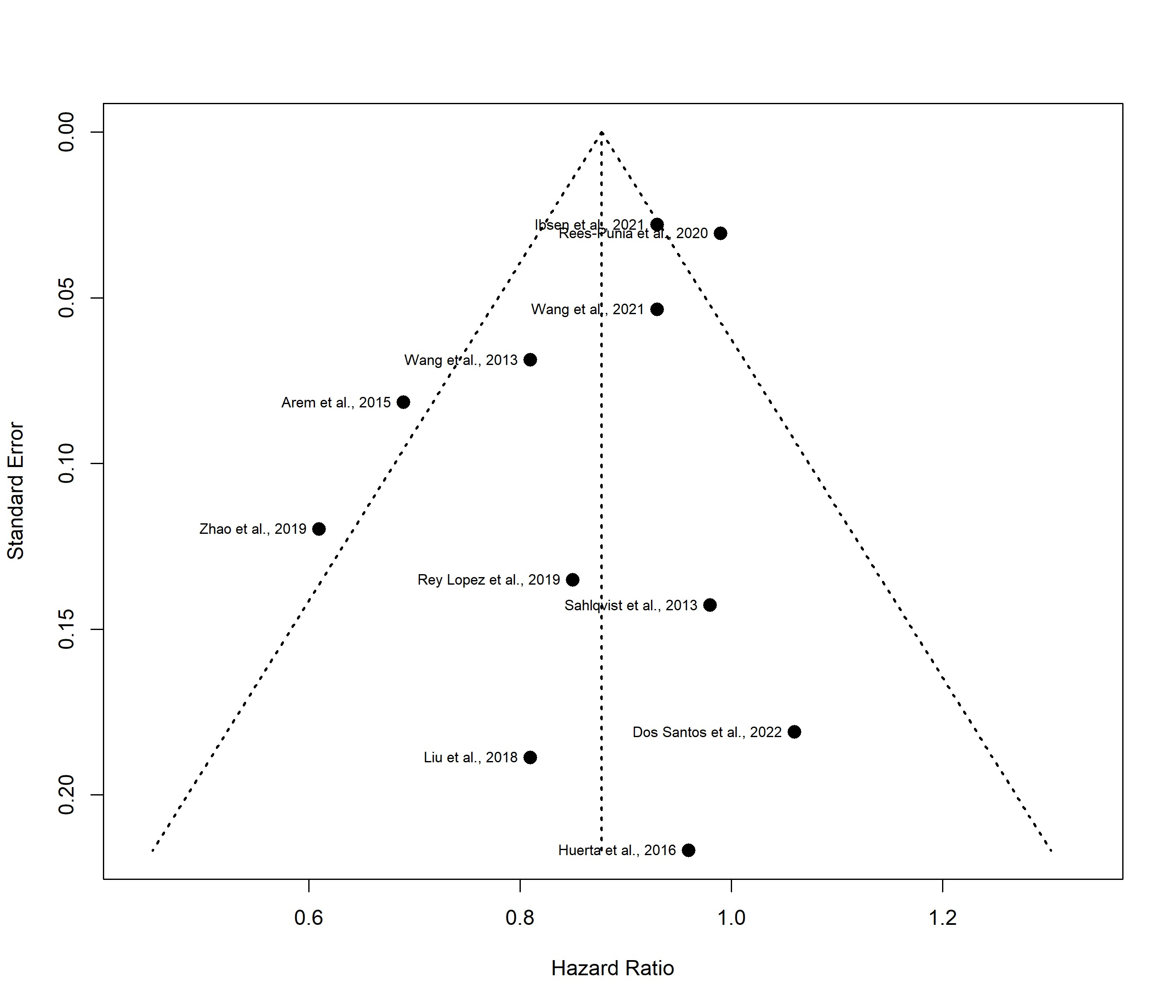 | 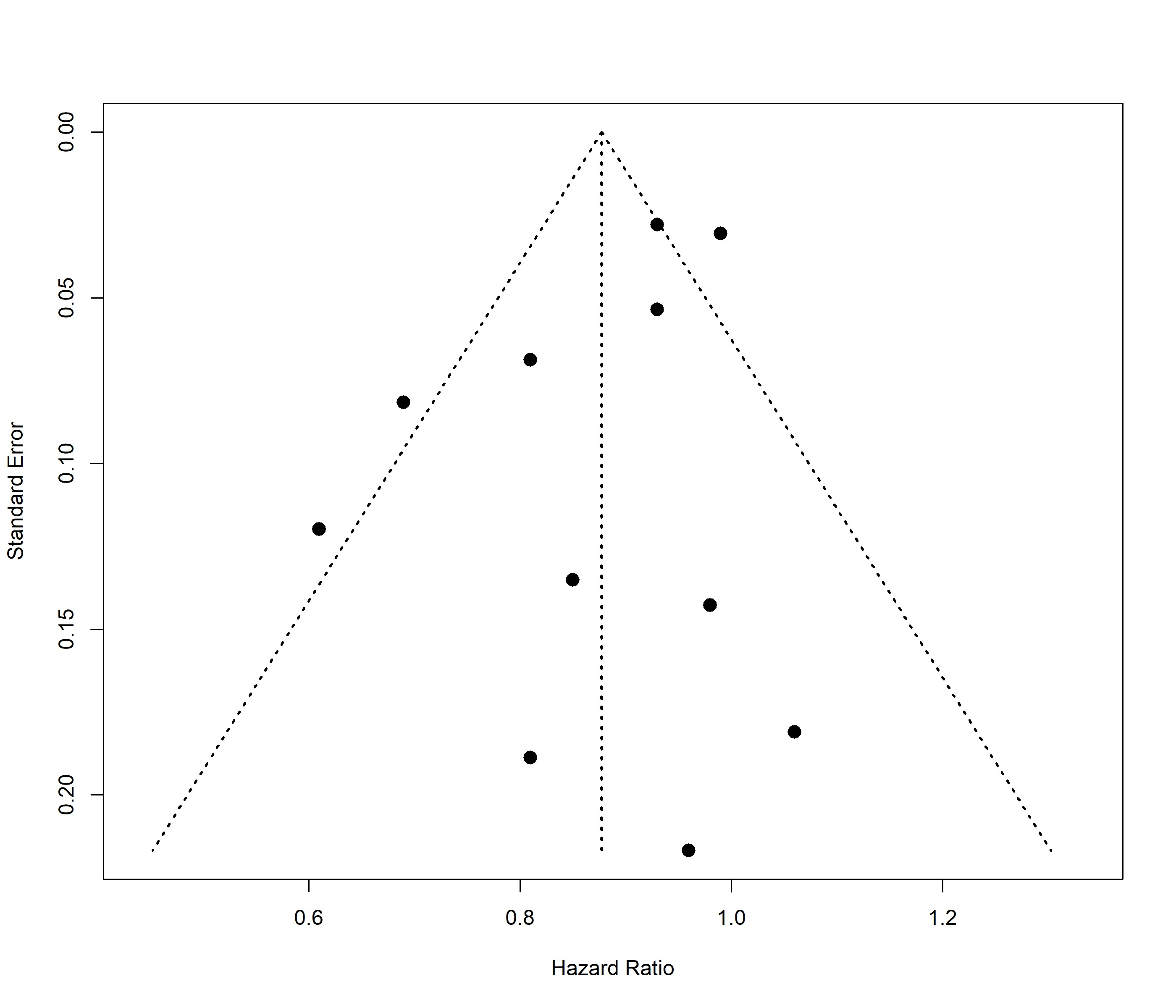 |
| --- | --- |

**Supplemental Figure 2: Aggregated meta-analysis results after exclusion of unpublished IPD estimates**

A) **Moderate intensity and all-cause mortality**

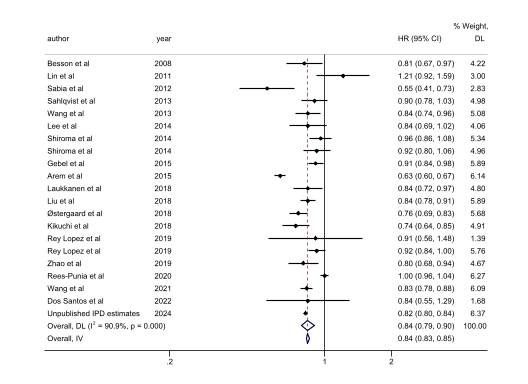

B) **Vigorous intensity and all-cause mortality**

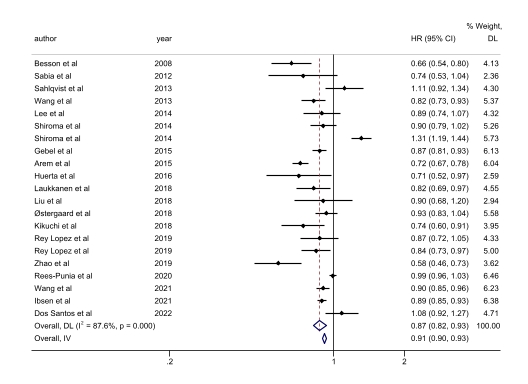

C) **Moderate intensity and cardiovascular disease mortality**

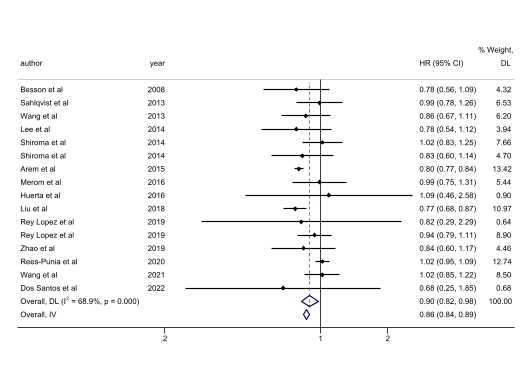

D) **Vigorous intensity and cardiovascular disease mortality**

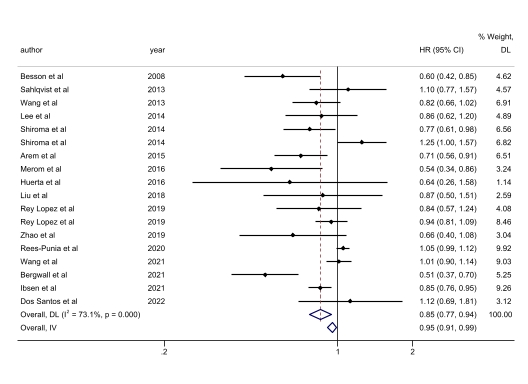

E) **Moderate intensity and cancer mortality**

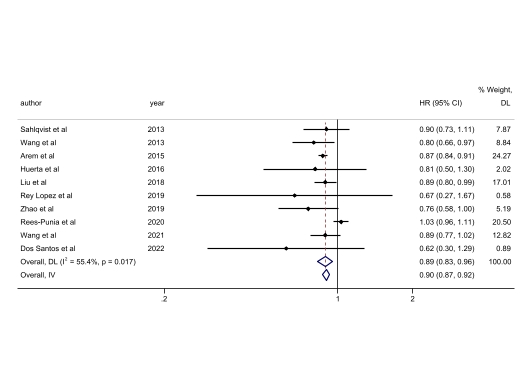

F) **Vigorous intensity and cancer mortality**

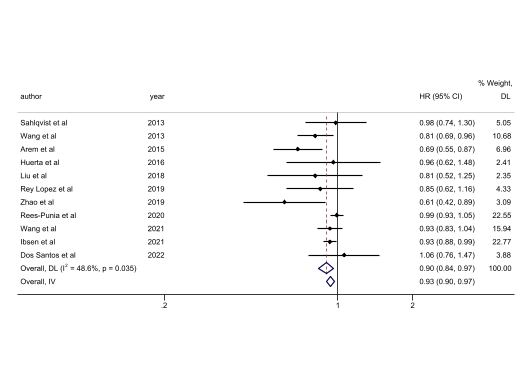

**Figure 3a: Leave-one-out sensitivity analysis:** Meta-analysis of high vs low MODERATE INTENSITY leisure time-physical activity and ALL-CAUSE MORTALITY

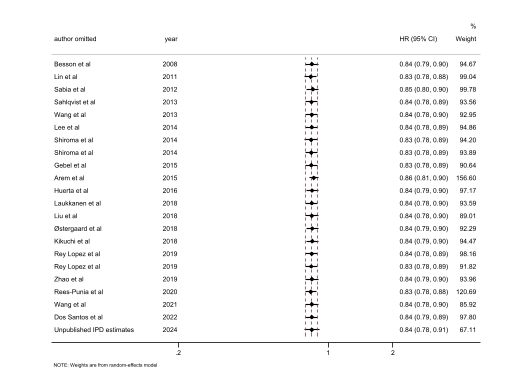

**Figure 3b: Leave-one-out sensitivity analysis:** Meta-analysis of high vs low MODERATE INTENSITY leisure time-physical activity and CANCER MORTALITY

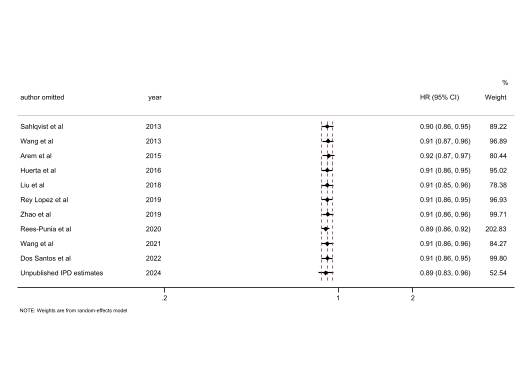

**Figure 3c: Leave-one-out sensitivity analysis**: Meta-analysis of high vs low MODERATE INTENSITY leisure time-physical activity and CARDIOVASCULAR DISEASE MORTALITY

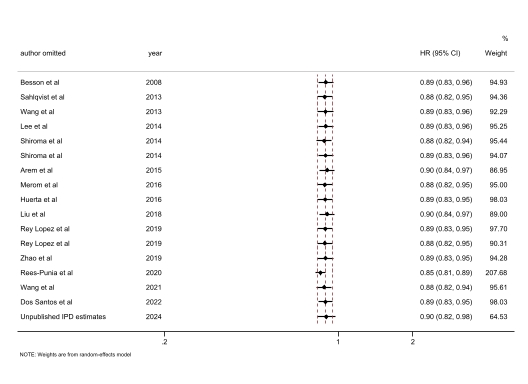

**Figure 3d: Leave-one-out sensitivity analysis**: Meta-analysis of high vs low VIGOROUS INTENSITY leisure time-physical activity and ALL-CAUSE MORTALITY

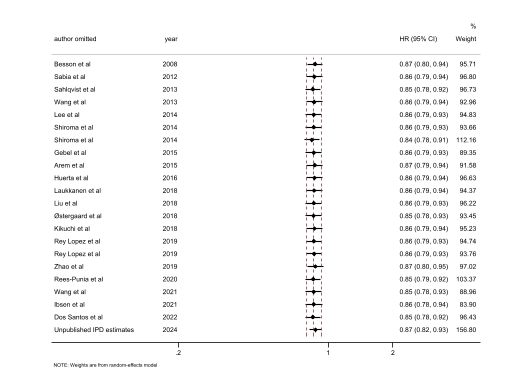

**Figure 3e: Leave-one-out sensitivity analysis**: Meta-analysis of high vs low VIGOROUS INTENSITY leisure time-physical activity and CANCER MORTALITY

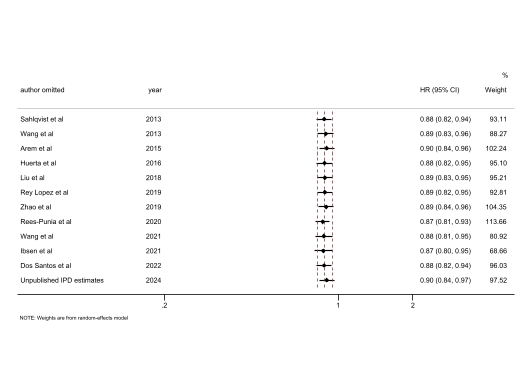

**Figure 3f: Leave-one-out sensitivity analysis**: Meta-analysis of high vs low VIGOROUS INTENSITY leisure time-physical activity and CARDIOVASCULAR DISEASE MORTALITY

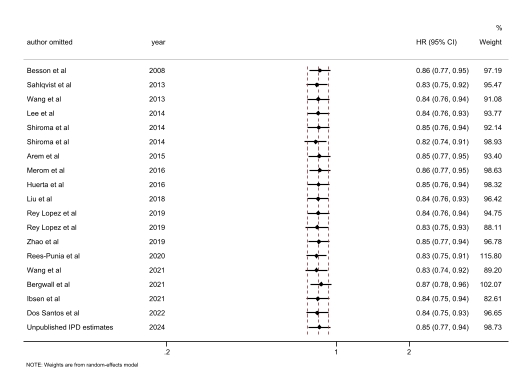

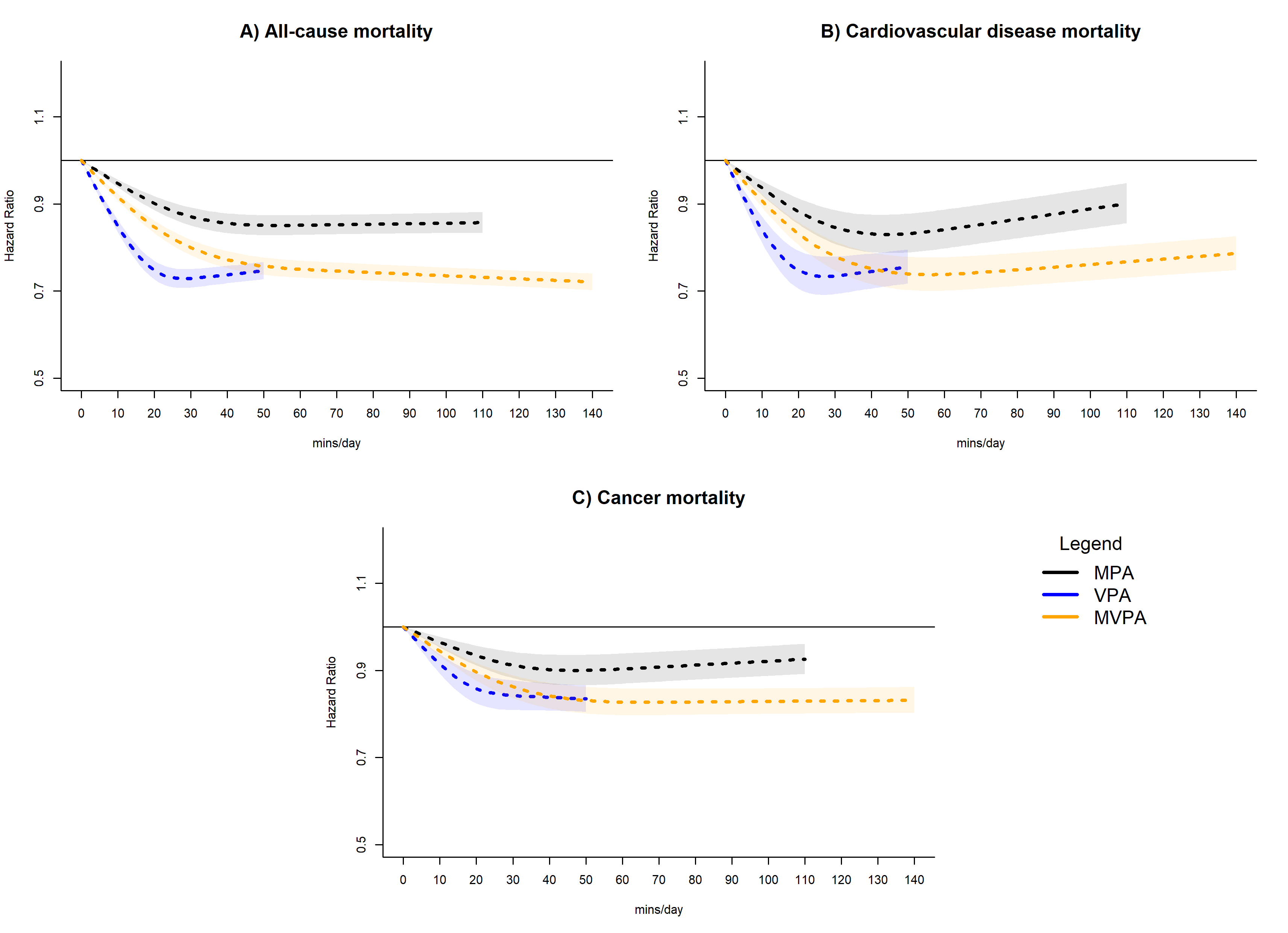
**Supplemental Figure 4: Dose-response association of physical activity intensity volume with all-cause, cardiovascular disease, and cancer mortality; exclusion of participants with <5 years of follow-up**

Adjusted for age, sex, smoking status, alcohol consumption, body mass index, education, prevalent cvd (for all-cause and cancer mortality), prevalent cancer (for all-cause and cardiovascular disease mortality)

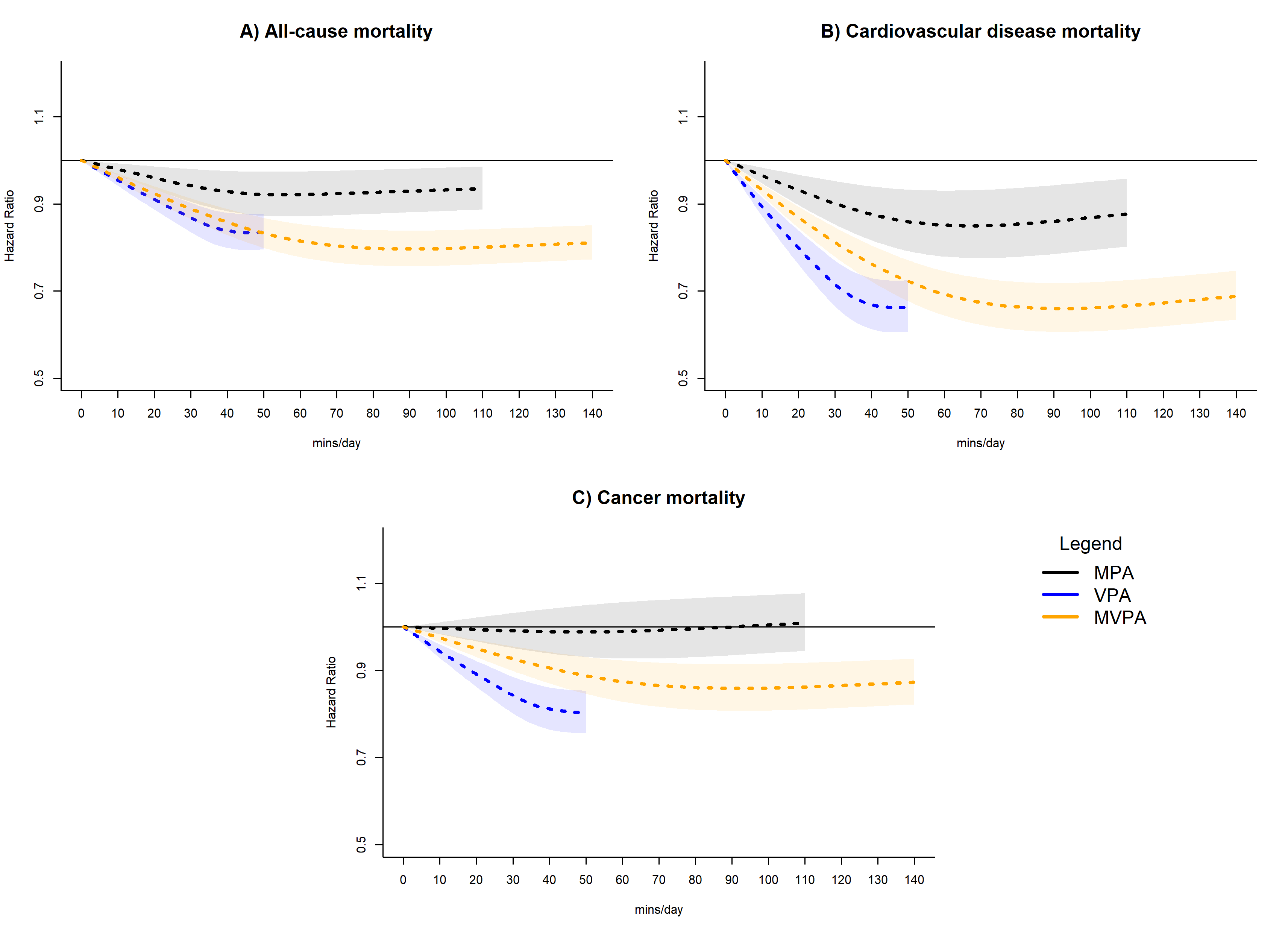
**Supplemental Figure 5: Dose-response association of physical activity intensity volume with all-cause, cardiovascular disease, and cancer mortality; exclusion of participants with poor or fair self-rated health**

Adjusted for age, sex, smoking status, alcohol consumption, body mass index, education, prevalent cvd (for all-cause and cancer mortality), prevalent cancer (for all-cause and cardiovascular disease mortality)

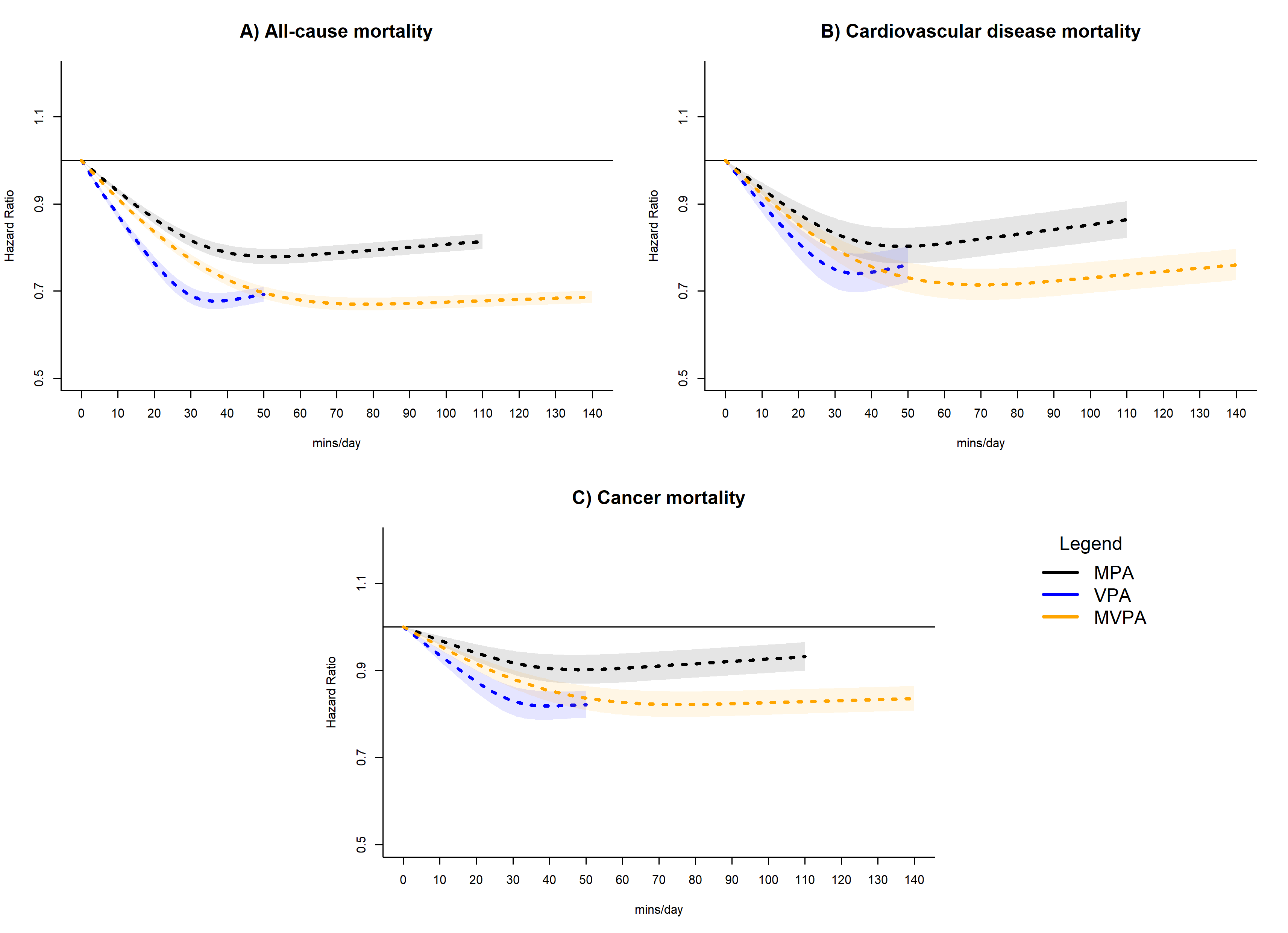
**Supplemental Figure 6: Dose-response association of physical activity intensity volume with all-cause, cardiovascular disease, and cancer mortality; exclusion of participants under 40 years of age**

Adjusted for age, sex, smoking status, alcohol consumption, body mass index, education, prevalent cvd (for all-cause and cancer mortality), prevalent cancer (for all-cause and cardiovascular disease mortality)

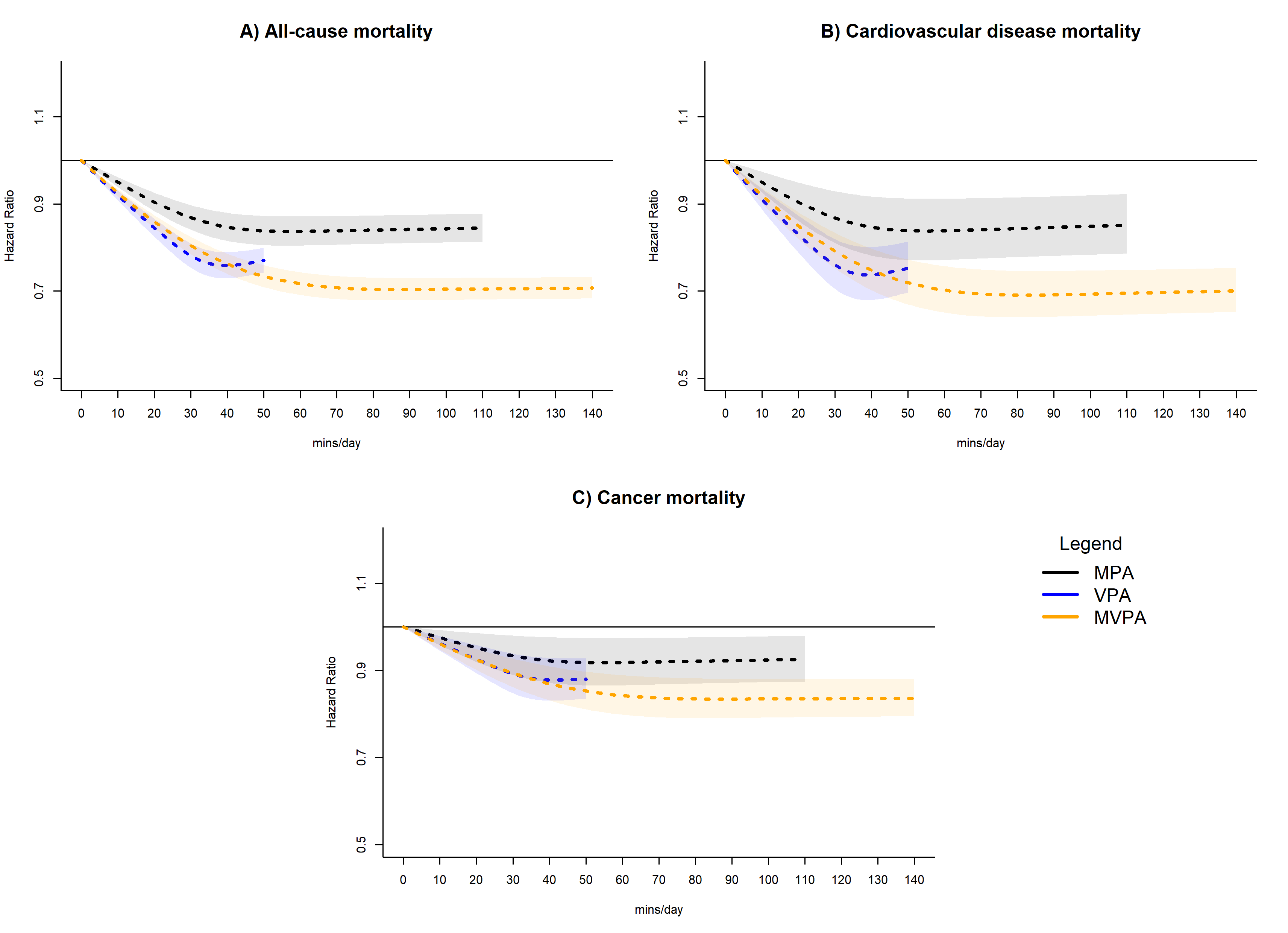
**Supplemental Figure 7: Dose-response association of physical activity intensity volume with all-cause, cardiovascular disease, and cancer mortality; adjustment for biomarkers**

Adjusted for age, sex, smoking status, alcohol consumption, body mass index, education, prevalent cvd (for all-cause and cancer mortality), prevalent cancer (for all-cause and cardiovascular disease mortality), high and low density lipoprotein, triglycerides, and diastolic and systolic blood pressure

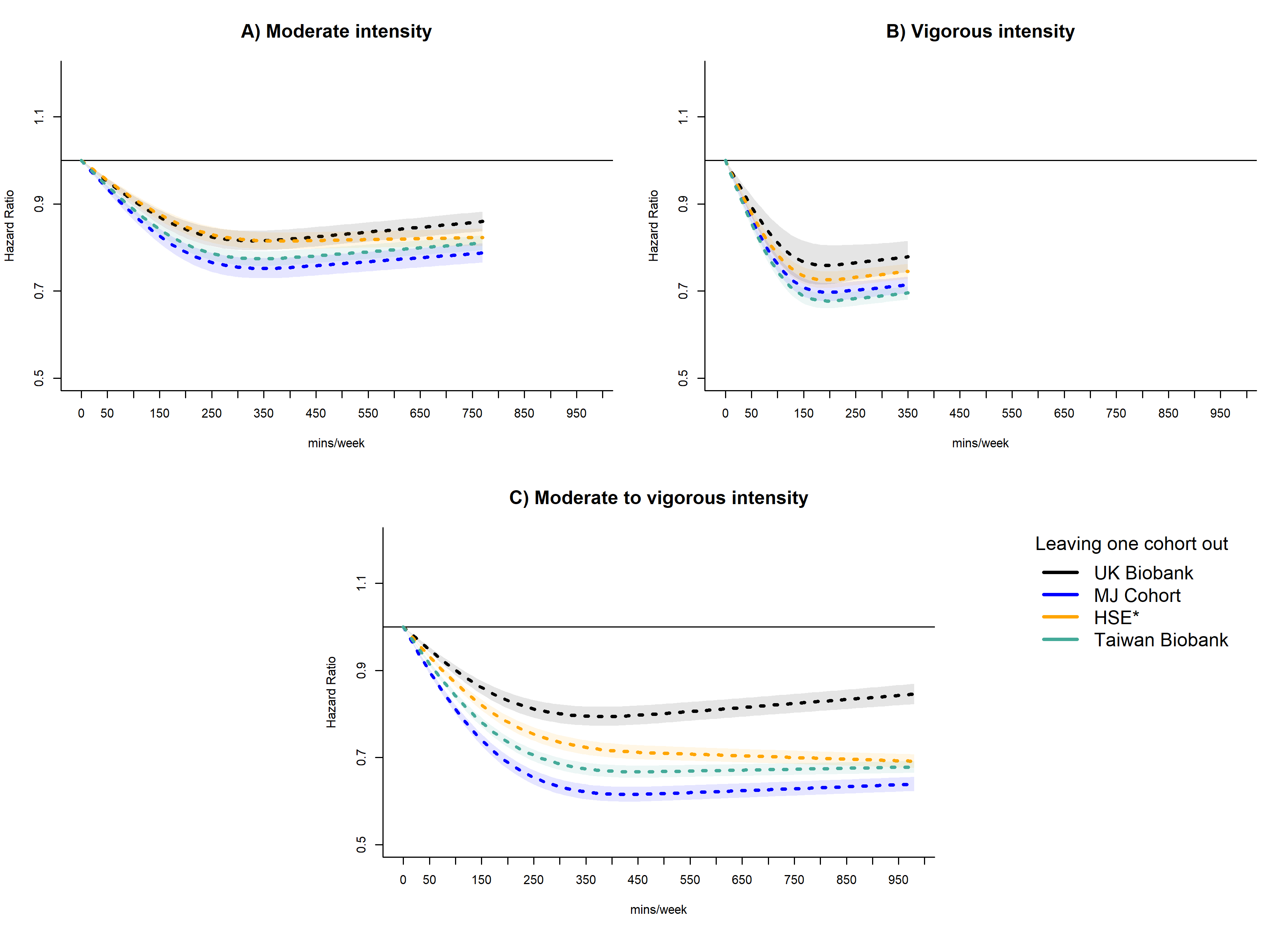
**Supplemental Figure 8: Leave-one-cohort-out dose response association with all-cause mortality**

Adjusted for age, sex, smoking status, alcohol consumption, body mass index, education, prevalent cvd and cancer. *Health Survey for England and Scottish Health Survey

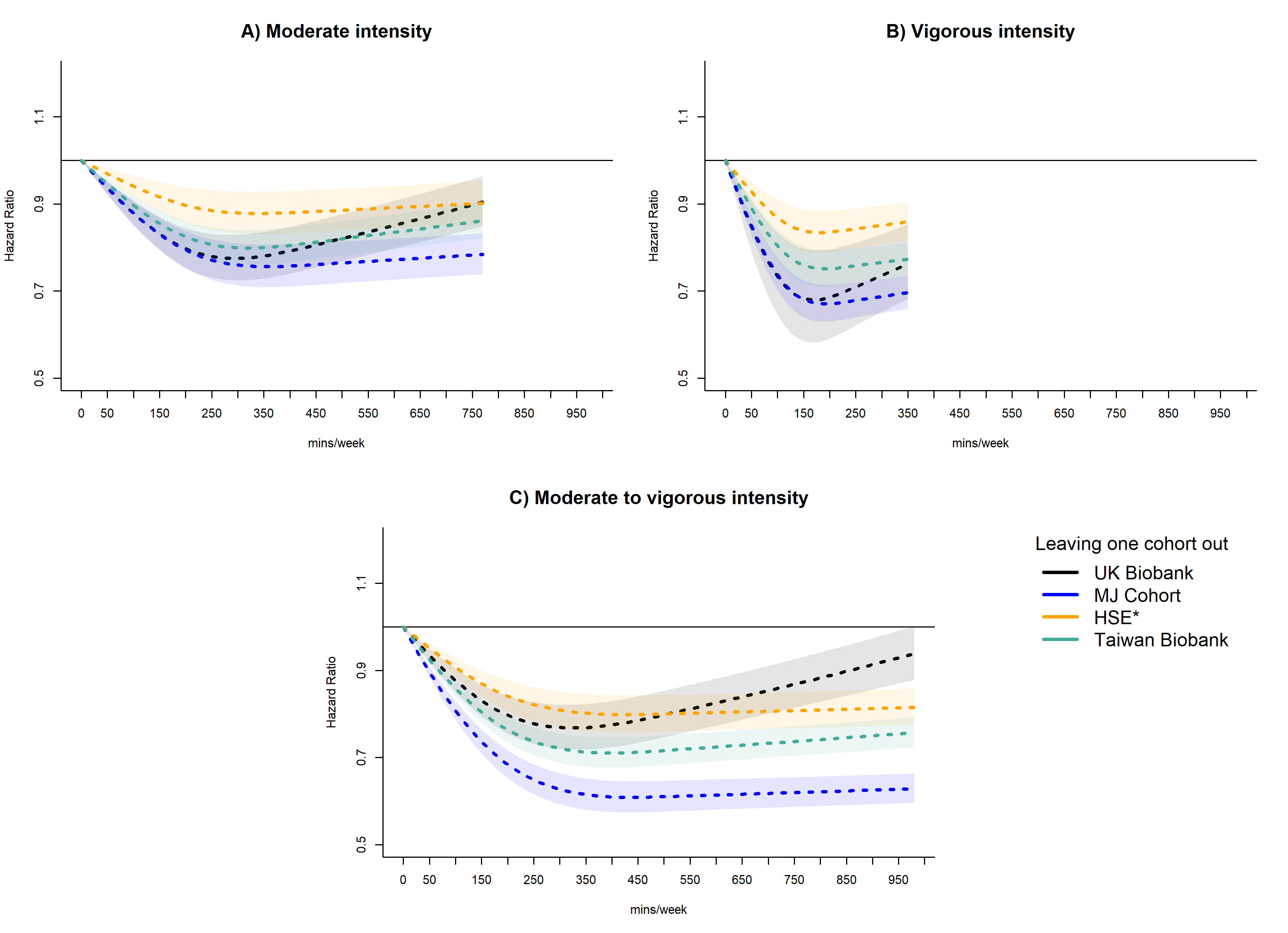
**Supplemental Figure 9: Leave-one-cohort-out dose response association with cardiovascular disease mortality**

Adjusted for age, sex, smoking status, alcohol consumption, body mass index, education, prevalent cancer. *Health Survey for England and Scottish Health Survey

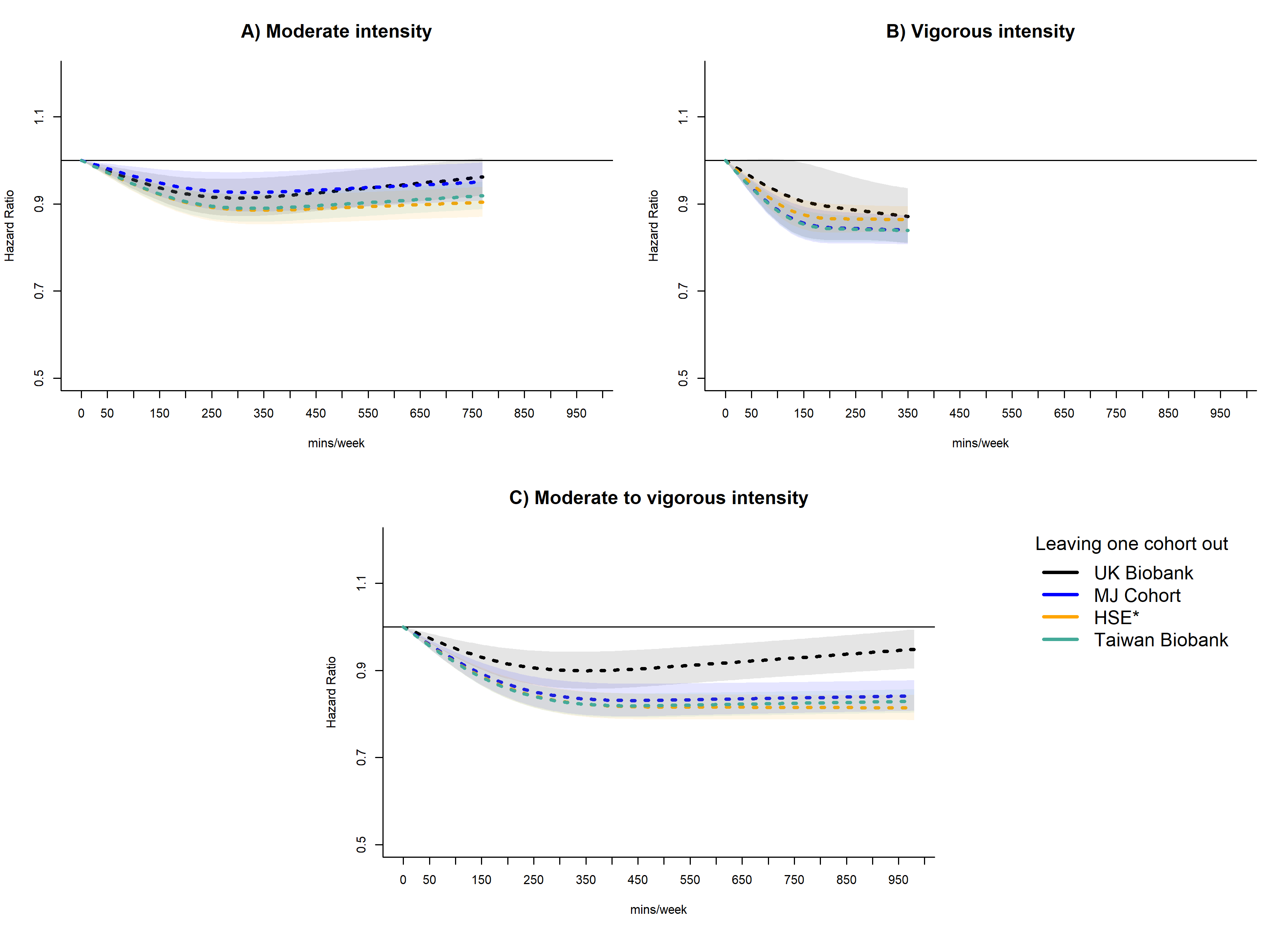
**Supplemental Figure 10: Leave-one-cohort-out dose response association with cancer mortality**

Adjusted for age, sex, smoking status, alcohol consumption, body mass index, education, prevalent cvd. *Health Survey for England and Scottish Health Survey
